# Alternative PFAS Are Associated with Ovarian Cancer–Related Lipidomic Remodeling and Cross-Matrix Metabolic Signatures

**DOI:** 10.64898/2026.09.24.26363829

**Authors:** Haohan Liu, Jing Ni, Na Gao, Cong Zhang, Xiuting Li, Nan Jiang, Shizhi Wang, Xiaoxiang Chen

## Abstract

Per- and polyfluoroalkyl substances (PFAS) are persistent environmental contaminants with the potential to disrupt metabolic processes and contribute to carcinogenesis; however, molecular evidence linking alternative and emerging PFAS to ovarian cancer (OC) remains limited. We conducted a biomarker-based case–control study integrating exposureomics with multi-omics profiling. Plasma samples from 128 patients with OC and 128 controls were analyzed for 26 PFAS, untargeted metabolomics, lipidomics, and clinical phenotypes, while urine samples from an additional 100 patients and 100 controls underwent untargeted metabolomic and clinical profiling. Single-compound, burden-based, and mixture models were integrated with PFAS–omics association scans, cross-matrix metabolite prioritization, observational exposure–omics–clinical association-chain analyses, and repeated internal cross-validation. Higher PFAS burdens were consistently associated with OC case status, and DONA, GenX, PFNA, PFPeS, PFUdA, and PFTrDA constituted an OC-prioritized six-PFAS mixture. DONA and GenX exhibited the broadest lipid-related bridging signals, particularly across phospholipids, lysophospholipids, sphingolipids, neutral lipids, and acylcarnitines. Integrated plasma and urine analyses further identified five shared metabolites—elaidic acid, hippuric acid, betaine, corticosterone, and taurocholic acid—that provided incremental and complementary discrimination beyond reference models based on clinical variables and tumor markers in internal cross-validation. Collectively, these findings demonstrate that both alternative and emerging PFAS are associated with a distinct OC-related molecular phenotype involving coordinated lipidomic, metabolomic, and hematologic alterations, while highlighting DONA and GenX as the most prominent exposure-related signals.

**Highlights:** • First epidemiologic evidence linked DONA and GenX to OC-related lipid remodeling.

• Single-compound, class-burden, and mixture models consistently linked PFAS to OC.

• Plasma–urine integration yielded a Core-5 signature beyond clinical markers.

• PFAS–omics–clinical networks highlighted lipid modules and RDW-SD phenotypes.

## 1 Introduction

Per- and polyfluoroalkyl substances (PFAS) comprise a broad class of synthetic organofluorine chemicals that have been extensively used in industrial processes and consumer products because of their thermal stability, surface-active properties, and resistance to degradation(*1*, *2*). However, these same physicochemical properties also contribute to their environmental persistence and mobility. The widespread detection of PFAS in drinking water, food, indoor environments, wildlife, and human biospecimens indicates pervasive and ongoing human exposure (*3*, *4*).

Regulatory and public health efforts have historically focused on legacy PFAS, particularly perfluorooctanoic acid (PFOA) and perfluorooctanesulfonic acid (PFOS). Following the phase-out and restriction of selected long-chain PFAS, shorter-chain and ether-based alternatives have been adopted as substitutes or processing aids in industrial applications. These compounds include hexafluoropropylene oxide dimer acid and its ammonium salt, commonly referred to as GenX chemicals, and 4,8-dioxa-3H- perfluorononanoic acid and its ammonium salt (DONA) (*2*, *5*).Their detection in surface waters across multiple countries and, for some novel drinking-water-associated PFAS, in human blood provides evidence of environmental dissemination and internal exposure (*5*, *6*). Nevertheless, epidemiologic evidence and human-relevant toxicologic data for most alternative and emerging PFAS remain limited, and their toxicokinetics, mixture effects, and long-term health consequences are incompletely characterized (*1*, *7*).

A growing body of experimental and epidemiologic evidence indicates that PFAS exposure may perturb lipid and bile acid metabolism, endocrine signaling, immune function, oxidative-stress pathways, hepatic homeostasis, and inflammatory responses; however, the strength and consistency of evidence differ substantially across individual PFAS and health outcomes (*1*, *8–10*). Several of these biological processes are relevant to carcinogenesis and to the systemic metabolic dysregulation associated with malignancy. However, direct evidence linking most PFAS—particularly alternative and emerging compounds—to cancer initiation or progression remains limited (*11*, *12*).

In 2023, the International Agency for Research on Cancer classified PFOA as carcinogenic to humans (Group 1), further underscoring the need to systematically evaluate the carcinogenic and metabolic hazards associated with environmentally relevant PFAS exposures (*13*).

Nevertheless, most population-based studies have focused on a limited number of legacy PFAS and conventional clinical outcomes (*3*, *12*, *12*). Whether alternative and emerging PFAS are associated with distinct molecular phenotypes, and whether these phenotypes can help clarify exposure-related disease patterns in human populations, remains poorly understood (*1*, *7*).

Ovarian cancer (OC) is among the most lethal gynecologic malignancies worldwide (*14*, *15*). The detection of PFAS in human follicular fluid indicates that these compounds can reach the ovarian microenvironment, consistent with their transfer across the blood–follicle barrier (*16*, *17*). Ovarian function is tightly regulated by endocrine signaling, immune and inflammatory processes, and lipid metabolism, all of which may be perturbed by PFAS exposure (*16*). A recent prospective nested case– control study using prediagnostic serum samples reported positive associations of PFOS and its precursor, 2-(N-methyl-perfluorooctane sulfonamido) acetic acid (MeFOSAA), with subsequent OC risk. Perfluorononanoic acid (PFNA) and perfluorodecanoic acid (PFDA) also showed suggestive associations, further supporting the need to investigate the potential role of PFAS exposure in ovarian malignancy (*18*).

Several important gaps remain from an environmental health perspective. First, molecular alterations associated with alternative and emerging PFAS in OC remain largely unknown (*18*, *19*). Second, PFAS exposures occur as correlated mixtures in real-world settings rather than as isolated compounds; consequently, single-pollutant models may not adequately capture the joint effects and correlation structure of these exposures (*20*). Third, molecular features connecting internal PFAS exposure biomarkers with the systemic metabolic and clinical phenotypes of OC have rarely been investigated and remain poorly defined (*21*).

High-resolution metabolomics and lipidomics provide powerful tools for characterizing the biological responses associated with PFAS exposure (*21*). Metabolites and lipids are downstream products of interactions among environmental exposures, host physiology, disease status, diet, microbial activity, and clinical conditions (*22*). Previous metabolomic and lipidomic studies of OC have revealed extensive alterations in fatty acids, phospholipids, lysophospholipids, sphingolipids, neutral lipids, amino acids, bile acids, and host–microbial co-metabolites (*23–27*). Population-based studies have also linked PFAS exposure to circulating lipid levels, cholesterol homeostasis, amino acid metabolic pathways, and broader metabolic disturbances (*21*, *28*). Together, these converging lines of evidence suggest that lipid and small-molecule metabolic phenotypes may represent an important molecular interface between PFAS exposure and OC-related systemic biological alterations.

Continued advances in multi-omics approaches offer opportunities to improve the characterization of these exposure-related molecular patterns. In particular, cross-matrix integration can provide complementary biological information and enhance the interpretability of identified features. Plasma metabolomics and lipidomics capture aspects of circulating metabolic status, molecular transport, inflammatory responses, and lipid remodeling, whereas urine metabolomics provides complementary information on metabolite excretion, renal handling, host–microbial co-metabolism, and systemic metabolic turnover (*29*).

Because individual metabolites often exhibit substantial matrix specificity, molecular features consistently detected in both plasma and urine may represent more robust candidates for subsequent targeted validation. Integrating exposureomics with plasma metabolomics, plasma lipidomics, urine metabolomics, and clinical phenotypes may therefore move beyond conventional exposure–disease association analyses and provide a more biologically informative framework for environmental health research (*30*).

We conducted a biomarker-based case–control study integrating exposureomics with multi-omics profiling. Using an UPLC–MS/MS platform, we quantified 26 PFAS in plasma, encompassing legacy, short-chain, long-chain, alternative, and emerging compounds. These exposure measurements were integrated with concurrently generated untargeted plasma metabolomic, plasma lipidomic, untargeted urine metabolomic, and routine clinical phenotype data.

To systematically evaluate associations at both the individual-compound and mixture levels, we applied single-compound models, grouped PFAS burden analyses, quantile g-computation, generalized weighted quantile sum regression, and Bayesian kernel machine regression (*20*, *31*). We further conducted differential omics analyses, PFAS– omics association scans, cross-matrix metabolite prioritization, and observational PFAS–omics–clinical bridging analyses to identify molecular features associated with PFAS exposure.

We hypothesized that plasma concentrations of alternative and emerging PFAS would be more strongly associated with OC case status than those of legacy PFAS and would coincide with coordinated lipidomic and metabolomic alterations. We further hypothesized that integrating plasma and urine metabolomic profiles would enable the prioritization of a concise set of shared cross-matrix metabolites reflecting exposure-related systemic metabolic phenotypes. By linking PFAS exposure biomarkers with variation in lipidomic, metabolomic, and clinical phenotypes, this study aimed to establish an integrated environmental molecular epidemiology framework for OC and to identify candidate biological pathways for further evaluation in prospective, mechanistic, and translational studies.

## 2 Materials and Methods

### 2.1 Study Design, Participants, and Biospecimen Collection

This case–control multi-omics study incorporated biomarker-based assessment of PFAS exposure and was jointly conducted by Southeast University and Jiangsu Cancer Hospital. Two complementary analytical datasets were included. Dataset 1 comprised 256 participants, including 128 patients with OC and 128 healthy controls, and was used for targeted quantification of plasma PFAS, untargeted plasma small-molecule metabolomics, plasma lipidomics, and clinical laboratory measurements. Dataset 2 comprised 200 participants, including 100 patients with OC and 100 healthy controls, and was used for untargeted urine small-molecule metabolomics and clinical laboratory measurements.

OC case status was treated as the primary binary outcome in all association analyses, bridging-path prioritization analyses, and internal prediction models. Among patients with OC, International Federation of Gynecology and Obstetrics (FIGO) stage was further categorized as stages I–II or III–IV for descriptive stage-stratified comparisons. The study protocol was approved by the Ethics Committee of Jiangsu Cancer Hospital (KY-2024-139). All participants provided written informed consent.

### 2.2 Clinical Data

Demographic characteristics, clinical laboratory measurements, comorbidities, tumor markers, and FIGO stage were collected using a standardized clinical data collection form. Continuous variables were summarized as the mean and standard deviation or the median and interquartile range, according to their distributions. Categorical variables were summarized as counts and percentages.

### 2.3 Targeted Quantification of Plasma PFAS

Plasma aliquots from Dataset 1 were used for targeted PFAS quantification. Materials and consumables free of PFAS contamination were used throughout blood collection, plasma separation, and sample storage. Contamination-control procedures were implemented with reference to the principles described in U.S. Environmental Protection Agency Methods 537.1 and 533.

A total of 26 PFAS were measured, encompassing legacy compounds, C4–C18 homologues, and alternative and emerging PFAS. Targeted analysis was performed using an ultra-performance liquid chromatography–triple quadrupole tandem mass spectrometry system (UPLC–MS/MS; Triple Quad 5500, AB Sciex) operated in multiple reaction monitoring (MRM) mode.

PFAS concentrations were quantified using isotope-labeled internal standards and external calibration curves. Only PFAS with detection frequencies of at least 90% in both cases and controls were retained for subsequent analyses.

Optimized MRM parameters, calibration performance, correlation coefficients, limits of detection, limits of quantification, and relative standard deviations (RSDs) are provided in Supplementary Tables S32–S33. Detailed information on reagent selection and sample preparation is presented in Supplementary Method S1 and in our previous publication (*32*).

### 2.4 Untargeted Metabolomics and Lipidomics

Plasma samples from Dataset 1 were also subjected to untargeted small-molecule metabolomic analysis. Case and control samples were randomized across sample-preparation batches and instrumental-analysis batches. A pooled quality-control (QC) sample was prepared by combining a 20-μL aliquot from each study sample. The QC sample was processed and analyzed alongside the study samples to monitor extraction reproducibility, instrument stability, and batch drift.

Metabolic features were acquired in positive and negative electrospray ionization modes using a Vanquish ultra-high-performance liquid chromatography system coupled to a Q Exactive Orbitrap high-resolution mass spectrometer (Thermo Fisher Scientific). Raw data were processed using Compound Discoverer v3.3 for feature detection, peak alignment, missing-peak filling, QC-based normalization, and metabolite annotation. Only features with a relative standard deviation (RSD) <30% in the QC sample and high-confidence metabolite annotations were retained for subsequent analyses. Detailed sample-preparation procedures and experimental conditions are provided in Supplementary Method S2. The LC–MS acquisition parameters and chromatographic gradient conditions have been fully described in our previous study (*33*).

Plasma aliquots from Dataset 1 were further subjected to lipidomic analysis to characterize OC case-related lipid remodeling and identify a candidate feature set for PFAS–lipid bridging. Lipids were extracted using a modified biphasic Matyash method (*34*) and analyzed using an ultra-high-performance liquid chromatography–Orbitrap high-resolution mass spectrometry system operated in full-scan/data-dependent tandem mass spectrometry (Full-MS/ddMS2) mode.

LipidSearch v5.1 was used for data deconvolution, peak extraction, lipid identification, and retention-time alignment. The mass spectra were subsequently reviewed manually to remove redundant annotations, isotope peaks, duplicate adducts, false-positive identifications, and structurally implausible lipid species, thereby improving the reliability of lipid annotation. Detailed sample-preparation procedures and experimental conditions are provided in Supplementary Method S3. The optimized LC– MS acquisition parameters and chromatographic gradient conditions have been fully described in our previous study (*35*).

Urine samples from Dataset 2 were subjected to untargeted small-molecule metabolomic analysis using the same UHPLC–HRMS workflow as that applied for plasma metabolomics. To reduce variation arising from differences in urine dilution, urinary metabolite signals were normalized to urinary creatinine before statistical analysis. Urinary creatinine concentrations were measured using an automated biochemical analyzer (AU5800, Beckman Coulter, Brea, CA, USA).

### 2.5 Clinical, PFAS, and Mixture Exposure Analyses

Logistic regression was used to evaluate associations between individual clinical indicators and OC case status. The clinical models were specified sequentially as follows: an unadjusted model (Model 1), an age-adjusted model (Model 2), a model adjusted for age and body mass index (BMI; Model 3), and a model further adjusted for age, BMI, and hypertension (Model 4). Effect estimates are reported as odds ratios (ORs) with 95% confidence intervals (CIs).

Descriptive analyses of PFAS included summaries of detection frequencies, distributions of standardized concentrations, Spearman correlation matrices, principal component and loading analyses, and exposure-burden summaries by PFAS class.

Detailed specifications of the clinical indicator–PFAS association models are provided in **Supplementary Method S4**.

To further investigate the joint associations of PFAS co-exposure with OC, we applied three complementary mixture-analysis approaches: quantile g-computation (qgcomp), generalized weighted quantile sum regression (gWQS), and Bayesian kernel machine regression (BKMR). All three approaches were adjusted for the covariates included in Model 4.

Briefly, PFAS concentrations were categorized into quartiles for both qgcomp and gWQS. The gWQS analysis used 100 bootstrap iterations, with 60% of the participants assigned to the validation dataset. BKMR was implemented using 20,000 Markov chain Monte Carlo iterations with variable selection. Additional implementation details are provided in Supplementary Method S5.

### 2.6 Differential Omics Analyses, Pathway Annotation, and Cross-Matrix Metabolite Prioritization

Case–control differences in plasma and urine small-molecule metabolites were evaluated using log₂-transformed abundance data. Differential metabolites were prespecified as those with *P* < 0.05 and |log₂FC| > log₂ (1.5), with Benjamini–Hochberg false discovery rate (BH-FDR) values reported in parallel. Differential analysis of plasma lipids applied the same log₂FC threshold and multiple-testing correction framework. Associations between individual lipids and OC case status were estimated using logistic regression.

Differential metabolites were annotated using the Kyoto Encyclopedia of Genes and Genomes (KEGG), and their functional distributions were visualized to characterize pathway-level alteration patterns. When metabolite–gene mapping information was available, Gene Ontology (GO) analysis was additionally performed as an exploratory functional annotation. Lipid pathway and lipid-class analyses were based on lipid annotations, KEGG identifiers, or lipid-class mappings derived from the lipid database implemented in LipidSearch v5.1.

To identify cross-matrix metabolite candidates that were not restricted to a single biospecimen matrix, differential plasma metabolites from Dataset 1 were matched against differential urine metabolites from Dataset 2. For metabolites shared across the two matrices, we compared the directions of case–control differences in plasma and urine, as well as the directions and magnitudes of covariate-adjusted logistic regression coefficients.

Shared differential metabolites were further prioritized using least absolute shrinkage and selection operator (LASSO) logistic regression. Each repeated analysis used stratified 10-fold cross-validation, and variables with nonzero coefficients at λ₁ₛₑ were considered selected. Selection frequencies of 30% to < 50% were defined as suggestive support, whereas frequencies ≥ 50% were defined as robust support.

Finally, cross-matrix differential status, concordance in effect direction, covariate-adjusted association evidence, and LASSO selection stability were jointly considered to define the Core-5 feature set, comprising elaidic acid, hippuric acid, betaine, corticosterone, and taurocholic acid.

### 2.7 PFAS–Omics Association Scanning and Candidate Feature Prioritization

Systematic PFAS–omics association analyses were first conducted for the six disease- priority PFAS: DONA, GenX, PFPeS, PFNA, PFUdA, and PFTrDA. For a more focused high-dimensional integration analysis, the four compounds with the largest numbers of FDR-significant lipid associations in Supplementary Table S17—DONA, GenX, PFNA, and PFPeS—were further selected to constitute the four-PFAS omics integration set.

Covariate-adjusted linear regression models were fitted with standardized log₂- transformed omics-feature abundances as the outcome variables and standardized log₂- transformed PFAS concentrations as the exposure variables, with adjustment for the covariates included in Model 4. Multiple testing was controlled using both global BH-FDR correction and PFAS-specific BH-FDR correction.

Candidate bridging molecular features were required to meet both of the following criteria: (1) association with OC case status or a difference in abundance between cases and controls; and (2) a statistical association with a PFAS exposure biomarker, with the direction of association consistent with the corresponding OC-related omics alteration.

PFAS–omics–case status association chains and PFAS–omics–clinical phenotype–case status association chains were evaluated using an observational product -of-coefficients framework combined with bootstrap resampling.

### 2.8 Observational Bridging Prioritization and Tandem Association-Chain Analyses

An observational product-of-coefficients framework was used to evaluate the PFAS– omics–case status and PFAS–omics–clinical phenotype–case status association chains. For the PFAS–lipid and PFAS–shared metabolite association chains, Model 4-adjusted linear regression models were fitted to estimate associations between PFAS and individual omics features, whereas logistic regression models were used to evaluate associations between individual omics features and OC case status.

Briefly, 300 bootstrap samples were generated by resampling participant-level records with replacement. Bootstrap samples that did not yield valid estimates were excluded, and an analysis was considered unsuccessful when fewer than 50 valid bootstrap estimates were obtained. For bootstrap-evaluated lipid pairs, BH correction was applied across all pairs. For shared-metabolite pairs, BH correction was performed within each analytical level and exposure. For tandem association chains, both global BH correction and BH correction stratified by the first feature type were applied.

### 2.9 Internal Prediction Modeling

Internal prediction models were further developed to evaluate whether the Core-5 cross-matrix shared metabolite feature set provided additional discriminatory ability beyond reference models based on routine clinical variables and tumor markers. Candidate feature sets included basic clinical variables, routine clinical indicators, a tumor marker positivity-based reference model, the Core-5 metabolites, and different combinations of these feature sets.

Detailed methods are provided in Supplementary Method S7. Briefly, unpenalized logistic regression with prespecified predictors, LASSO logistic regression, and elastic net logistic regression were evaluated using repeated stratified 5-fold cross-validation at the participant level; the penalized models were implemented using glmnet. To minimize information leakage, missing-value imputation, variable standardization, feature selection, and model fitting were performed entirely within each training fold. The resulting preprocessing parameters and fitted models were then applied to the corresponding validation fold.

XGBoost models were also developed as a sensitivity analysis using a nonlinear internal prediction approach. Model performance was evaluated using repeated stratified 5-fold cross-validation at the participant level, with early stopping implemented within each training fold.

### 2.10 Statistical Software

All analyses were conducted using R version 4.4.2. The primary R packages included tidyverse, readxl, tableone, mixOmics, qgcomp, gWQS, bkmr, glmnet, xgboost, and ComplexHeatmap. All statistical tests were two-sided. Unless otherwise specified, *P* < 0.05 was considered the threshold for nominal statistical significance, and BH-FDR correction was applied to control for multiple comparisons.

## 3 Results

### 3.1 Reproducible Clinical Phenotypic Differences between Controls and OC Cases

Using Dataset 1 (plasma) and Dataset 2 (urine), we established a multilevel analytical framework integrating clinical phenotypes, PFAS exposure profiles, plasma metabolomics and lipidomics, urine metabolomics, cross-matrix shared-metabolite prioritization, observational bridging association-chain analyses, and modeling with internal cross-validation. XGBoost sensitivity analyses were additionally performed to evaluate the stability of candidate discriminatory features across different modeling strategies (Figure 1A and Supplementary Tables S1–S6).

**Figure 1.**
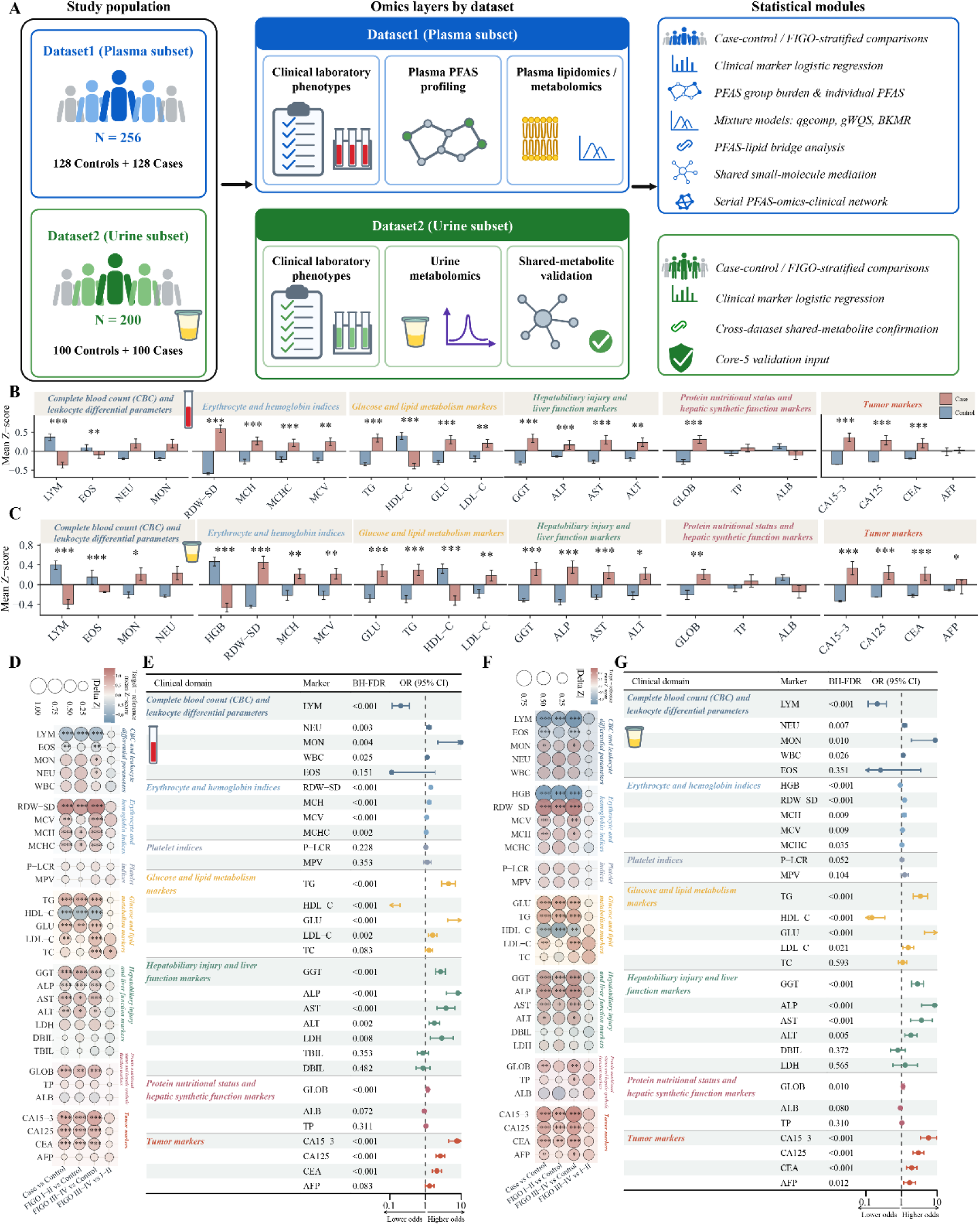
Clinical differences between OC cases and controls. (A) Study design and analytical framework. (B, C) Standardized mean levels of clinical measures in cases and controls in Dataset 1 (B) and Dataset 2 (C). Bars represent group mean z scores, and error bars represent SEs. Statistical tests were performed using the original values, followed by BH-FDR correction. (D, F) Standardized differences in clinical measures for the overall case–control comparison and FIGO stage-stratified comparisons in Dataset 1 (D) and Dataset 2 (F). Point color indicates the direction of the standardized mean difference, and point size represents −log₁₀(FDR). Asterisks denote statistically significant differences after multiple-testing correction. (E, G) Multivariable logistic regression associations between clinical measures and OC case status. Points represent ORs, horizontal lines represent 95% CIs, and the vertical dashed line indicates OR = 1. Models correspond to Model 3 and were adjusted for age and BMI. Sensitivity analyses corresponding to Model 4, with additional adjustment for hypertension, are shown in Supplementary Figures S1 and S9.

Age and BMI did not differ significantly between cases and controls in either biospecimen subset, indicating overall comparability in basic demographic characteristics (Supplementary Tables S1 and S4). Analyses of routine clinical measurements, however, revealed multidimensional differences between OC cases and controls, with generally consistent directions of change across Datasets 1 and 2.

Specifically, OC cases exhibited coordinated alterations across several clinically relevant domains, including leukocyte differentials, such as lymphocytes (LYM) and eosinophils (EOS); erythrocyte- and hemoglobin-related indices, including hemoglobin (HGB), red blood cell distribution width–standard deviation (RDW-SD), mean corpuscular hemoglobin (MCH), and mean corpuscular volume (MCV); glucose and lipid metabolism markers, including glucose (GLU), triglycerides (TG), high-density lipoprotein cholesterol (HDL-C), and low-density lipoprotein cholesterol (LDL-C); hepatobiliary function markers, including γ-glutamyl transferase (GGT), alkaline phosphatase (ALP), aspartate aminotransferase (AST), and alanine aminotransferase (ALT); indicators of protein and nutritional status, including total protein (TP), albumin (ALB), and globulin (GLOB); and tumor markers, including carbohydrate antigen 125 (CA125), carbohydrate antigen 15-3 (CA15-3), carcinoembryonic antigen (CEA), and alpha-fetoprotein (AFP) (Figures 1B–C).

Overall, lymphocyte-related measures were generally lower in OC cases, whereas selected inflammatory cell indices, measures of erythrocyte heterogeneity, glucose- and lipid-related variables, hepatobiliary function markers, and tumor markers, including CA15-3, CA125, and CEA, were elevated. These findings indicate broad OC-related phenotypic alterations involving immune and inflammatory status, metabolism, and hepatobiliary function.

After incorporating FIGO stage, multiple clinical variables differed not only between cases and controls overall but also across FIGO stage categories (Figures 1D and 1F; Supplementary Figures S1A–B and S9A–B; Supplementary Tables S2–S3 and S5–S6). Thus, the clinical phenotypic differences associated with OC were not confined to a single variable or biospecimen subset but formed a relatively consistent disease-related phenotypic pattern spanning inflammatory and immune, hematologic, glucose and lipid metabolic, hepatobiliary, and tumor-marker domains.

To further evaluate the independent associations between these clinical variables and OC case status, we fitted covariate-adjusted logistic regression models. Multiple routine laboratory indicators in both datasets were significantly associated with OC case status (Figures 1E and 1G and Supplementary Tables S7–S9). Overall, lymphocyte counts were inversely associated with OC case status, whereas monocyte and neutrophil counts, red blood cell distribution width, MCH, glucose- and lipid-related variables, hepatobiliary function markers, and tumor markers generally showed positive associations. Sensitivity analyses yielded similar association patterns, supporting the robustness of these clinical phenotypic differences across alternative model specifications (Supplementary Figures S1C and S9C).

Collectively, these findings delineated a reproducible host phenotypic context in OC cases, characterized by the co-occurrence of inflammatory, hematologic, metabolic, and hepatobiliary alterations. This clinical phenotypic structure provided an essential reference layer for subsequent analyses linking PFAS exposure biomarkers to plasma lipidomic remodeling, cross-matrix metabolic perturbations, and exposure-related molecular phenotypes.

### 3.2 Plasma PFAS Exposure Levels Were Associated with OC Case Status

In plasma samples from Dataset 1, 26 PFAS were quantified using UPLC–MS/MS, of which 20 with relatively high detection frequencies were retained for subsequent statistical analyses (Figure 2A). Overall, plasma PFAS profiles in the study population were characterized by broad detection coverage, heterogeneous composition patterns, and structured co-exposure features (Figures 2B–C and Supplementary Table S10). Both controls and OC cases were concurrently exposed to multiple PFAS, although the relative composition and standardized concentration distributions differed across compounds. Spearman correlation analysis revealed significant positive correlations among multiple PFAS, particularly among compounds belonging to similar chemical classes or potentially sharing common exposure sources (Figure 2D). Principal component loading analysis further showed that the PFAS exposure profiles exhibited clustering patterns and lower-dimensional structures related to chemical class (Figure 2F and Supplementary Figure S3A). These findings indicated that participants were exposed to mixtures of multiple correlated PFAS, supporting the use of PFAS class- burden and multipollutant mixture analyses in addition to single-compound models. In multivariable logistic regression models, higher PFAS class-specific burdens were significantly associated with greater odds of OC case status. In Model 4, per 1-SD increase in the log₂-transformed burden, the OR was 1.932 (95% CI: 1.430–2.611) for total PFAS, 1.754 (95% CI: 1.309–2.350) for perfluoroalkyl carboxylic acids (PFCAs), 1.416 (95% CI: 1.074–1.866) for perfluoroalkyl sulfonic acids (PFSAs), 2.564 (95% CI: 1.844–3.566) for alternative PFAS, 1.624 (95% CI: 1.220–2.163) for legacy PFAS, and 2.278 (95% CI: 1.663–3.119) for emerging alternative PFAS (Figure 2G and Supplementary Table S11). These results indicated that alternative and emerging alternative PFAS were the exposure categories most strongly associated with OC case status in this case–control population.

**Figure 2.**
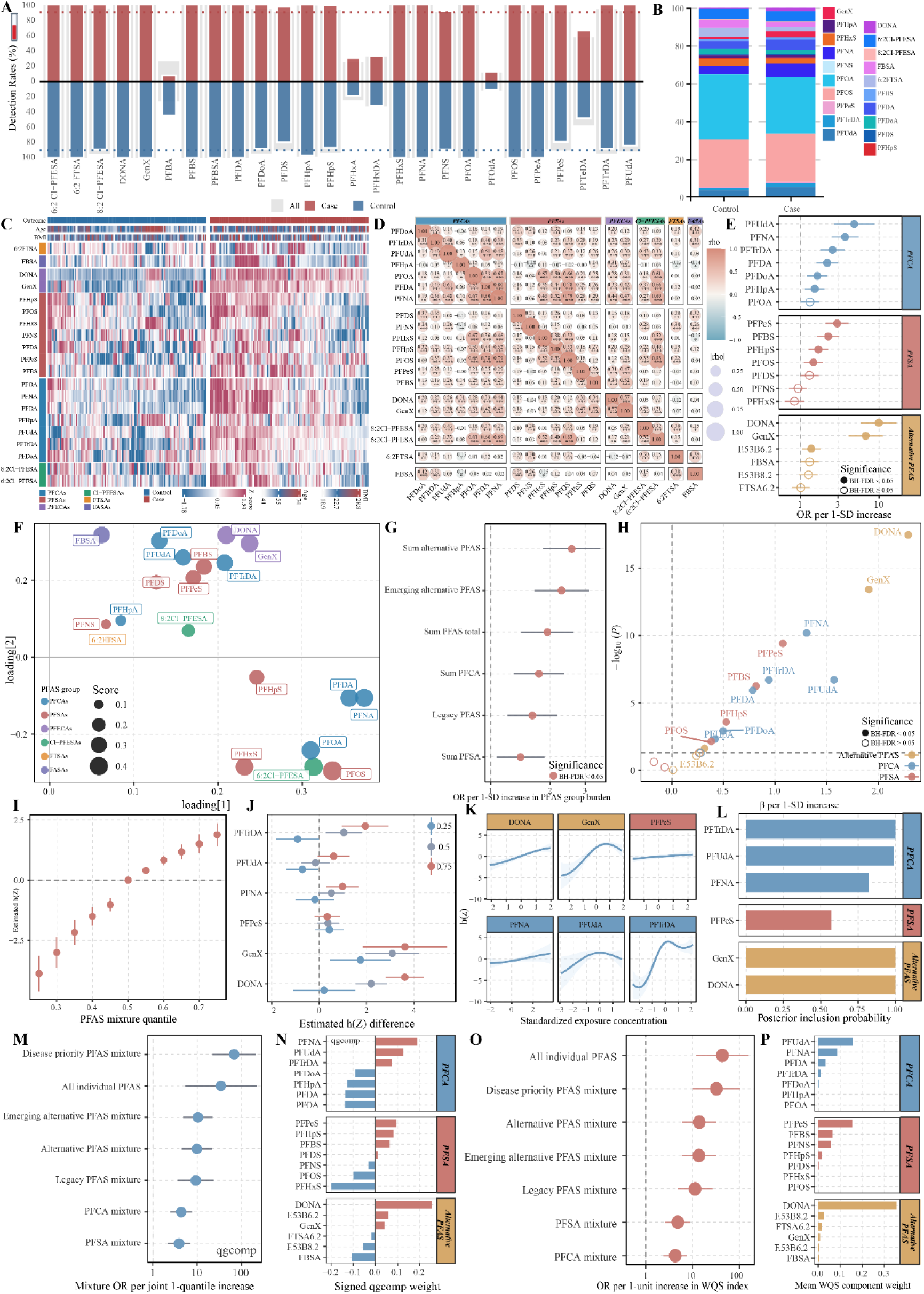
Plasma PFAS exposure profiles and associations with OC case status. (A) Detection frequencies of 26 PFAS in plasma samples from Dataset 1, shown for the overall study population, OC cases, and controls. (B) Stacked bar plots showing the relative PFAS composition in OC cases and controls. (C) Hierarchically clustered heatmap of plasma PFAS exposure profiles in Dataset 1. Side annotations indicate OC case status, age, BMI, and PFAS class. (D) Spearman correlation matrix among PFAS. Color indicates the direction of correlation, point size represents correlation strength, and asterisks denote statistical significance after FDR correction. (E) Multivariable logistic regression associations between individual PFAS and OC case status. (F) PCA loading plot of PFAS exposure profiles. Point size represents the absolute loading value, and color indicates PFAS class. (G) ORs for associations between PFAS class-specific exposure burdens and OC case status. (H) Volcano plot of associations between individual PFAS and OC case status. The x- axis represents the adjusted logistic regression coefficient (β) per 1-SD increase in the log₂-transformed PFAS concentration, and the y-axis represents −log₁₀(*P*). Solid points indicate BH-FDR < 0.05. (I–P) Associations of PFAS mixtures with OC case status and the contributions of individual mixture components, evaluated using BKMR (I–L), qgcomp (M, N), and gWQS (O, P). For qgcomp, the effect estimate corresponds to a joint one-quantile increase across all mixture components; for gWQS, it corresponds to a one-unit increase in the weighted quantile index.

Single-PFAS analyses identified six compounds that constituted the OC-prioritized mixture. In Model 4, DONA (OR, 9.881; 95% CI: 5.896–16.560), GenX (OR, 6.742; 95% CI: 4.112–11.053), PFUdA (OR, 4.812; 95% CI: 2.663–8.696), PFNA (OR, 3.691; 95% CI: 2.495–5.462), PFPeS (OR, 2.931; 95% CI: 2.093–4.104), and PFTrDA (OR, 2.558; 95% CI: 1.795–3.644) showed relatively large effect estimates, with BH-FDR values < 0.001 for all six compounds (Figures 2E and 2H and Supplementary Tables S12–S13). These associations remained positive across Models 1–4 (Supplementary Figures S3B–C).

To characterize the overall association of PFAS mixtures and the contributions of individual components under co-exposure conditions, we applied qgcomp, gWQS, and BKMR, with adjustment for age, BMI, and hypertension. Both qgcomp and gWQS showed positive associations between PFAS mixtures and OC case status, with the six- PFAS disease-priority mixture yielding the largest effect estimates [qgcomp: OR, 68.107; 95% CI: 22.138–209.533; positive-direction gWQS model: OR, 32.270; 95% CI: 10.175–102.346; both *P* < 0.001] (Figures 2I–K, 2M, and 2O; Supplementary Figure S3D and Supplementary Table S15). DONA received a relatively high weight in both the qgcomp and gWQS analyses, whereas BKMR estimated posterior inclusion probabilities (PIPs) of 1.000 for DONA, GenX, and PFTrDA (Figures 2L, 2N, and 2P and Supplementary Table S14).

Collectively, plasma PFAS exposure biomarkers were associated with OC case status across single-compound, class-burden, and mixture models. DONA, GenX, PFUdA, PFNA, PFPeS, and PFTrDA jointly constituted the six-PFAS OC-prioritized mixture. These findings established an exposureomics basis for subsequent analyses linking PFAS exposure biomarkers to metabolic perturbations in plasma and urine and to OC-related systemic molecular phenotypes.

### 3.3 Plasma Metabolomic and Lipidomic Profiles Revealed Systemic Remodeling Associated with OC

In Dataset 1, untargeted plasma metabolomics was further used to characterize the overall metabolic differences between OC cases and controls. The results revealed widespread perturbations in the plasma metabolome of OC cases (Figure 3A and Supplementary Figures S2A–B). PLS-DA variable-importance analysis further showed that metabolites contributing substantially to the separation between cases and controls were involved in lipid, amino acid, glucose, and inflammation-related metabolic processes (Figure 3B).

**Figure 3.**
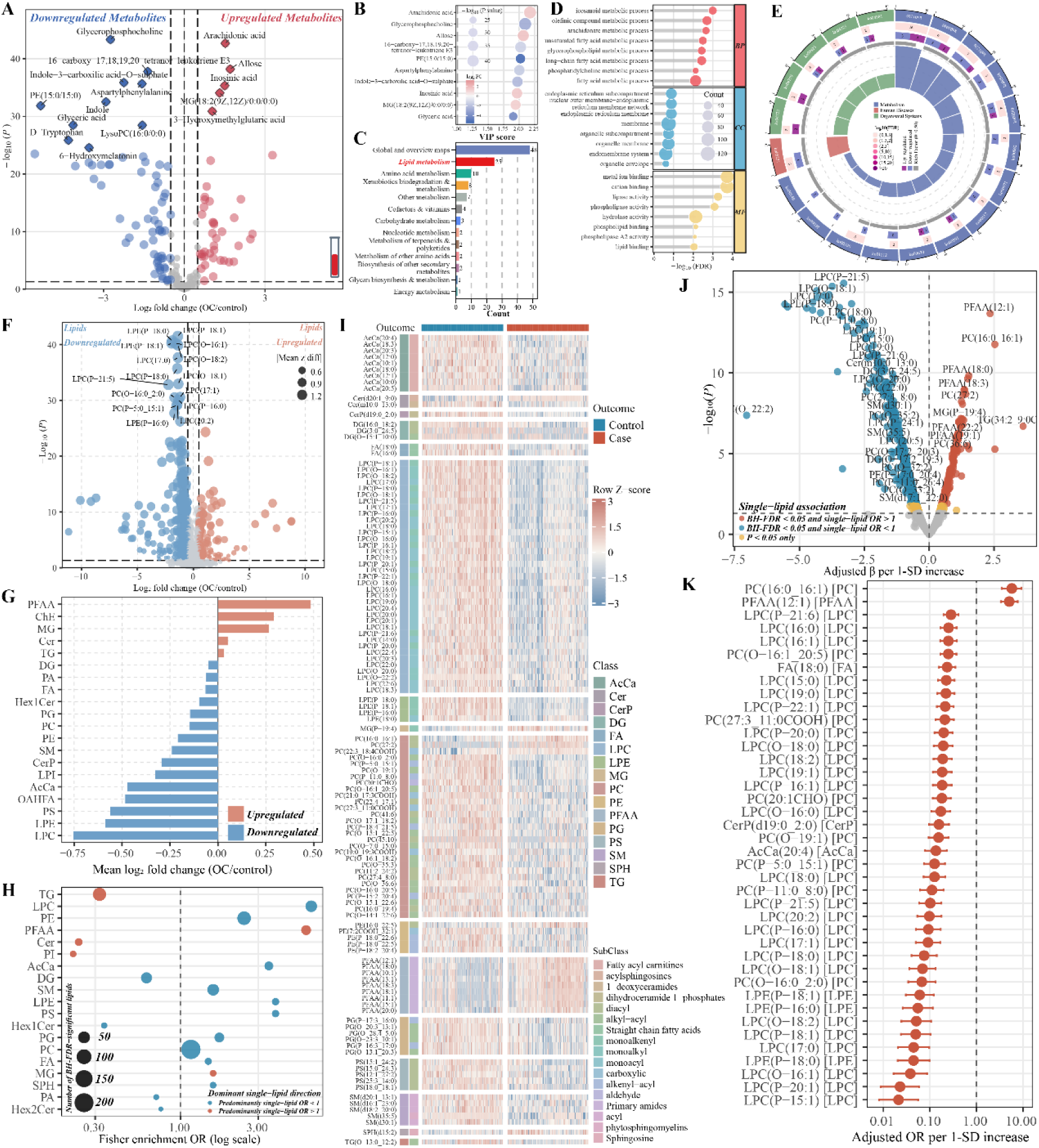
Plasma metabolomics and lipidomics revealed systemic metabolic remodeling associated with OC. (A) Volcano plot of differential plasma metabolites between OC cases and controls. The x-axis represents log₂FC (cases/controls), and the y-axis represents −log₁₀(*P*). (B) Representative plasma metabolites with the highest VIP scores in the PLS-DA model and the largest contributions to case–control separation. (C) KEGG category distribution of differential plasma metabolites. (D, E) Overview of GO (D) and KEGG (E) enrichment annotations for differential plasma metabolites. (F) Volcano plot of differential plasma lipids between OC cases and controls. (G, H) Lipid class-level summaries of plasma lipid alterations. (G) Bar plot showing the overall direction of abundance changes within each lipid class in OC cases. (H) Bubble plot showing the distribution of differential lipids across lipid classes; bubble size represents the number of differential lipids, and color indicates the predominant direction of individual lipid changes within each class. (I) Hierarchically clustered heatmap of plasma lipidomic profiles ordered by OC case status. Side annotations indicate lipid classes. (J) Volcano plot of associations between individual lipids and OC case status. The x- axis represents the adjusted logistic regression coefficient (β) per 1-SD increase in the log₂-transformed lipid signal intensity, and the y-axis represents −log₁₀(*P*). (K) Forest plot showing adjusted ORs and 95% CIs for selected OC-associated lipid species. ORs correspond to a 1-SD increase in the log₂-transformed lipid signal intensity and are displayed on a logarithmic scale.

Metabolite-class annotation showed that lipid metabolism-related features accounted for the largest proportion of differential metabolites. Other represented categories included amino acid metabolism, xenobiotic biodegradation and metabolism, carbohydrate metabolism, and cofactor and vitamin metabolism (Figure 3C).

Functional enrichment analysis further indicated that the differential plasma metabolites were predominantly enriched in biological processes related to fatty acid metabolism, carboxylic acid metabolism, monocarboxylic acid metabolism, long-chain fatty acid metabolism, phosphatidylcholine metabolism, lipid transport, lipid binding, and fatty acid binding (Figures 3D–E). These findings indicated extensive remodeling of the plasma metabolome in OC cases, with lipid-related metabolism representing one of the most prominent altered modules.

Given the pronounced lipid-related signals observed in the plasma metabolome, we further performed systematic lipidomic profiling of plasma samples from Dataset 1. Differential lipidomic analysis revealed widespread differences in individual lipid abundances between OC cases and controls, with decreased lipid abundances predominating in cases (Figure 3F). At the lipid-class level, lysophosphatidylcholine (LPC), lysophosphatidylethanolamine (LPE), phosphatidylcholine (PC), phosphatidylethanolamine (PE), sphingomyelin (SM), ceramide-1-phosphate (CerP), lysophosphatidylinositol (LPI), phosphatidylserine (PS), O-acyl-ω-hydroxy fatty acids (OAHFAs), acylcarnitines (AcCa), fatty acids (FA), phosphatidic acid (PA), and diacylglycerols (DG) species generally showed lower abundances in cases. In contrast, primary fatty acid amides (PFAA), cholesteryl esters (ChE), monoacylglycerols (MG), ceramides (Cer), and triacylglycerols (TG) species were relatively elevated (Figures 3G–H). The lipidomic heatmap ordered by case–control status further showed distinct modular patterns across individual samples, with coordinated abundance changes in multiple lipid classes between cases and controls (Figure 3I). These findings suggested that OC-related lipid alterations were not randomly distributed but instead exhibited lipid class-specific remodeling patterns.

We subsequently fitted multivariable logistic regression models adjusted for age and BMI to evaluate associations between individual lipid features and OC case status. Multiple lipid species remained significantly associated with case status (Figures 3J –K and Supplementary Figures S5A–C). Most significant lipids had ORs below 1, including multiple LPC, LPE, PC, SM, and CerP species, whereas selected primary fatty acid amide, MG, Cer, and PC species had ORs above 1. This directional heterogeneity indicated that OC-related lipid alterations depended on both lipid class and individual molecular species.

Collectively, the plasma metabolomic and lipidomic findings from Dataset 1 consistently indicated systemic perturbations of lipid-related metabolism in OC cases. Plasma metabolomics identified lipid metabolic abnormalities at the pathway and metabolite-class levels, whereas lipidomics further resolved this pattern at the levels of lipid classes, individual molecular species, and covariate-adjusted associations. These disease-related lipidomic features provided a molecular context for the subsequent identification of PFAS-associated lipid alterations and the evaluation of observational PFAS–lipid–OC bridging relationships.

### 3.4 Urine Metabolomics Prioritized Cross-Matrix Shared Metabolites Associated with OC

In Dataset 2, untargeted urine small-molecule metabolomics was further used to evaluate OC-related metabolic perturbations. Differential and dimensionality-reduction analyses revealed widespread alterations in the urinary metabolome of OC cases (Figure 4A and Supplementary Figures S10A–C). Pathway-level enrichment analysis further indicated that the differential urinary metabolites involved multiple metabolic modules, including lipid-related metabolism, steroid hormone biosynthesis, cortisol synthesis and secretion, bile acid-related metabolism, amino acid metabolism, xenobiotic metabolism, and energy metabolism (Figure 4B). These findings indicated that urine metabolomics captured systemic metabolic perturbations associated with OC and partially recapitulated the plasma metabolomic findings related to lipid- and hormone-associated pathways.

**Figure 4.**
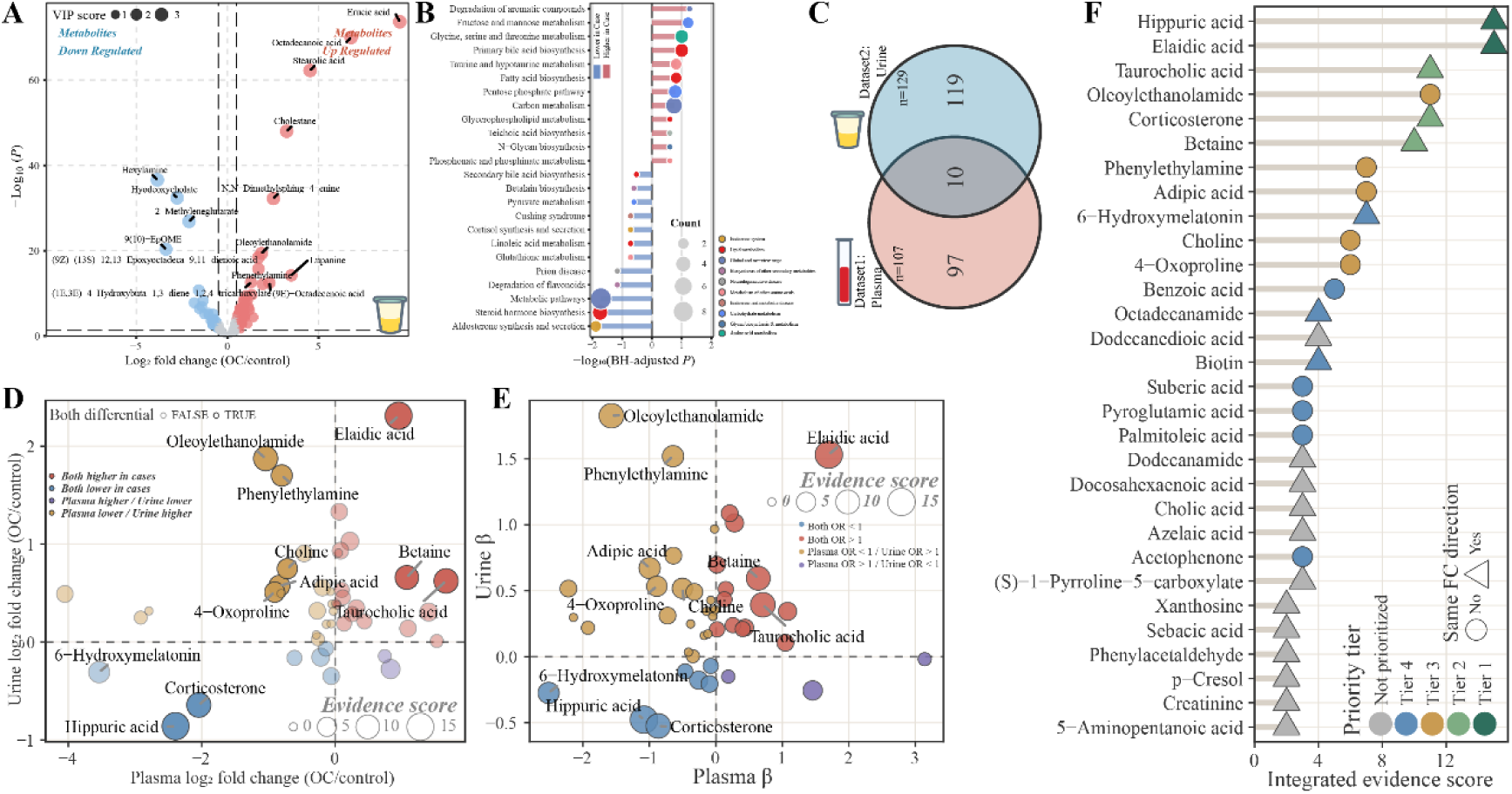
Urine metabolomics and cross-matrix prioritization of shared metabolites associated with OC. (A) Volcano plot of differential urinary metabolites between OC cases and controls. (B) KEGG pathway enrichment analysis of differential urinary metabolites. (C) Overlap between differential urinary metabolites (n = 129) and differential plasma metabolites (n = 107). (D, E) Cross-matrix comparisons of metabolites with matched annotations in plasma and urine, highlighting shared differential metabolites. (D) Scatterplot comparing case–control log₂FC values in plasma and urine. (E) β–β plot comparing Model 4 logistic regression coefficients in plasma and urine. In both panels, point size represents the weighted composite evidence score. (F) Integrated evidence scores for metabolites with matched cross-matrix annotations. The score incorporated shared differential status across matrices, concordance of log₂FC directions, cross-matrix association evidence, concordance of β-coefficient directions, and stability in repeated LASSO analyses. LASSO used stratified 10-fold cross-validation repeated 100 times, with covariates included as unpenalized terms and nonzero coefficients at λ₁ₛₑ defining variable selection. Selection frequencies of 30% to < 50% were classified as suggestive support, and frequencies ≥ 50% as robust support. The Core-5 feature set comprised elaidic acid, hippuric acid, betaine, corticosterone, and taurocholic acid.

To identify candidate metabolites that were less dependent on a single biospecimen matrix, we integrated the urine metabolomic data from Dataset 2 with the plasma metabolomic data from Dataset 1. Based on the prespecified criteria of *P* < 0.05 and |log₂FC| > log₂(1.5), 129 differential urinary metabolites and 107 differential plasma metabolites were identified, yielding 10 shared differential metabolites across the two matrices (Figure 4C). Metabolites exhibiting differential signals in both plasma and urine may represent more reproducible candidate features associated with OC case status.

We subsequently evaluated the cross-matrix concordance of these shared metabolites in terms of both differential abundance and adjusted associations. Several shared metabolites showed concordant directions of change in plasma and urine. Elaidic acid, betaine, and taurocholic acid were elevated in OC cases in both matrices, whereas hippuric acid and corticosterone were generally decreased (Figure 4D). Covariate-adjusted β–β mapping further classified the shared metabolites into quadrants representing directionally concordant and discordant association patterns (Figure 4E). These findings suggested that cross-matrix metabolite prioritization should jointly consider effect direction, covariate-adjusted association evidence, cross-matrix concordance, and model stability.

To further refine the shared-metabolite set, we performed 100 repeated LASSO analyses of the 10 shared differential metabolites, with covariates included as unpenalized variables for adjustment (Supplementary Figure S11A). Variables with nonzero coefficients at λ₁ₛₑ were considered selected. Selection frequencies of 30% to < 50% were defined as suggestive support, whereas frequencies ≥ 50% were defined as robust support. An integrated evidence score was subsequently constructed by combining differential status, directional concordance, covariate-adjusted logistic regression evidence, and LASSO selection stability. On this basis, elaidic acid, hippuric acid, betaine, corticosterone, and taurocholic acid were prioritized as the Core-5 cross-matrix shared metabolite feature set (Figure 4F).

Overall, urine metabolomics provided independent support for OC-related small-molecule metabolic perturbations. Integration with plasma metabolomics further refined the candidate features from matrix-specific differential metabolites to a more stable and biologically interpretable set of shared cross-matrix metabolites. These findings provided a basis for subsequent evaluation of PFAS-related cross-matrix metabolic phenotypes, construction of PFAS–metabolite–OC case status association chains, and future targeted validation.

### 3.5 Priority PFAS Were Associated with Plasma Small-Molecule and Lipid Perturbations

In Dataset 1, we further evaluated whether PFAS exposure biomarkers were associated with OC-related plasma metabolic perturbations and mapped the metabolic pathways underlying these associations. Linear regression models were fitted to assess PFAS– metabolite associations, followed by pathway enrichment analysis. Multiple PFAS were associated with a broad range of plasma small-molecule metabolites. Lipid metabolism remained the most prominent metabolic module, with additional associations involving amino acid metabolism, xenobiotic metabolism, carbohydrate metabolism, and cofactor and vitamin metabolism (Figure 5A).

**Figure 5.**
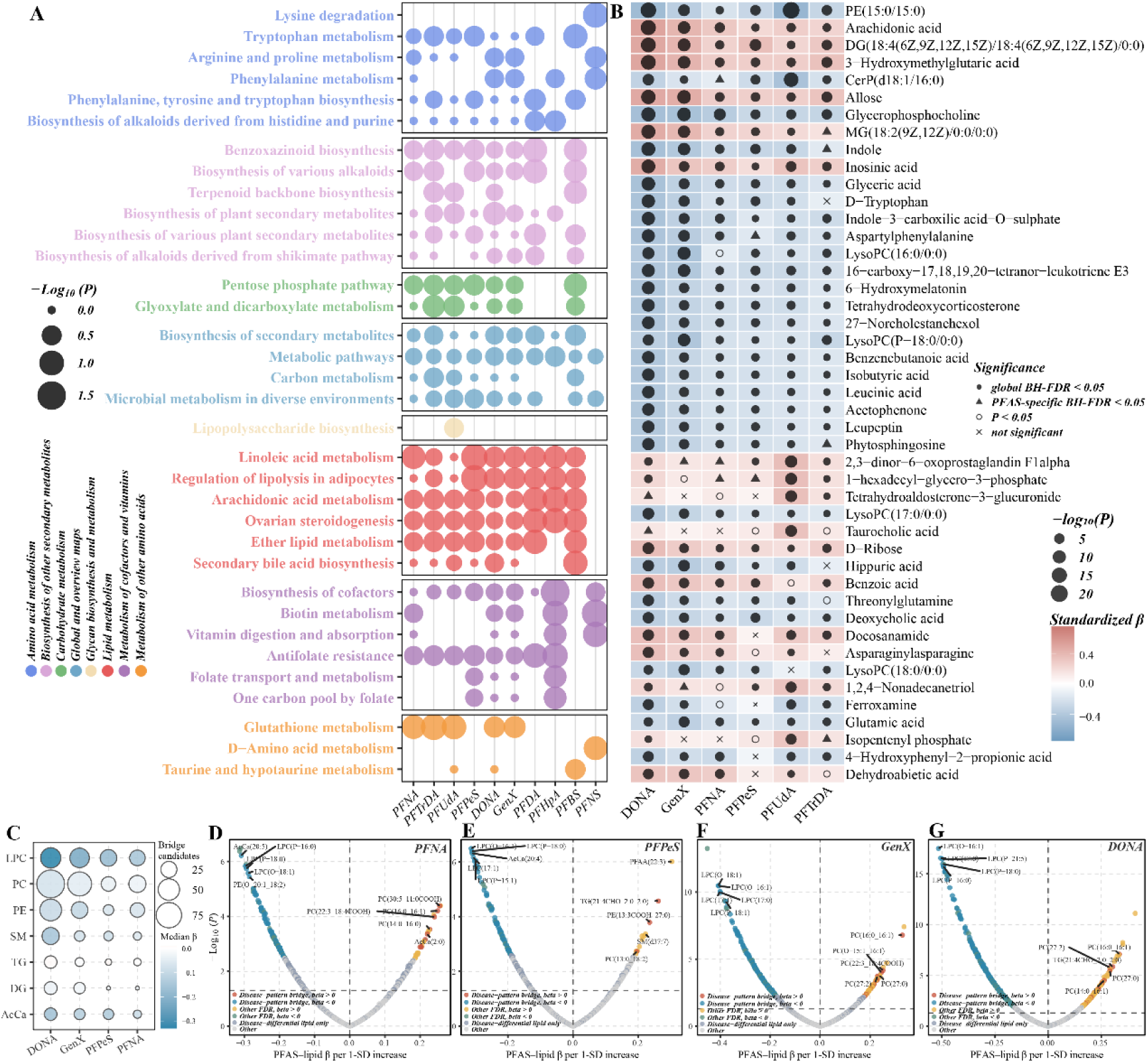
Observational PFAS–omics associations across the six-PFAS disease-priority scan and the four-PFAS omics integration set. (A) KEGG pathways enriched among plasma metabolites associated with DONA, GenX, PFPeS, PFNA, PFUdA, and PFTrDA. Point size represents −log₁₀(*P*), and color indicates the KEGG pathway category. (B) Covariate-adjusted heatmap of plasma metabolite associations in the six-PFAS disease-priority scan. Tile color represents the adjusted linear regression coefficient (β), defined as the SD change in the log₂-transformed metabolite signal per 1-SD increase in the log₂-transformed PFAS concentration. Point size represents −log₁₀(*P*), and point shape indicates the evidence tier: global BH-FDR < 0.05, PFAS-specific BH-FDR < 0.05, nominal *P* < 0.05, or no statistical support. (C) Lipid class overview for the four-PFAS omics integration set comprising DONA, GenX, PFNA, and PFPeS. Bubble size represents the number of candidate bridging lipids, and color indicates the median adjusted linear regression coefficient (β) for PFAS–lipid associations. (D–G) Volcano plots of PFAS–lipid associations for PFNA (D), PFPeS (E), GenX (F), and DONA (G). Candidate bridging lipids were required to be associated with both OC case status and PFAS exposure, with the direction of the PFAS–lipid association concordant with the corresponding OC-related lipid alteration.

The heatmap of metabolite associations with the six disease-priority PFAS further revealed both shared and heterogeneous patterns. For example, arachidonic acid was generally positively associated with multiple PFAS, whereas glycerophosphocholine was generally inversely associated with several priority PFAS. Other metabolites, including taurocholic acid, exhibited more heterogeneous association patterns across individual PFAS (Figure 5B). Thus, covariate-adjusted PFAS–metabolite associations showed both common and compound-specific patterns across the priority PFAS. Representative scatterplots further illustrated the directions, data dispersion, and approximate linearity of selected positive and inverse PFAS–metabolite associations (Supplementary Figure S4). These findings indicated structured associations between PFAS exposure biomarkers and OC-related metabolic alterations and provided a basis for the subsequent prioritization of shared cross-matrix metabolites and exposure-related metabolic features.

Given that both plasma metabolomics and lipidomics highlighted lipid-related pathways, we further conducted PFAS–lipid association scans for the six-PFAS disease- priority set to identify candidate lipidomic bridging features. DONA, GenX, PFNA, and PFPeS had the largest numbers of FDR-significant lipid associations and were therefore selected to form the four-PFAS omics integration set for subsequent focused high-dimensional analyses (Figures 5C–G and Supplementary Table S17).

DONA was associated with 513 lipids after FDR correction, of which 258 met the criteria for candidate bridging lipid features. GenX was associated with 242 FDR-significant lipids, including 165 candidate bridging features (Figures 5D–G and Supplementary Table S17). PFNA and PFPeS showed FDR-significant associations with 120 and 95 lipids, respectively, whereas PFUdA and PFTrDA were associated with 54 and 43 lipids, respectively. To reduce the multiple-testing burden and model complexity in downstream analyses, PFUdA and PFTrDA were not included in the subsequent focused integration analyses. Candidate bridging lipid features were predominantly distributed across LPC, PC, PE, SM, TG, DG, and acylcarnitine classes (Supplementary Tables S18–S20).

Representative scatterplots further illustrated the covariate-adjusted directions and participant-level distributions of selected PFAS–lipid associations. In particular, DONA-related signals involved multiple membrane lipids, sphingolipids, neutral lipids, and acylcarnitines (Supplementary Figure S6).

Overall, the four-PFAS omics integration set was associated with variation in both the plasma metabolome and lipidome. DONA and GenX showed the broadest sets of candidate bridging lipid associations. These observational findings provided a basis for the subsequent integrated analysis of PFAS–lipid–metabolite–clinical phenotype association chains.

### 3.6 PFAS–Omics–Clinical Integration Identified Multilayer Candidate Association Structures

To establish exposure–omics–clinical links, we further prioritized multilayer observational association chains using product-of-coefficients estimates and bootstrap resampling, thereby exploring potential connections between the four-PFAS omics integration set, OC-related omics features, and clinical phenotypes.

We first evaluated PFAS–lipid–OC association structures. DONA, GenX, PFNA, and PFPeS formed candidate associations with multiple OC-related lipid species. At the lipid-class level, supported associations were concentrated in LPC, PC, PE, SM, DG, TG, and acylcarnitines, with particularly prominent signals involving LPC and PC. DONA and GenX yielded more candidate associations than PFNA and PFPeS (Figures 6A, 6C, and 6H; Supplementary Figure S8). These findings suggested relatively consistent observational links between the four-PFAS omics integration set and OC-related lipid remodeling, particularly within lysophospholipid- and membrane lipid-related modules.

**Figure 6.**
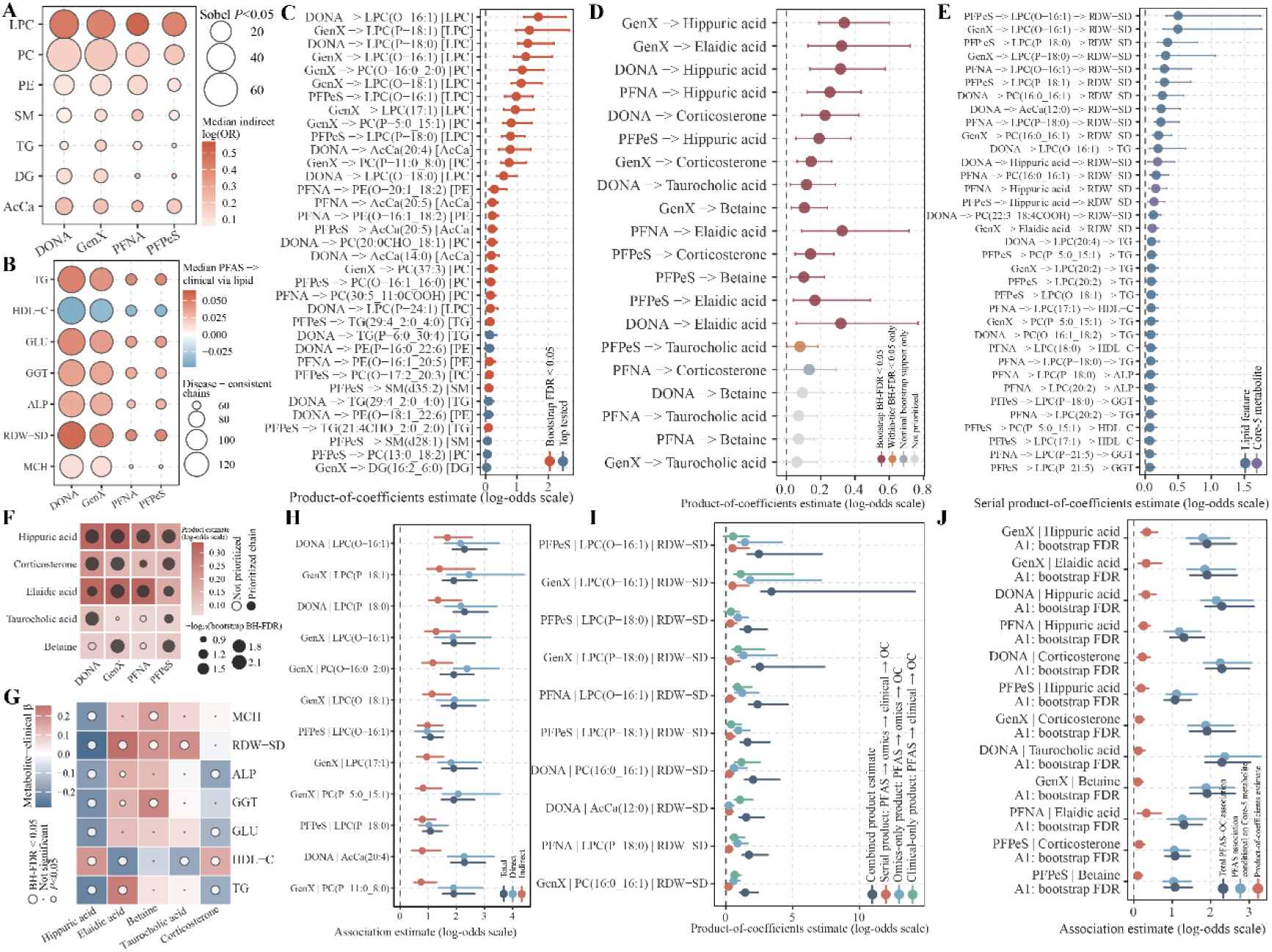
Observational PFAS–omics–clinical association chains related to OC case status. (A) Lipid class overview of candidate PFAS–lipid–OC case status association chains. Bubble size represents the number of chains with Sobel *P* < 0.05, and color indicates the median product-of-coefficients estimate on the log-odds scale. (B) Overview of association structures linking PFAS to clinical measures through lipid features. (C–E) Top-ranked PFAS–lipid–OC case status chains (C), PFAS–shared metabolite– OC case status chains (D), and tandem PFAS–omics feature–clinical phenotype–OC case status chains (E). Points represent product-of-coefficients estimates, and horizontal lines represent bootstrap 95% CIs. (F) Heatmap of product-of-coefficients estimates linking the four-PFAS omics integration set to OC case status through the Core-5 shared metabolites. Color represents the direction and magnitude of the estimate, point size represents −log₁₀- transformed bootstrap BH-FDR, and solid symbols indicate chains retained under the prespecified prioritization criteria. (G) Covariate-adjusted association heatmap between the Core-5 shared metabolites and OC-related clinical measures. Tile color represents the standardized regression coefficient (β), and symbols indicate statistical support after BH-FDR correction. (H) Decomposition of representative PFAS–lipid–OC case status association structures, showing the total PFAS–OC association, the PFAS conditional association after inclusion of the candidate lipid, and the product-of-coefficients estimate. Points represent estimates, horizontal lines represent bootstrap 95% CIs, and solid symbols indicate bootstrap BH-FDR < 0.05. (I) Decomposition of product-of-coefficients estimates for representative tandem association chains, showing the full tandem chain, the chain through the omics feature alone, and the chain through the clinical phenotype alone. (J) Decomposition of representative PFAS–Core-5 shared metabolite–OC case status association structures, showing the total PFAS–OC association, the PFAS conditional association after inclusion of the Core-5 metabolite, and the product-of-coefficients estimate.

We next evaluated observational association chains linking the four-PFAS omics integration set to OC case status through the Core-5 shared metabolites. After bootstrap resampling and BH-FDR correction, hippuric acid and elaidic acid received support in analyses of DONA, GenX, PFNA, and PFPeS, whereas corticosterone, betaine, and taurocholic acid showed more PFAS-specific support patterns (Figures 6D, 6F, and 6J; Supplementary Figure S12A and Supplementary Tables S21–S23).

To more comprehensively characterize multilayer PFAS–omics feature–clinical indicator–OC case status association structures, we further integrated the candidate omics features with OC-related clinical phenotypes. The association heatmap showed that the Core-5 metabolites were associated with TG, HDL-C, GLU, GGT, ALP, RDW-SD, and MCH, with heterogeneous directions of association (Figure 6G and Supplementary Table S24). Lipid–clinical indicator analyses likewise identified associations between multiple lipid features and these clinical measures. These associations were particularly concentrated among PC, LPC, PE, SM, and acylcarnitines and involved RDW-SD, GLU, GGT, ALP, HDL-C, and TG (Figure 6B, Figure 7B, and Supplementary Figures S7A–B).

**Figure 7.**
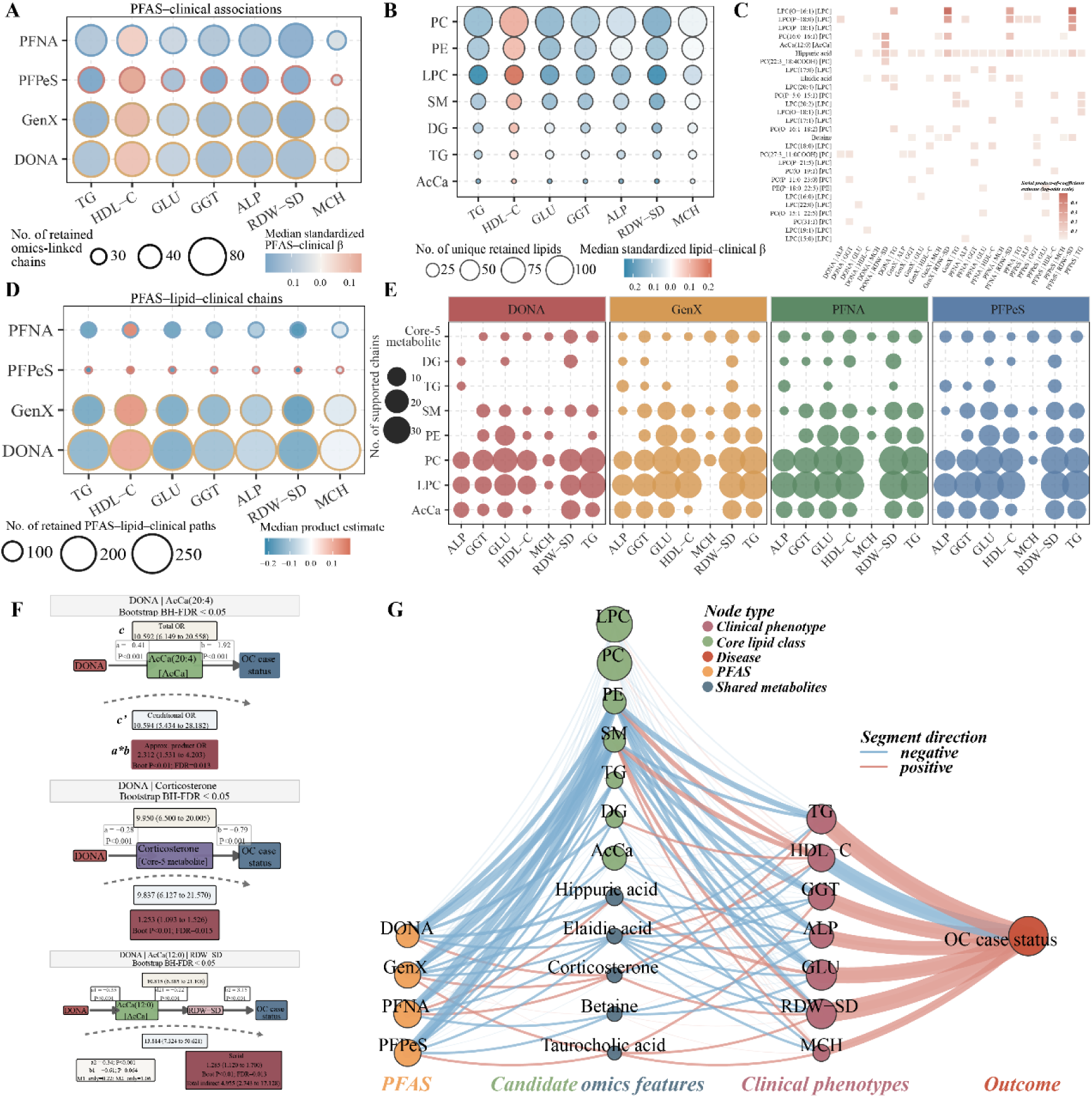
Integrated observational evidence network linking the four-PFAS omics integration set, candidate omics features, clinical phenotypes, and OC case status. (A) Retained association pairs between the four-PFAS omics integration set and OC- related clinical measures. Bubble size represents the number of retained omics association chains for each PFAS–clinical measure combination, and color indicates the median covariate-adjusted standardized linear regression coefficient (β) for the PFAS– clinical measure association. (B) Summary of associations between lipid classes and clinical measures. Bubble size represents the number of distinct retained lipid species, and color indicates the median linear regression coefficient (β). (C) Heatmap of product-of-coefficients estimates for tandem PFAS–omics feature– clinical phenotype–OC case status association chains. Color represents the direction and magnitude of the tandem estimate on the log-odds scale. (D) Retained PFAS–lipid–clinical measure association structures. (E) Supported association structures summarized by PFAS, omics feature, and clinical measure. Bubble size represents the number of supported structures. (F) Representative schematic of DONA-related lipids, Core-5 shared metabolites, and tandem association chains. The diagram shows the total PFAS–OC association, the PFAS conditional association after inclusion of the candidate omics feature, and the product-of-coefficients estimate, with results supported after bootstrap resampling and BH-FDR correction indicated. (G) Integrated observational evidence network connecting the four-PFAS omics integration set, candidate omics features, clinical phenotypes, and OC case status. Node color indicates node type, and node size represents the number of retained connections. Edge color indicates the direction of association, and edge width is proportional to the number of retained association chains contributing to the edge.

In the multilayer integration analysis, multiple retained association structures linked the four-PFAS omics integration set to clinical phenotypes and OC case status through core lipid or shared-metabolite modules. DONA and GenX showed the broadest connectivity within PFAS–lipid–clinical indicator associations, whereas PFNA and PFPeS retained fewer signals despite generally consistent directions (Figures 7A and 7D). Stratified summaries by PFAS, omics module, and clinical indicator showed that supported associations were concentrated in PC, LPC, PE, SM, acylcarnitines, and the Core-5 shared-metabolite module and repeatedly involved RDW-SD, GLU, GGT, ALP, TG, and HDL-C (Figures 6E and 6I and Figures 7C and 7E).

Regression decomposition of representative tandem association structures further characterized the components of the prioritized PFAS–omics feature–clinical phenotype–OC case status chains. Supported structures involving DONA, GenX, PFNA, and PFPeS repeatedly included LPC, PC, or acylcarnitine features and subsequently connected to RDW-SD. The tandem product of coefficients, the product of coefficients through the omics feature alone, and the product of coefficients through the clinical phenotype alone were displayed on the log-odds scale (Supplementary Figure S14).

Among the clinical phenotypes, RDW-SD repeatedly occurred in both lipid-related and shared metabolite-related association structures. This pattern suggested that a phenotype related to erythrocyte distribution heterogeneity may represent a recurrent observational connection node linking PFAS-associated molecular perturbations to the broader OC-related clinical phenotypic structure (Supplementary Tables S25–S26). Representative association structures further illustrated the organization of these multilayer connections. For example, DONA was associated not only with several specific OC-related acylcarnitine species but also with Core-5 shared metabolites, including corticosterone. The DONA–acylcarnitine–RDW-SD–OC association structure further illustrated how molecular features and clinical phenotype nodes may jointly contribute to exposure-related phenotypic patterns (Figure 7F).

Finally, the retained PFAS–omics–clinical evidence was integrated into a network. This network connected DONA, GenX, PFNA, and PFPeS with OC-related lipid classes, the Core-5 shared metabolites, and clinical phenotypes, thereby providing an evidence map of exposure-related molecular and phenotypic associations (Figure 7G and Supplementary Table S27).

Overall, the PFAS–omics–clinical integration analysis further supported multilayer observational links among the four-PFAS omics integration set, lipid remodeling, shared metabolites, and clinical phenotypic alterations. DONA and GenX, LPC/PC-related lipid modules, hippuric acid and elaidic acid, and RDW-SD were the most recurrent nodes in the integrated network.

### 3.7 Core-5 Shared Metabolites Provided Complementary Information beyond Conventional Clinical Features

The Core-5 feature set was evaluated in Datasets 1 and 2 using repeated stratified cross-validation. The modeling approaches included unpenalized logistic regression with prespecified predictors, LASSO logistic regression, and elastic net logistic regression; the penalized models were implemented using glmnet. Candidate feature sets comprised basic clinical variables, routine laboratory indicators, tumor markers, the Core-5 metabolites, and different combinations of these features (Figure 8A and Supplementary Table S28).

**Figure 8.**
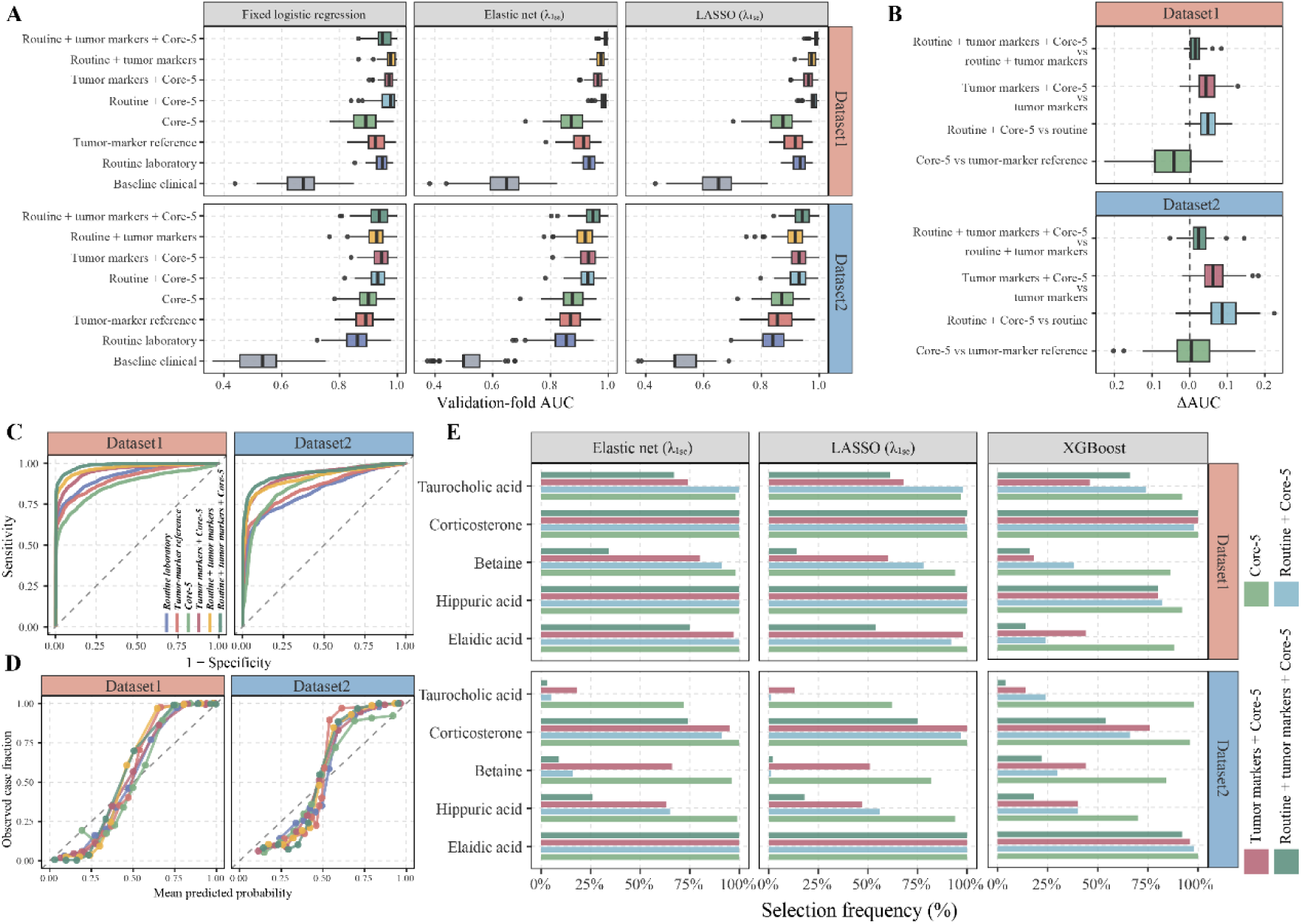
Core-5 shared metabolites provided complementary information beyond conventional clinical features and tumor markers. (A) Validation-fold AUC distributions across algorithms and feature sets in repeated stratified cross-validation in Datasets 1 and 2. Models included unpenalized logistic regression with prespecified predictors, elastic net logistic regression, and LASSO logistic regression; λ₁ₛₑ was used for elastic net and LASSO. Feature sets included basic clinical variables, routine laboratory measures, tumor marker reference features, the Core-5 shared metabolites, and their integrated combinations. Boxplots summarize the distribution of internal performance across validation folds and repeats. (B) Paired validation-fold ΔAUC distributions for four prespecified model comparisons: the Core-5-only model versus the tumor marker reference model, and the incremental addition of Core-5 to models based on routine laboratory measures, tumor markers, or their combination. ΔAUC was defined as the AUC of the Core-5-related model minus that of the corresponding reference model within the same validation fold; positive values indicate a higher validation-fold AUC for the Core-5-related model. Corresponding empirical 2.5th–97.5th percentile intervals are provided in Supplementary Table S29. (C) ROC curves based on participant-level out-of-fold predicted probabilities generated during repeated cross-validation. The x-axis represents 1 − specificity, the y-axis represents sensitivity, and the diagonal dashed line indicates no discrimination. (D) Internal calibration curves for models based on different feature sets, showing the relationship between the mean out-of-fold predicted probability and the observed case proportion within groups of predicted risk. (E) Selection frequencies of the Core-5 metabolites in elastic net, LASSO, and XGBoost models. For elastic net and LASSO, selection was defined as a nonzero coefficient in the λ₁ₛₑ model fitted within a training fold. Color indicates the different candidate feature sets containing the Core-5 metabolites.

In the primary glmnet LASSO analysis, adding the Core-5 metabolites to models based on routine clinical features provided partial incremental discrimination in both datasets. Similar improvements were observed when the Core-5 metabolites were added to the tumor marker reference models, providing descriptive internal evidence that the Core-5 feature set contained complementary information beyond conventional clinical predictors (Figure 8B and Supplementary Table S29).

Out-of-fold receiver operating characteristic (ROC) and calibration curves were used to summarize the internal performance of models based on different feature sets (Figures 8C–D). Overall, models incorporating the Core-5 metabolites showed higher AUC distributions than the basic clinical models, although calibration deviations remained within some ranges of predicted probability. Feature-selection frequencies varied by biospecimen matrix, modeling algorithm, and background predictor set; however, corticosterone consistently emerged as an important feature in both datasets (Figure 8E).

We further performed XGBoost sensitivity analyses to assess the robustness of these findings across a nonlinear modeling approach. The results were generally consistent with those of the primary analyses. After adding the Core-5 metabolites to the routine clinical feature models, the median paired ΔAUC values were 0.070 and 0.056 in Datasets 1 and 2, respectively. When the Core-5 metabolites were added to the tumor marker models, the corresponding median paired ΔAUC values were 0.055 and 0.026, respectively (Supplementary Figures S13B–D and Supplementary Tables S30–S31).

Collectively, this study generated three levels of observational evidence. First, plasma PFAS exposure biomarkers were associated with OC case status. Second, the four- PFAS omics integration set showed structured associations with metabolomic, lipidomic, and clinical phenotypes. Third, the Core-5 feature set provided complementary information related to OC case status under internal cross-validation.

## 4 Discussion

In this study, we integrated plasma PFAS exposure biomarkers with plasma metabolomics, plasma lipidomics, urine metabolomics, and clinical phenotype profiles to systematically characterize exposure-related molecular features in OC. This framework extended conventional exposure–disease analyses by linking environmental exposures to molecular alterations across multiple omics layers and biospecimen matrices.

A notable finding was that the burdens of alternative and emerging PFAS showed stronger associations with OC case status than several legacy PFAS categories. These associations were particularly prominent for DONA and GenX, two perfluoroether carboxylic acids (PFECAs) introduced as alternatives to PFOA (*36*, *37*). Following the phase-out and restriction of selected long-chain PFAS, such fluorinated alternatives have been developed and adopted; however, their population exposure profiles, toxicologic effects, toxicokinetics, and long-term health implications remain insufficiently characterized, particularly for most less-studied PFAS (*19*, *38*, *39*). Evidence regarding PFECAs in gynecologic malignancies is especially limited, despite their reported associations with lipid perturbations (*18*, *19*, *40*). In our analyses, DONA and GenX repeatedly emerged as highly connected exposure nodes across mixture models and omics association analyses, suggesting that their potential relevance to female reproductive health may have been underestimated.

These findings reinforce the importance of evaluating PFAS as a chemically diverse exposure class rather than treating them as a homogeneous group. Most previous population-based studies have focused on PFOA, PFOS, and a limited number of other legacy PFAS, whereas evidence for DONA, GenX, and related alternatives remains sparse (*21*, *37*). As alternative compounds increasingly replace selected long-chain PFAS, exposure assessment and health-risk evaluation should account for differences in chemical structure, toxicokinetics, biological activity, and mixture composition across individual PFAS (*38*, *39*, *41*).

The observed associations between PFAS and OC-related biological features are supported by converging evidence involving endocrine signaling, immune regulation, inflammation, hepatic function, and lipid metabolism (*16*, *28*). Previous studies have linked PFAS exposure to dyslipidemia, altered cholesterol homeostasis, immune dysfunction, changes in liver-related biomarkers, endocrine disruption, and potential carcinogenic effects (*13*, *28*). Many of these biological domains overlap with pathways involved in ovarian function and systemic responses to ovarian malignancy. Recent epidemiologic evidence has also suggested that prediagnostic concentrations of selected PFAS may be associated with subsequent OC risk (*18*). Building on this evidence, our study moved beyond the exposure–disease association alone by systematically characterizing exposure-related molecular phenotypes. The repeated prioritization of alternative PFAS and lipid-related molecular modules across multiple analytical layers provides a biologically interpretable basis for future prospective and mechanistic investigations.

The plasma metabolomic and lipidomic findings suggest that lipid metabolism may represent a major molecular interface linking PFAS exposure to OC-related systemic phenotypes (*23*, *25*, *28*). Untargeted plasma metabolomics revealed broad perturbations involving fatty acid metabolism, carboxylic acid metabolism, phosphatidylcholine metabolism, lipid transport, lipid binding, and fatty acid binding. Lipidomic profiling further resolved class-specific remodeling across LPC, LPE, PC, PE, SM, CerP, TG, DG, acylcarnitines, ceramides, and other lipid species. These patterns suggest that tumor-related metabolic remodeling and environmental chemical exposure may converge on biological processes involving membrane lipid composition, lipid transport, inflammatory signaling, oxidative stress, and energy metabolism (*23–26*). The PFAS–lipid association analyses further supported this interpretation. DONA and GenX showed the largest numbers of candidate bridging lipid associations, particularly involving LPC, PC, PE, SM, TG, DG, and acylcarnitines. These lipid classes participate in membrane structure, phospholipid turnover, lysophospholipid signaling, mitochondrial fatty acid transport, and neutral lipid storage (*25*). From an environmental molecular epidemiology perspective, these findings indicate that alternative PFAS may be associated with high-dimensional lipidomic variation that is not adequately captured by conventional circulating lipid measurements alone, consistent with recent experimental and population-based lipidomics studies (*28*, *40*, *42*).

Cross-matrix metabolite prioritization further strengthened the biological interpretability of the findings. Plasma and urine reflect complementary dimensions of systemic metabolism: plasma captures circulating metabolic status, molecular transport, inflammatory responses, and lipid remodeling, whereas urine reflects metabolite excretion, renal handling, host–microbial co-metabolism, and systemic metabolic turnover. Because metabolites often exhibit substantial matrix specificity, features consistently observed in both plasma and urine may represent more robust candidates for subsequent targeted validation.

Among the Core-5 metabolites, elaidic acid and hippuric acid appeared repeatedly in the PFAS–omics–clinical integration analyses. Elaidic acid may reflect alterations in fatty acid handling and lipid metabolic status, whereas hippuric acid is commonly linked to host–microbial co-metabolism and the metabolic processing of exogenous compounds (*43*, *44*). Betaine, corticosterone, and taurocholic acid further pointed to one-carbon metabolism, steroid-related signaling, and bile acid metabolism, respectively (*45–47*). These metabolic domains are relevant to both OC-related systemic physiology and environmental exposure biology (*21*, *25*). The recurrence of these metabolites across biospecimen matrices supports their prioritization as candidate shared cross-matrix metabolic features.

The integrated PFAS–omics–clinical network connected exposure biomarkers with candidate molecular features and measurable clinical phenotypes. DONA, GenX, LPC/PC-related lipid modules, hippuric acid, elaidic acid, and RDW-SD were among the most recurrent nodes. In particular, RDW-SD repeatedly occurred in both lipid- and shared metabolite-related association structures, suggesting that erythrocyte distribution heterogeneity may represent an observational connection between high-dimensional molecular variation and broader systemic phenotypic alterations in OC. Internal cross-validation analyses indicated that the Core-5 metabolites provided complementary information beyond basic clinical variables, routine laboratory indicators, and tumor markers. The incremental discriminatory patterns observed across LASSO, elastic net, and XGBoost models supported the internal stability of the Core- 5 feature set across different modeling strategies.

This study has several strengths. First, we simultaneously evaluated legacy, short-chain, long-chain, alternative, and emerging PFAS in human plasma, enabling systematic analyses at the levels of individual compounds, class-specific burdens, and exposure mixtures. To our knowledge, this is the first epidemiologic study to identify associations of DONA and GenX with OC-related lipidomic remodeling. Second, the integration of exposureomics, plasma metabolomics, plasma lipidomics, urine metabolomics, and clinical phenotypes enabled multilayer characterization of exposure-related molecular patterns. Third, the inclusion of both plasma and urine allowed the prioritization of shared cross-matrix metabolic features and reduced reliance on a single biospecimen matrix. Fourth, the PFAS–omics–clinical network provided a transparent and systematic strategy for organizing high-dimensional associations into interpretable exposure-related molecular and phenotypic structures. Fifth, repeated internal cross - validation and sensitivity analyses using different algorithms provided additional evidence regarding the stability of the Core-5 feature set.

Several limitations should also be considered. First, the case–control design precluded determination of the temporal sequence between PFAS exposure, molecular alterations, and OC. Second, molecular and clinical measurements were obtained at a single time point, limiting inference regarding long-term exposure patterns and dynamic metabolic changes. Third, the product-of-coefficients and network analyses were intended solely for observational prioritization and should not be interpreted as causal pathway analyses. Fourth, owing to the sample size and lack of an external population, the Core-5 feature set has not yet been independently validated. Its reproducibility and generalizability should therefore be assessed in independent populations, prospectively collected biospecimens, and standardized targeted analytical platforms.

Overall, this study linked plasma PFAS exposure biomarkers, particularly the alternative and emerging PFAS DONA and GenX, to OC-related lipidomic, metabolomic, and clinical phenotypic alterations. The findings highlighted PFECAs as priority exposures in relation to OC and identified lipid metabolism and shared cross - matrix small-molecule features as key molecular domains connecting PFAS exposure with systemic phenotypic variation in OC.

From a broader environmental health perspective, these results underscore the need to expand PFAS biomonitoring and health assessment beyond well-characterized legacy compounds. The stronger and more extensive molecular associations observed for selected alternative PFAS suggest that chemical substitution should not be assumed to reduce health hazards without adequate epidemiological and toxicological evaluation. Incorporating alternative and emerging PFAS, mixture-aware analytical methods, and multi-matrix molecular profiling into future studies may improve the early identification of biologically responsive exposure patterns and provide a more comprehensive evidence base for chemical substitution policies, exposure reduction strategies, and environmental health risk assessment.

## Abbreviations

AcCa, acylcarnitine; AFP, alpha-fetoprotein; ALB, albumin;

ALP, alkaline phosphatase; ALT, alanine aminotransferase;

AST, aspartate aminotransferase; AUC, area under the curve;

BKMR, Bayesian kernel machine regression; BMI, body mass index;

BH, Benjamini–Hochberg; CA125, carbohydrate antigen 125;

CA15-3, carbohydrate antigen 15-3; CEA, carcinoembryonic antigen; Cer, ceramide;

CerP, ceramide-1-phosphate; ChE, cholesteryl ester;

CI, confidence interval;

Core-5, a five-metabolite cross-matrix feature set comprising elaidic acid, hippuric acid, betaine, corticosterone, and taurocholic acid;

DG, diacylglycerol; EOS, eosinophils; FA, fatty acid;

FDR, false discovery rate;

FIGO, International Federation of Gynecology and Obstetrics; GGT, gamma-glutamyl transferase;

GLOB, globulin;

GO, Gene Ontology;

GLU, glucose;

gWQS, generalized weighted quantile sum regression;

HDL-C, high-density lipoprotein cholesterol; HGB, hemoglobin;

KEGG, Kyoto Encyclopedia of Genes and Genomes; LDL-C, low-density lipoprotein cholesterol;

LPE, lysophosphatidylethanolamine; LPI, lysophosphatidylinositol;

LASSO, least absolute shrinkage and selection operator; LPC, lysophosphatidylcholine;

LYM, lymphocytes;

MCH, mean corpuscular hemoglobin; MCV, mean corpuscular volume;

MeFOSAA, 2-(N-methyl-perfluorooctane sulfonamido)acetic acid; MG, monoacylglycerol;

MRM, multiple reaction monitoring; NEU, neutrophils;

OAHFA, O-acyl-ω-hydroxy fatty acid; OC, ovarian cancer;

OR, odds ratio;

PA, phosphatidic acid; PC, phosphatidylcholine;

PCA, principal component analysis; PE, phosphatidylethanolamine;

PFAS, per- and polyfluoroalkyl substances;

PFAA, primary fatty acid amide (LipidSearch nomenclature); PFECAs, perfluoroether carboxylic acids;

PFCAs, perfluoroalkyl carboxylic acids; PFSAs, perfluoroalkyl sulfonic acids; PFOA, perfluorooctanoic acid;

PFOS, perfluorooctanesulfonic acid; PIP, posterior inclusion probability;

PLS-DA, partial least squares discriminant analysis; qgcomp, quantile g-computation;

QC, quality control;

RDW-SD, red blood cell distribution width–standard deviation; ROC, receiver operating characteristic;

RSD, relative standard deviation; SM, sphingomyelin;

TG, triacylglycerol (lipidomic class); TG, triglycerides (clinical measure); TP, total protein;

UPLC–MS/MS, ultra-performance liquid chromatography–tandem mass spectrometry; VIP, variable importance in projection;

XGBoost, extreme gradient boosting.

## Competing interests

All authors declare that they have no conflict of interest.

## Data Availability

The de-identified data that support the findings of this study are available from the corresponding author upon reasonable request, subject to applicable ethical and institutional requirements. Data that could potentially compromise participant privacy are not publicly available. All other data supporting the findings of this study are included in the manuscript and its Supplementary Information.

## Acknowledgements

This work was supported by the National Natural Science Foundation of China (82302974), Natural Science Foundation of Jiangsu Province (BK20241102), Jiangsu Provincial Scientific Research and Health Project (ZD2022005), Yishan Research Project of Jiangsu Cancer Hospital (YSPY202401), Qunfeng Project of Jiangsu Cancer Hospital (DFXK202504), Research Project of Jiangsu Cancer Hospital (RCQY202402), and Postgraduate Research & Practice Innovation Program of Jiangsu Province (KYCX24_0489).

## Notes

### Competing Interest Statement

The authors have declared no competing interest.

### Author Declarations

The study protocol was approved by the Ethics Committee of Jiangsu Cancer Hospital (KY-2024-139). All participants provided written informed consent.

