## Supplementary Information for "Alternative PFAS Are Associated with Ovarian Cancer–Related Lipidomic Remodeling and Cross-Matrix Metabolic Signatures"

### Supplementary Figures

Supplementary Figure S1. Clinical characteristics and sensitivity analyses in Dataset 1

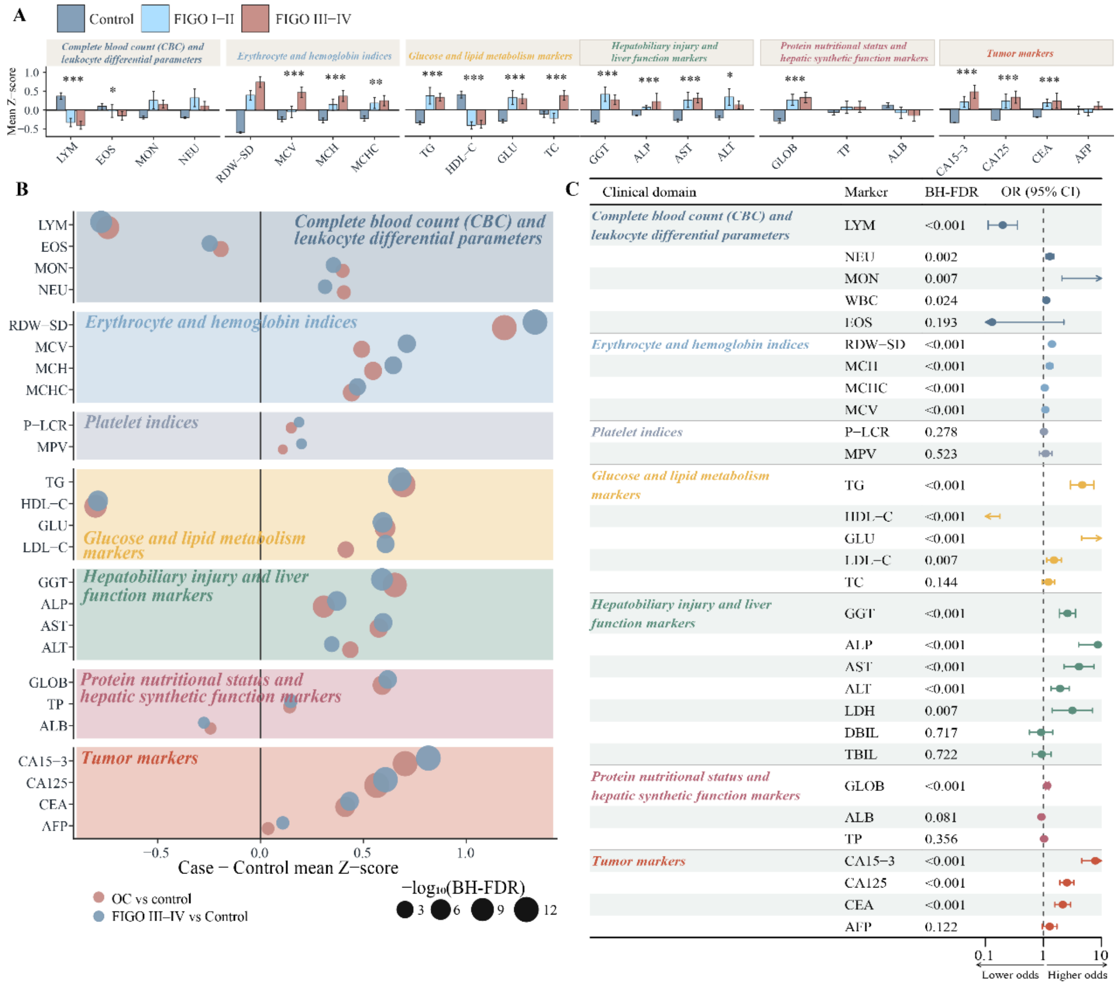

(A) Standardized mean values of clinical indicators among controls, patients with FIGO stage I–II ovarian cancer (OC), and patients with FIGO stage III–IV OC in Dataset 1. Bars represent group-specific mean z scores, and error bars indicate standard errors. Statistical tests were performed using the original-scale values. Asterisks indicate statistical significance in the overall three-group comparison after Benjamini–Hochberg false discovery rate correction: * BH-FDR < 0.05,  BH-FDR < 0.01, and * BH-FDR < 0.001.

(B) Cleveland dot plot showing the overall case–control differences and the differences between patients with advanced-stage OC and controls. Point size represents $-\log_{10}$(BH-FDR).

(C) Multivariable logistic regression associations between clinical indicators and OC case status, adjusted for age, body mass index (BMI), and hypertension.

Supplementary Figure S2. PCA and PLS-DA score plots of the plasma metabolome

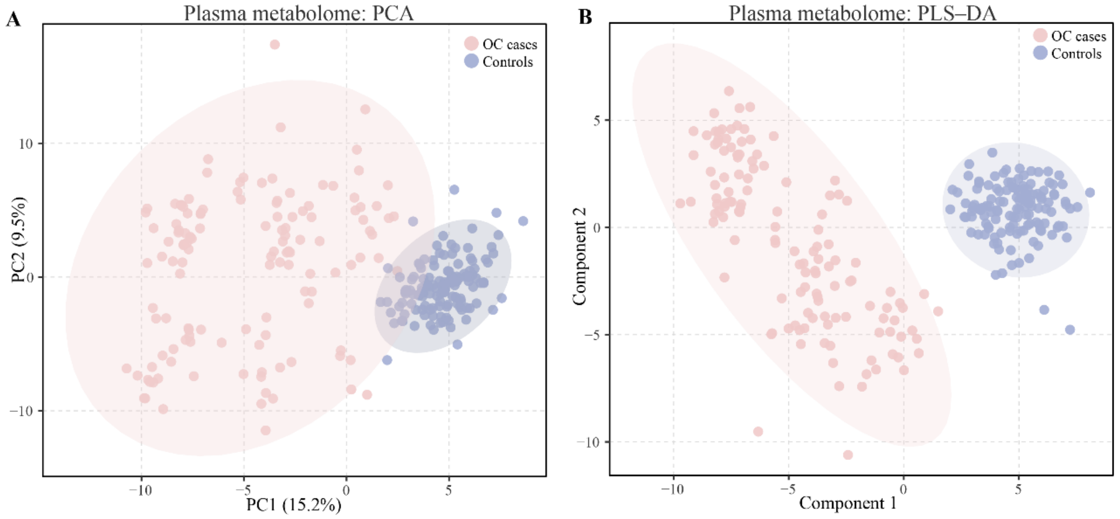

(A) Unsupervised principal component analysis (PCA) score plot of the plasma metabolome following $\log_2$ transformation and feature preprocessing.

(B) Supervised partial least-squares discriminant analysis (PLS-DA) score plot of the plasma small-molecule metabolome following $\log_2$ transformation and feature preprocessing.

Supplementary Figure S3. PFAS exposure structure and sensitivity analyses

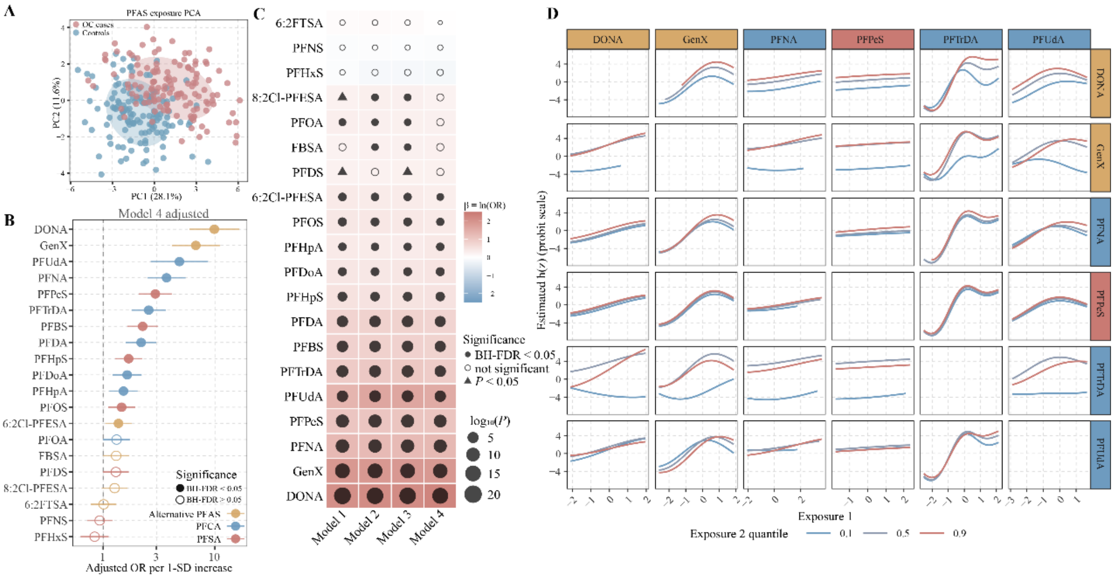

(A) PCA score plot of plasma per- and polyfluoroalkyl substance (PFAS) exposure profiles in Dataset 1.

(B) Sensitivity-analysis forest plot of the associations between individual PFAS and OC case status. Models were adjusted for age, BMI, and hypertension. Points represent adjusted odds ratios (ORs), horizontal lines represent 95% confidence intervals (CIs), and the vertical dashed line indicates OR = 1. ORs correspond to a 1-standard-deviation increase in $\log_2$-transformed PFAS concentrations and are displayed on a logarithmic scale. Point colors indicate PFAS classes, and filled versus open symbols indicate statistical support after BH-FDR correction.

(C) Heatmap of the associations between individual PFAS and OC case status across Models 1–4. Tile colors represent logistic regression coefficients (β), equivalent to ln(OR), and point sizes represent $-\log_{10}(P)$. Model 1 was unadjusted; Model 2 was adjusted for age; Model 3 was adjusted for age and BMI; and Model 4 was additionally adjusted for hypertension.

(D) Conditional bivariate exposure–response functions from Bayesian kernel machine regression (BKMR) for the six disease-prioritized PFAS.

Supplementary Figure S4. Scatterplots of representative PFAS–plasma metabolite associations

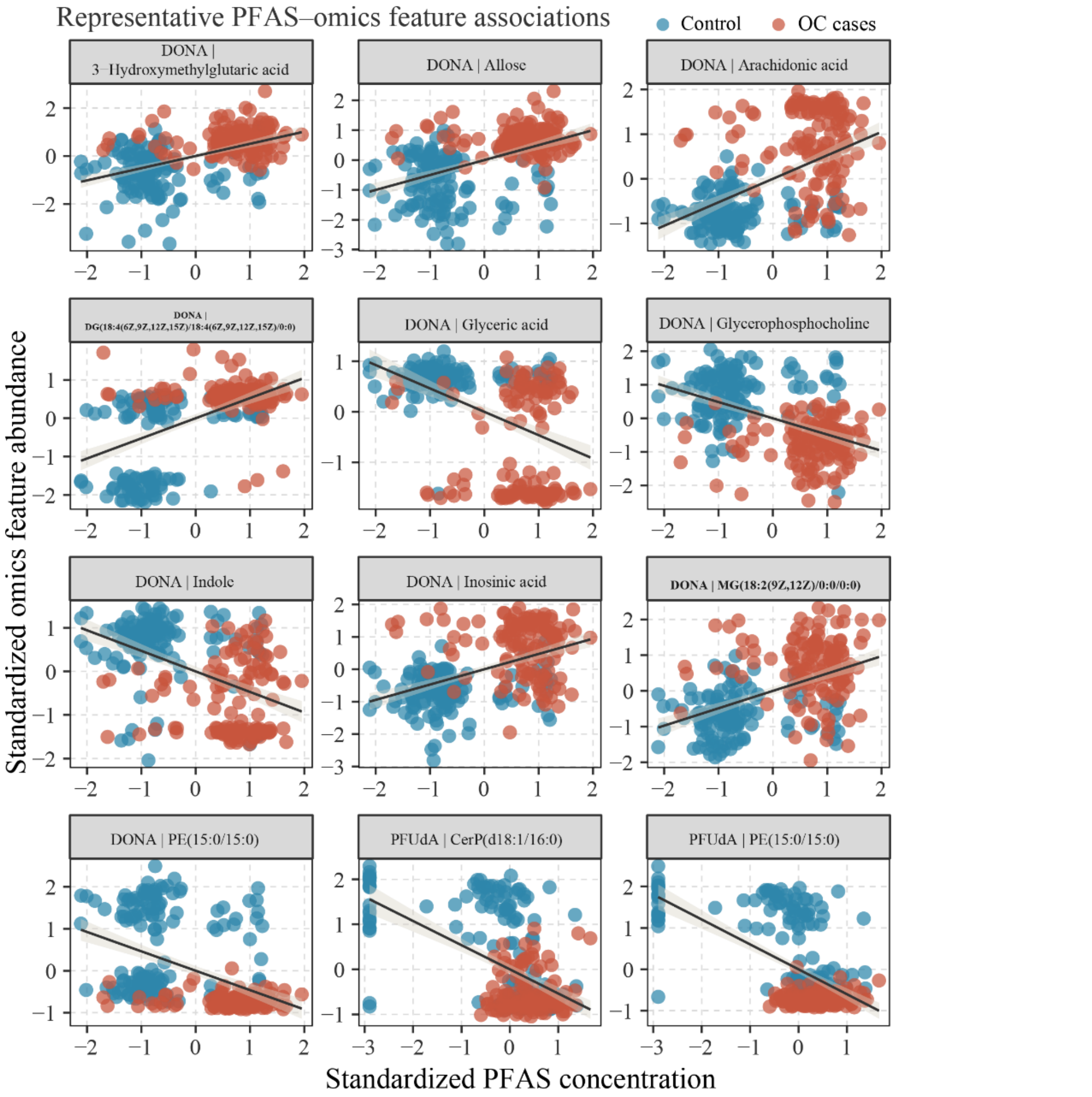

Each facet shows a representative association between a PFAS and a plasma metabolite. The x-axis represents $\log_2$-transformed and z-standardized PFAS concentrations, and the y-axis represents $\log_2$-transformed and z-standardized omics-feature abundances. Each point represents one participant, with colors indicating OC cases and controls. Solid lines represent covariate-adjusted fitted associations, and shaded areas indicate 95% confidence bands. The displayed associations were prioritized according to the absolute values of the standardized regression coefficients. Models were adjusted for age, BMI, and hypertension.

Supplementary Figure S5. Associations between plasma lipids and OC case status

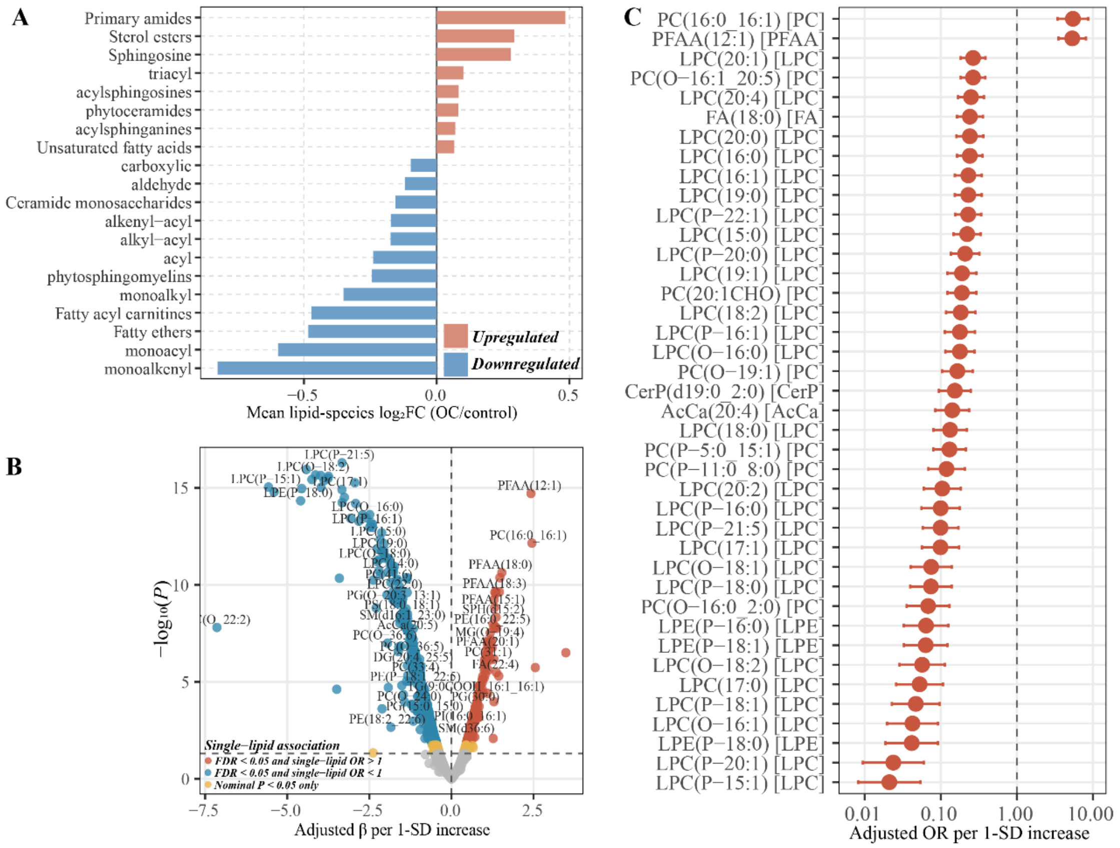

(A) Mean case–control $\log_2$ fold changes across annotated lipid subclasses. For each lipid species, the $\log_2$ fold change ($\log_2$FC) was defined as the mean $\log_2$ abundance in OC cases minus the mean $\log_2$ abundance in controls. Lipid-specific $\log_2$FC values were subsequently averaged within the corresponding lipid subclasses. Positive values indicate higher abundances in cases, whereas negative values indicate lower abundances in cases.

(B) Expanded volcano plot of the associations between individual plasma lipids and OC case status. Multivariable logistic regression models were adjusted for age and BMI. The x-axis represents the adjusted logistic regression coefficient (β) corresponding to a 1-standard-deviation increase in $\log_2$-transformed lipid abundance, and the y-axis represents $-\log_{10}$(P). Colors indicate mutually exclusive evidence categories defined according to BH-FDR, nominal P values, and the direction of association.

(C) Forest plot of individual lipid associations. Points represent ORs adjusted for age and BMI, horizontal lines represent 95% CIs, and the vertical dashed line indicates OR = 1. ORs correspond to a 1-standard-deviation increase in $\log_2$-transformed lipid abundance and are displayed on a logarithmic scale.

Supplementary Figure S6. Scatterplots of representative PFAS–candidate bridging lipid associations

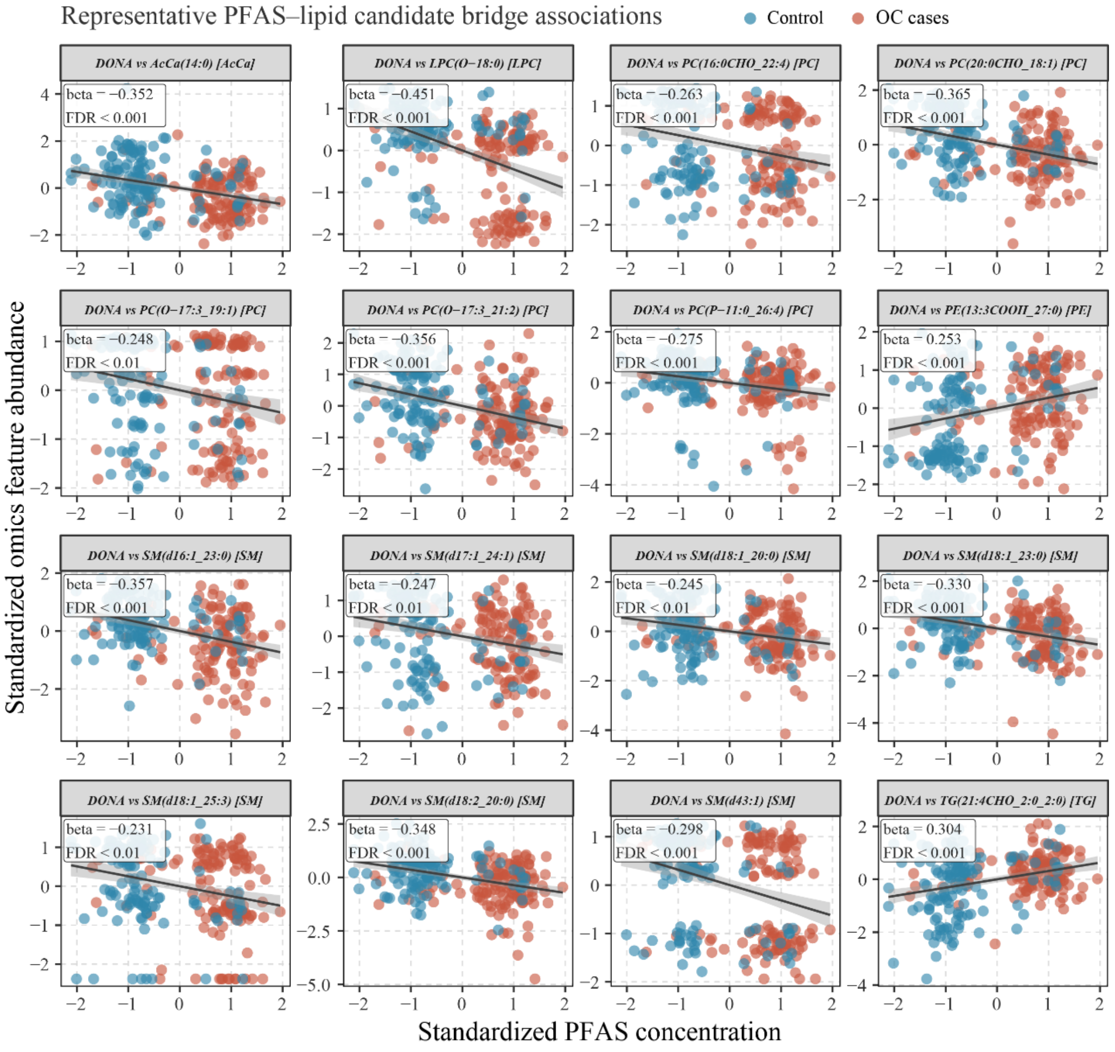

Each facet shows a representative covariate-adjusted association between a PFAS and a candidate bridging lipid. The x-axis represents $log_2$-transformed and z-standardized PFAS concentrations, and the y-axis represents $log_2$-transformed and z-standardized omics-feature abundances. Each point represents one participant, with colors indicating OC cases and controls. Solid lines represent covariate-adjusted fitted associations, and shaded areas indicate 95% confidence bands. The displayed associations were prioritized according to the absolute values of the standardized regression coefficients. Models were adjusted for age, BMI, and hypertension.

Supplementary Figure S7. Heatmaps of associations between candidate plasma omics features and clinical indicators

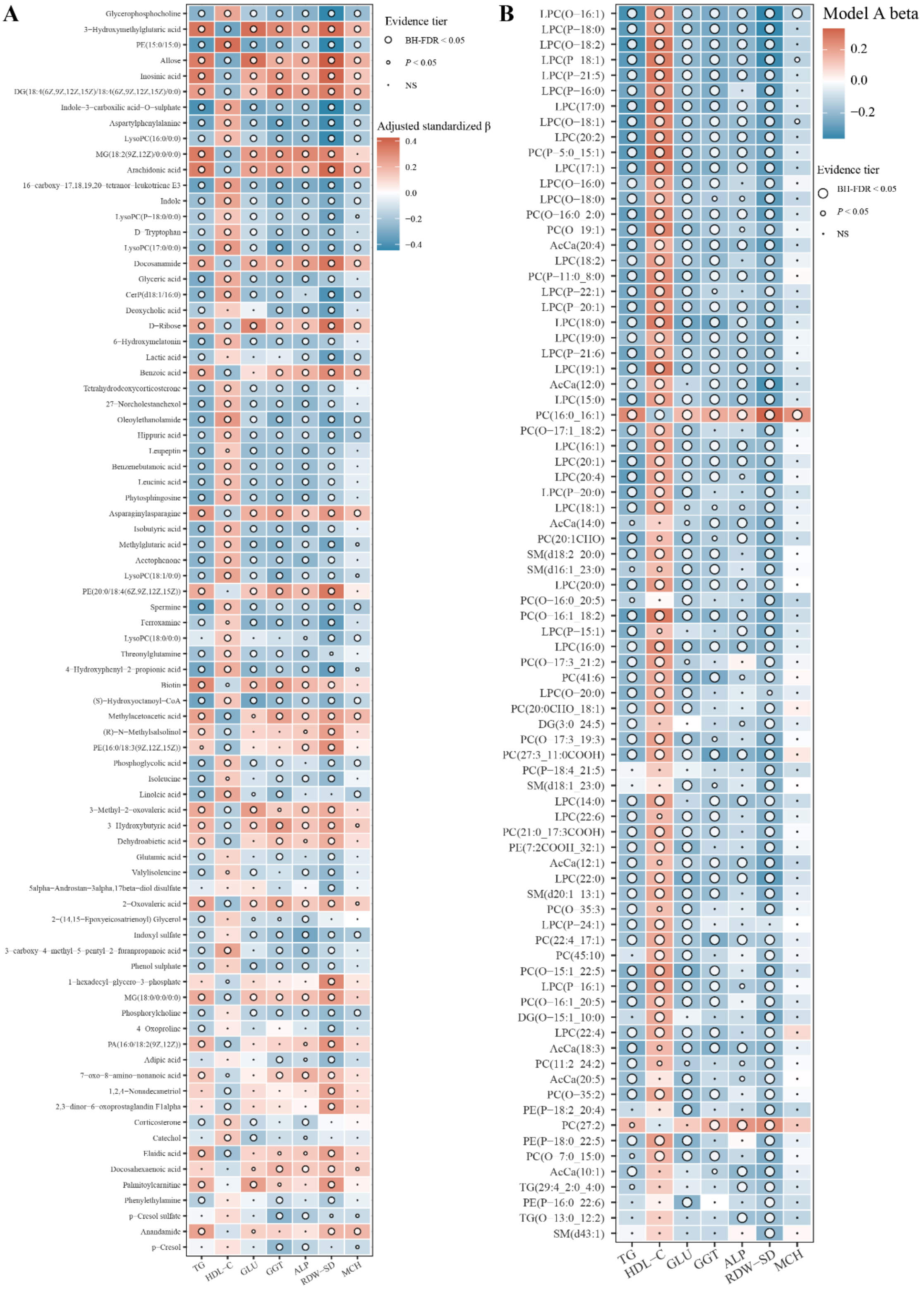

(A) Covariate-adjusted associations between candidate plasma metabolites and clinical indicators. (B) Covariate-adjusted associations between candidate plasma lipids and clinical indicators. Tile colors represent adjusted standardized regression coefficients (β). The coefficient β denotes the change in the corresponding clinical indicator, expressed in standard-deviation units, per 1-standard-deviation increase in the omics feature. Plasma metabolites were $log_2$-transformed and z-standardized. Lipids underwent half-minimum-value imputation, followed by $log_2$ transformation and z-standardization. Triglycerides (TG), glucose (GLU), γ-glutamyl transferase (GGT), and alkaline phosphatase (ALP) were $log_2$-transformed and z-standardized, whereas high-density lipoprotein cholesterol (HDL-C), red blood cell distribution width–standard deviation (RDW-SD), and mean corpuscular hemoglobin (MCH) were z-standardized on their original scales.

Supplementary Figure S8. Regression-based decomposition of prioritized PFAS–lipid–OC case-status association structures

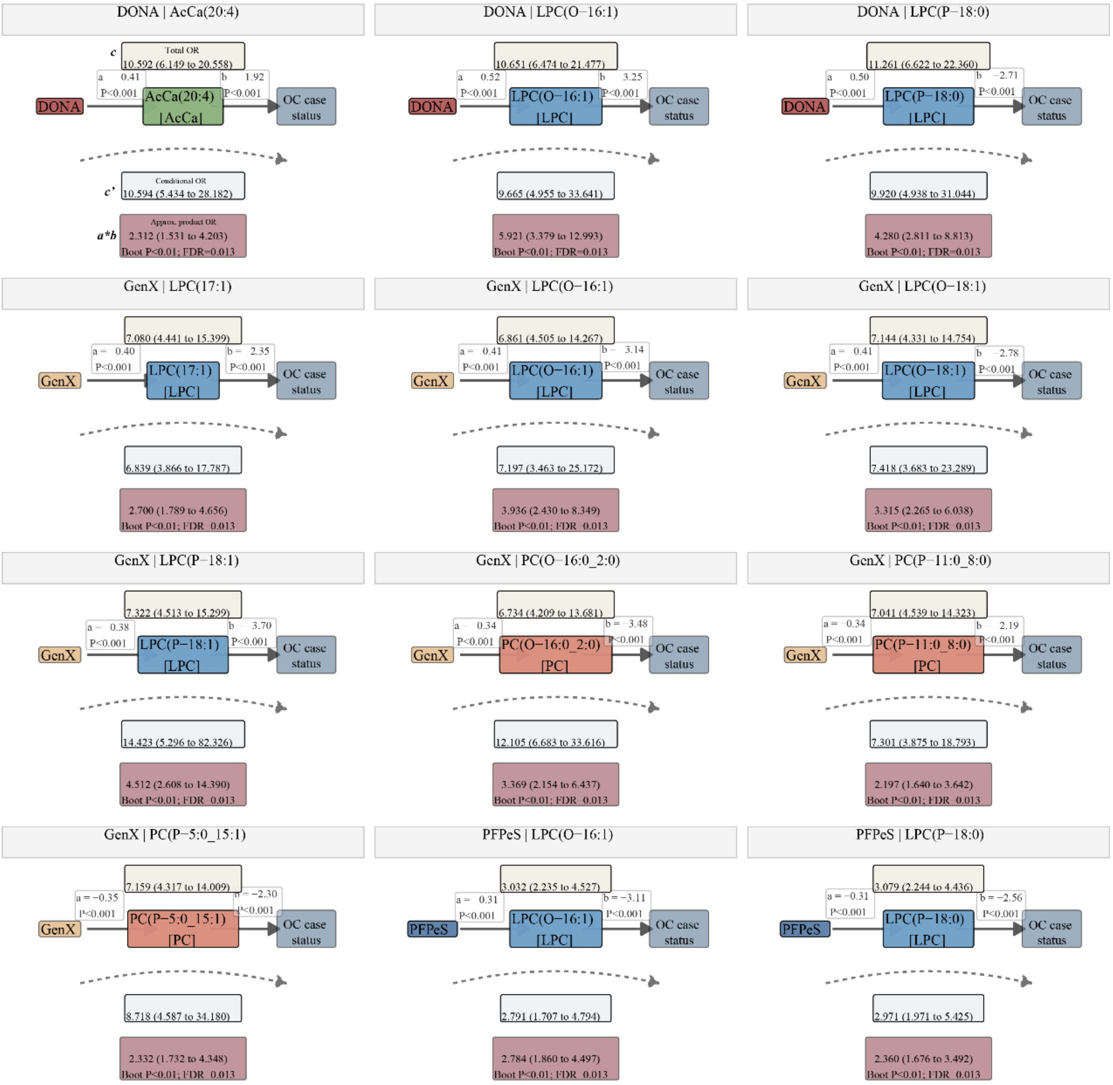

The figure presents representative bootstrap-supported PFAS–candidate bridging lipid–OC case-status association structures. The coefficient a represents the change in lipid abundance, expressed in standard-deviation units, per 1-standard-deviation increase in $\log_2$-transformed and z-standardized PFAS concentration. The coefficient b represents the change in the log odds of OC case status per 1-standard-deviation increase in lipid abundance after adjustment for the PFAS and covariates. The coefficient c represents the total association between the PFAS and OC case status, whereas c′ represents the conditional association between the PFAS and OC case status after inclusion of the candidate lipid. The coefficient-product estimate was defined as a × b, and all case-status-related estimates are presented on the log-odds scale. Values in parentheses indicate 95% CIs. Models were adjusted for age, BMI, and hypertension. Uncertainty in the coefficient-product estimates was evaluated using 300 participant-level nonparametric bootstrap resamples, and percentile-based 95% CIs were calculated. Bootstrap P values were corrected using the BH method across all candidate PFAS–lipid pairs included in the resampling analysis.

Supplementary Figure S9. Clinical characteristics and sensitivity analyses in Dataset 2

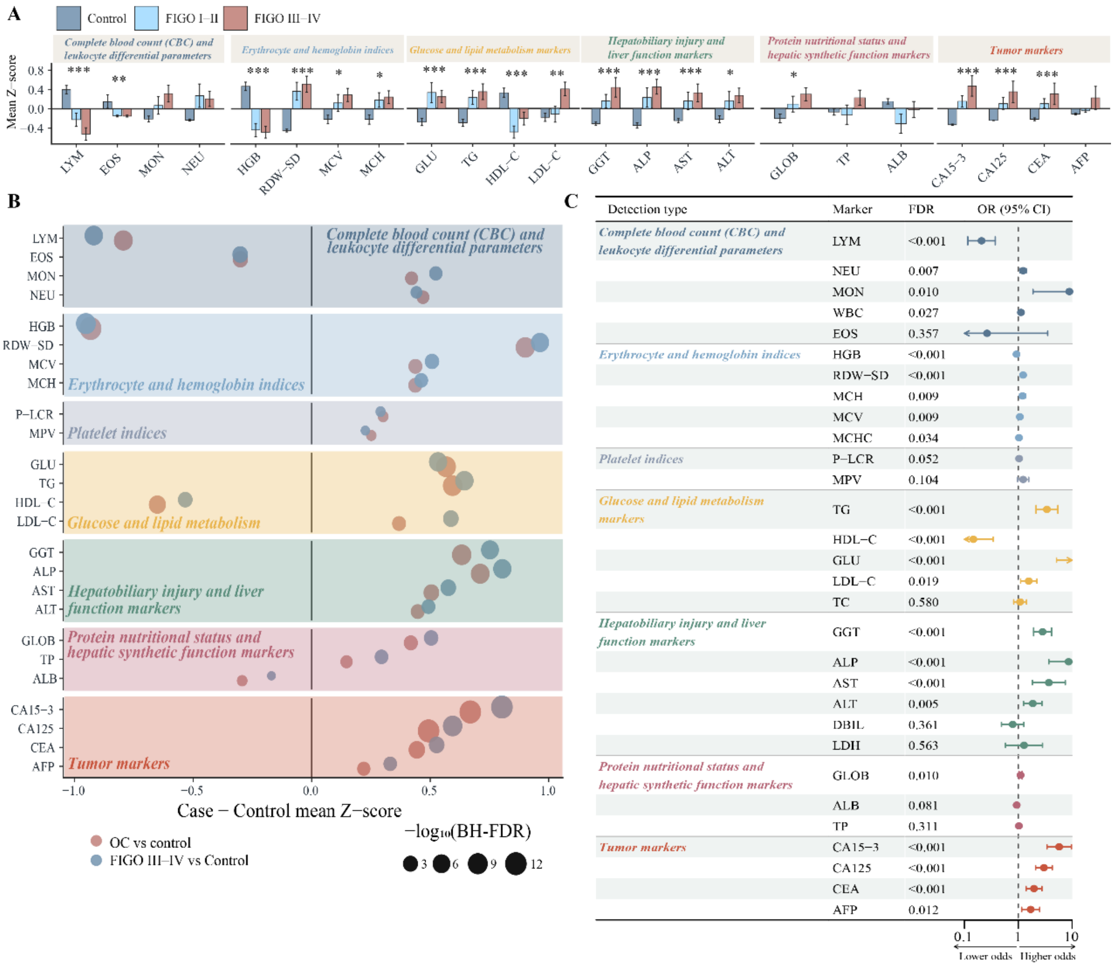

(A) Standardized mean values of clinical indicators among controls, patients with FIGO stage I–II OC, and patients with FIGO stage III–IV OC in Dataset 2. Bars represent group-specific mean z scores, and error bars indicate standard errors. Statistical tests were performed using the original-scale values. Asterisks indicate statistical significance in the overall three-group comparison after Benjamini–Hochberg false discovery rate correction: * BH-FDR < 0.05, BH-FDR < 0.01, and * BH-FDR < 0.001.

(B) Cleveland dot plot showing the overall case–control differences and the differences between patients with advanced-stage OC and controls. Point size represents −$\log_{10}$(BH-FDR).

(C) Multivariable logistic regression associations between clinical indicators and OC case status, adjusted for age, BMI, and hypertension.

Supplementary Figure S10. PCA, PLS-DA, and KEGG enrichment analyses of the urinary metabolome

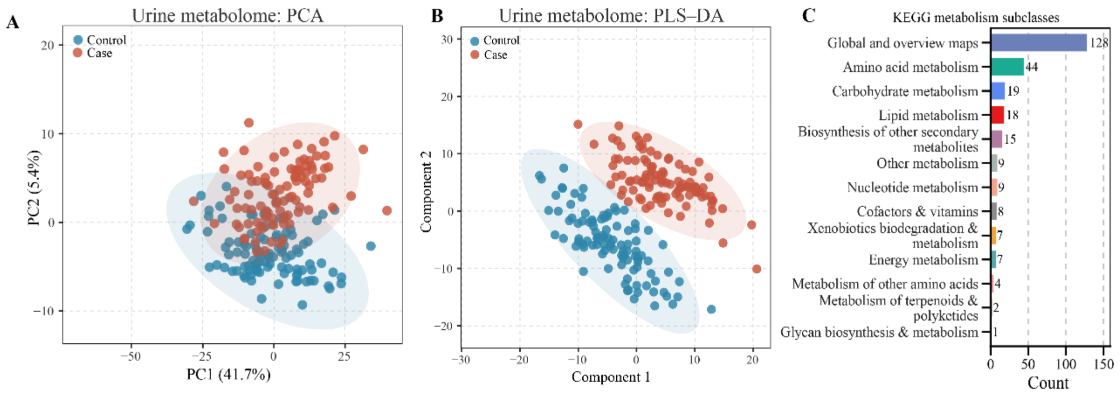

(A–B) PCA (A) and PLS-DA (B) score plots of the urinary metabolome following creatinine normalization, $\log_2$ transformation, and z-standardization.

(C) Counts of detected and differential urinary metabolites across Kyoto Encyclopedia of Genes and Genomes (KEGG) metabolic subclasses.

Supplementary Figure S11. Selection stability of shared plasma–urine metabolites in repeated LASSO analyses

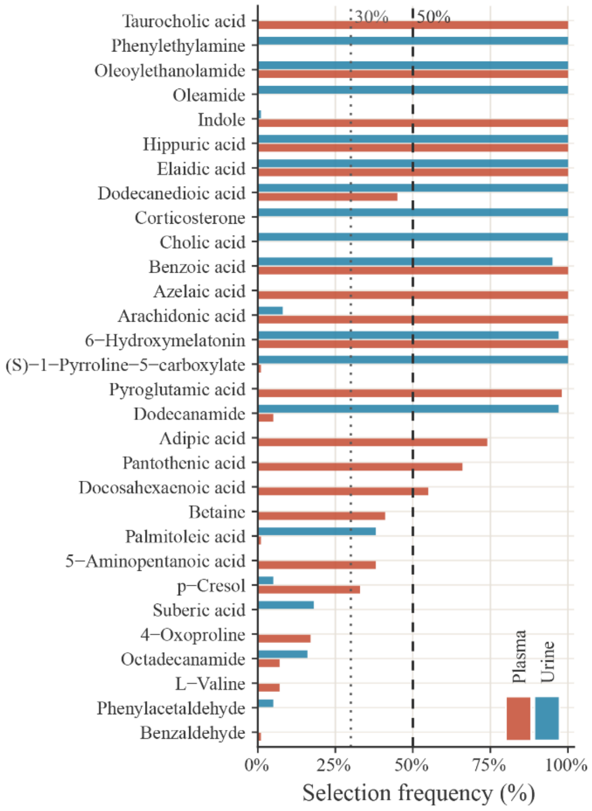

Repeated stratified least absolute shrinkage and selection operator (LASSO) logistic regression analyses were performed 100 times separately in the plasma and urine datasets using cross-matrix-matched metabolites. The primary analysis used the cross-validation-selected λ1se. A metabolite was considered selected when it had a nonzero coefficient in the λ1se model for the corresponding repetition. Selection frequency was defined as the proportion of successful repeated model fits in which the metabolite was selected among all successful fits in the corresponding biological matrix. Shared metabolites were included as penalized variables, whereas age, BMI, and hypertension were included as unpenalized covariates. Plasma metabolites were analyzed after $\log_2$ transformation and z-standardization. Urinary metabolites were analyzed after urinary creatinine normalization, $\log_2$ transformation, and z-standardization. The dotted and dashed lines denote the 30% suggestive-stability and 50% robust-stability thresholds, respectively. Selection frequencies from 30% to <50% were classified as suggestive support, whereas frequencies ≥50% were classified as robust support. These repeated LASSO analyses were used to assess candidate-feature stability and support cross-matrix prioritization.

Supplementary Figure S12. Prioritized PFAS–Core-5–OC case-status association structures

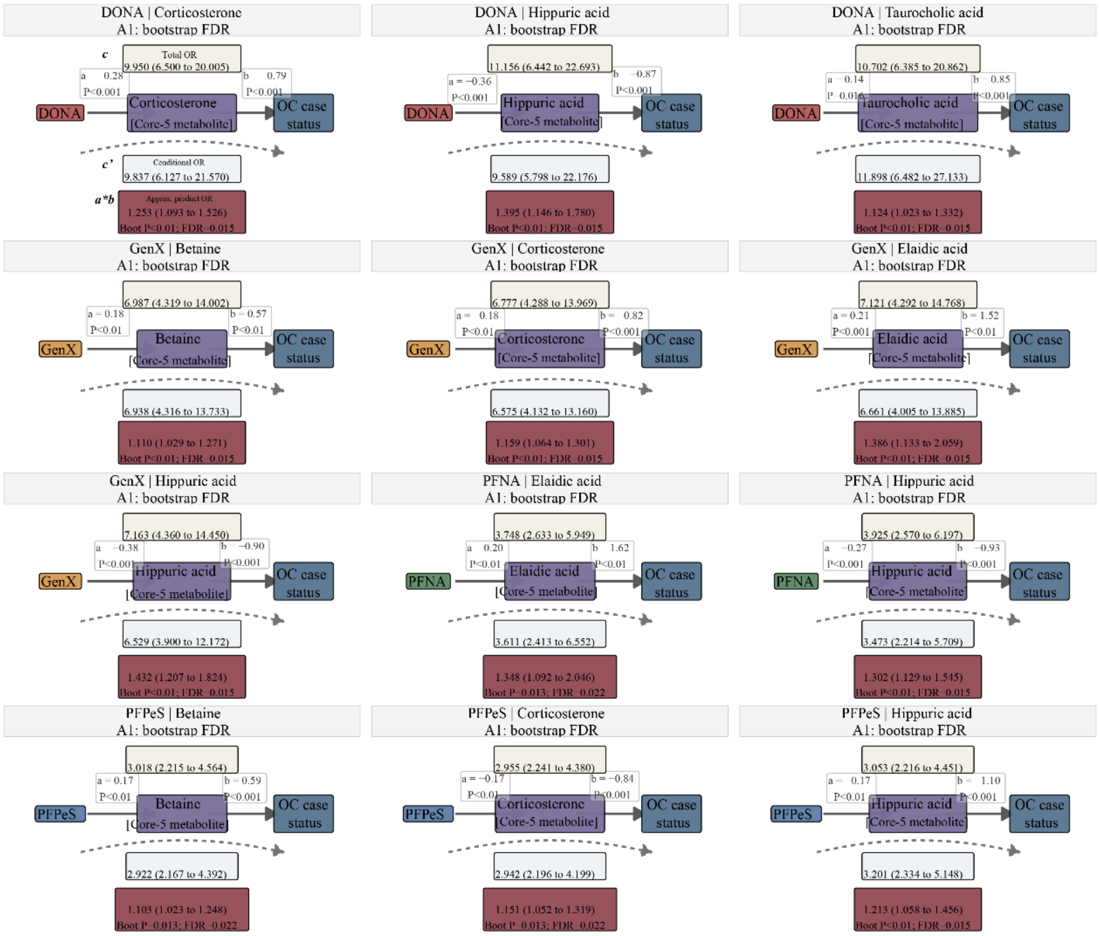

The figure presents representative bootstrap-supported PFAS–Core-5 shared metabolite–OC case-status association structures.

Supplementary Figure S13. Decision-curve and XGBoost sensitivity analyses for internally validated molecular-feature models

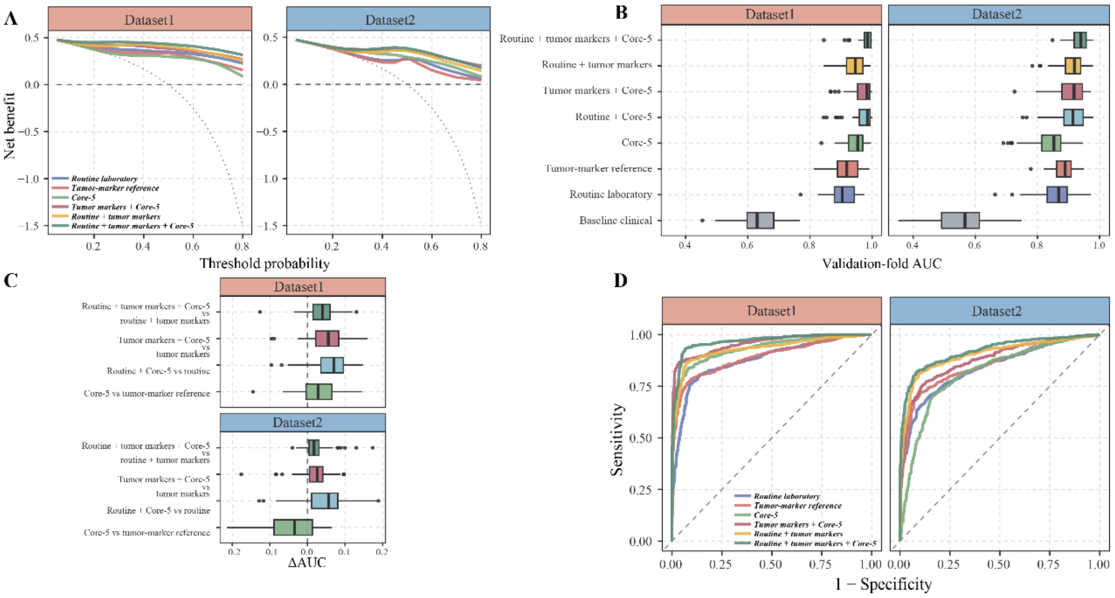

(A) Decision-curve sensitivity analysis for the primary glmnet LASSO 1-SE algorithm. Dashed and dotted lines indicate the treat-none and treat-all strategies, respectively.

(B) Cross-validated discrimination performance in the XGBoost sensitivity analysis using 5-fold cross-validation repeated 10 times.

(C) Increments in the area under the receiver operating characteristic curve (AUC) after adding the Core-5 shared metabolites to the XGBoost models.

(D) Summary out-of-fold receiver operating characteristic (ROC) curves from the XGBoost sensitivity analysis.

Supplementary Figure S14. Regression-based decomposition of prioritized serial PFAS–omics-feature–clinical-phenotype–OC case-status association structures

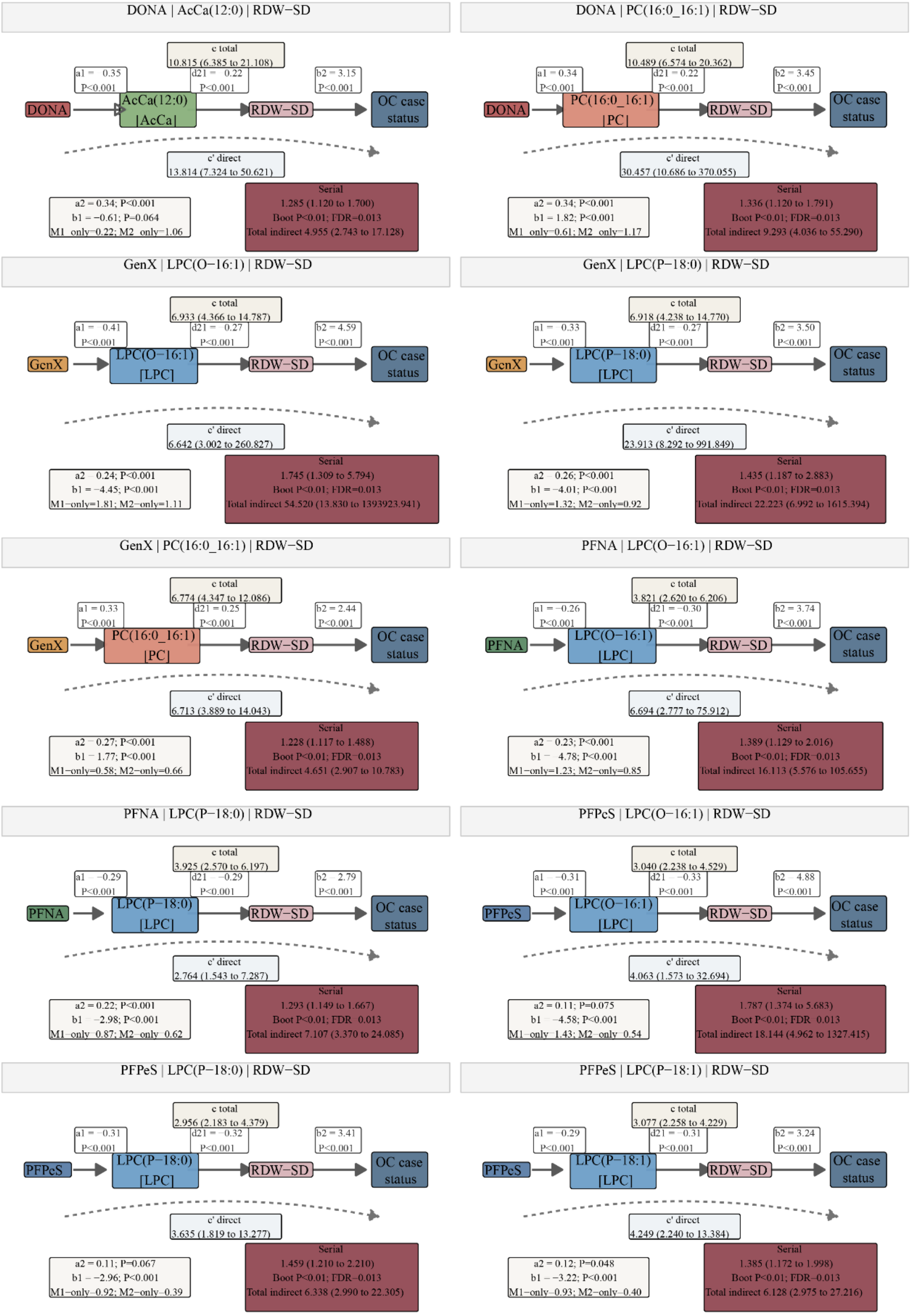
DONA | AcCa(12:0) | RDW−SD
DONA | PC(16:0_16:1) | RDW−SD
GenX | LPC(O−16:1) | RDW−SD
GenX | LPC(P−18:0) | RDW−SD
GenX | PC(16:0_16:1) | RDW−SD
PFNA | LPC(O−16:1) | RDW−SD
PFNA | LPC(P−18:0) | RDW−SD
PFPeS | LPC(O−16:1) | RDW−SD
PFPeS | LPC(P−18:0) | RDW−SD
PFPeS | LPC(P−18:1) | RDW−SD
OC case status

The figure presents representative bootstrap-supported serial PFAS–candidate omics feature–clinical phenotype–OC case-status association structures using the conventional a1, d21, b2, a2, b1, c, and c′ path notation. The coefficient a1 represents the change in the abundance of the first omics feature (M1), expressed in standard-deviation units, per 1-standard-deviation increase in $log_2$-transformed and standardized PFAS concentration. The coefficient d21 represents the change in the clinical phenotype (M2), expressed in standard-deviation units, per 1-standard-deviation increase in M1 after adjustment for the PFAS and covariates. The coefficient b2 represents the change in the log odds of OC case status per 1-standard-deviation increase in M2 after adjustment for the PFAS, M1, and covariates. The coefficient a2 represents the conditional association between the PFAS and M2, whereas b1 represents the logistic regression coefficient for the association between M1 and OC case status after adjustment for the PFAS, M2, and covariates. The coefficient c represents the total association between the PFAS and OC case status, whereas c′ represents the conditional association between the PFAS and OC case status after simultaneous inclusion of M1 and M2. The serial coefficient product was defined as a1d21b2. The coefficient products involving M1 alone and M2 alone were defined as a1b1 and a2b2, respectively, and the combined coefficient-product estimate was defined as the sum of these three products.

Uncertainty in the coefficient-product estimates was evaluated using 300 participant-level nonparametric bootstrap resamples, and horizontal intervals represent percentile-based 95% CIs. Bootstrap P values were corrected using the BH method within the prespecified multiple-testing family of serial association chains. Models were adjusted for age, BMI, and hypertension.

### Supplementary Materials

#### Table of Contents

*The supplementary tables are organized by dataset and analysis module for this case–control study. dataset1 denotes the plasma subset, and dataset2 denotes the urine subset.*

##### Part 1. Clinical and laboratory characteristics in dataset1

Table S1. Clinical and laboratory characteristics by case–control status in dataset1

Table S2. Clinical and laboratory characteristics of controls and ovarian cancer cases by FIGO stage group in dataset1

Table S3. Clinical and laboratory characteristics by FIGO stage group among ovarian cancer cases in dataset1

##### Part 2. Clinical and laboratory characteristics in dataset2

Table S4. Clinical and laboratory characteristics by case–control status in dataset2

Table S5. Clinical and laboratory characteristics of controls and ovarian cancer cases by FIGO stage group in dataset2

Table S6. Clinical and laboratory characteristics by FIGO stage group among ovarian cancer cases in dataset2

##### Part 3. Clinical laboratory marker models for ovarian cancer case status

Table S7. Logistic regression adjustment models for clinical laboratory marker associations with ovarian cancer case status

Table S8. Associations of clinical laboratory markers with ovarian cancer case status in dataset1

Table S9. Associations of clinical laboratory markers with ovarian cancer case status in dataset2

##### Part 4. PFAS exposure definitions and associations with ovarian cancer case status

Table S10. PFAS mixture set definitions

Table S11. Associations between PFAS group burdens and ovarian cancer case status

Table S12. Associations between individual PFAS and ovarian cancer case status

Table S13. Summary of individual PFAS associations by PFAS class in Model 4

#### Part 5. PFAS mixture associations with ovarian cancer case status

Table S14. Cross-method priority components within the disease-priority PFAS mixture for ovarian cancer case status

Table S15. PFAS mixture associations with ovarian cancer case status in qgcomp and gWQS models

Table S16. Component weights in the all-individual PFAS mixture for ovarian cancer case status

#### Part 6. PFAS-lipid association analyses in dataset1

Table S17. Associations of individual PFAS with ovarian cancer case status and lipid-association burden in dataset1

Table S18. Lipid-class summary of associations for disease-priority PFAS in dataset1

Table S19. Bridge-candidate counts by disease-priority PFAS and lipid class in dataset1

Table S20. Median covariate-adjusted PFAS-lipid association beta by disease-priority PFAS and lipid class in dataset1

#### Part 7. Mediation and integrated network analyses in dataset1

Table S21. Shared small-molecule mediation summary by disease-priority PFAS in dataset1

Table S22. Shared small-molecule mediation summary by mediator in dataset1

Table S23. Supported PFAS-shared metabolite-ovarian cancer mediation candidates in dataset1

Table S24. Summary of PFAS-shared metabolite-clinical marker chains in dataset1

Table S25. Serial mediation summary by disease-priority PFAS and omics mediator block in dataset1

Table S26. Serial mediation summary by clinical marker and omics mediator block in dataset1

Table S27. Network nodes for integrated PFAS-omics-clinical-ovarian cancer figures in dataset1

#### Part 8. Shared-metabolite biomarker validation models

Table S28. Feature sets used for shared-metabolite biomarker validation models using logistic regression

Table S29. Incremental performance comparisons for Core-5 shared-metabolite feature sets using logistic regression

Table S30. Feature sets used for shared-metabolite biomarker validation models using XGBoost

Table S31. Incremental performance comparisons for Core-5 shared-metabolite feature sets using XGBoost.

#### Part 9. Targeted PFAS assay parameters and validation

Table S32. Optimized multiple reaction monitoring (MRM) parameters for targeted PFAS analysis

Table S33. Calibration equations, coefficients of determination, limits of detection and quantification, recovery, and precis ion for targeted PFAS analysis

**Supplementary Table S1. Clinical and laboratory characteristics by case–control status in dataset1.**

| Marker | Characteristic | Category | Unit | Control (n = 128) | Case (n = 128) | P value |
|---|---|---|---|---|---|---|
| N | No. of participants | | | 128 | 128 | |
| Age | Age, median [IQR] | | years | 54.00 [47.00, 61.25] | 56.50 [52.00, 65.00] | 0.059 |
| BMI | Body mass index, median [IQR] | | kg/m² | 22.68 [20.96, 24.48] | 22.88 [21.11, 25.18] | 0.790 |
| Hypertension | Hypertension | No | n (%) | 79 (61.7) | 105 (82.0) | 0.001 |
| | | Yes | | 49 (38.3) | 23 (18.0) | |
| CEA | Carcinoembryonic antigen, median [IQR] | | ng/mL | 1.21 [0.84, 1.55] | 1.79 [1.03, 3.32] | <0.001 |
| ALB | Albumin, median [IQR] | | g/L | 46.05 [44.08, 47.43] | 45.95 [43.05, 47.70] | 0.290 |
| WBC | White blood cell count, median [IQR] | | 10^9/L | 5.36 [4.42, 6.46] | 5.46 [4.04, 7.41] | 0.803 |
| P-LCR | Platelet large cell ratio, median [IQR] | | % | 26.88 [22.28, 32.02] | 28.05 [20.65, 33.35] | 0.452 |
| MON | Monocyte count, median [IQR] | | 10^9/L | 0.36 [0.30, 0.44] | 0.39 [0.30, 0.55] | 0.062 |
| TC | Total cholesterol, median [IQR] | | mmol/L | 5.04 [4.52, 5.58] | 5.20 [4.62, 6.16] | 0.048 |
| LDL-C | LDL cholesterol, median [IQR] | | mmol/L | 2.96 [2.44, 3.42] | 3.29 [2.66, 4.00] | 0.002 |
| TG | Triglycerides, median [IQR] | | mmol/L | 1.04 [0.68, 1.46] | 1.79 [1.17, 2.41] | <0.001 |
| HDL-C | HDL cholesterol, mean (SD) | | mmol/L | 1.62 (0.39) | 1.31 (0.31) | <0.001 |
| GGT | Gamma-glutamyl transferase, median [IQR] | | U/L | 16.00 [11.00, 25.25] | 28.50 [19.00, 50.25] | <0.001 |
| ALT | Alanine aminotransferase, median [IQR] | | U/L | 15.00 [12.00, 21.00] | 18.00 [13.00, 31.00] | 0.002 |

| | | | | | |
|---|---|---|---|---|---|
| AST | Aspartate aminotransferase, median [IQR] | U/L | 19.00 [16.00, 22.00] | 23.00 [18.00, 31.00] | <0.001 |
| RDW-SD | Red cell distribution width-standard deviation, median [IQR] | fL | 42.45 [40.50, 44.05] | 52.35 [45.94, 58.02] | <0.001 |
| AFP | Alpha-fetoprotein, median [IQR] | ng/mL | 2.52 [1.49, 4.08] | 2.70 [1.88, 3.92] | 0.205 |
| ALP | Alkaline phosphatase, median [IQR] | U/L | 75.00 [62.75, 89.00] | 90.00 [76.00, 110.75] | <0.001 |
| LYM | Lymphocyte count, median [IQR] | 10^9/L | 1.77 [1.51, 2.18] | 1.35 [1.04, 1.74] | <0.001 |
| MCH | Mean corpuscular hemoglobin, median [IQR] | pg | 30.40 [29.70, 31.10] | 31.55 [29.50, 33.50] | <0.001 |
| MCHC | Mean corpuscular hemoglobin concentration, median [IQR] | g/L | 326.00 [321.00, 332.00] | 331.00 [324.00, 336.00] | <0.001 |
| MCV | Mean corpuscular volume, median [IQR] | fL | 92.85 [90.07, 94.90] | 94.90 [90.30, 101.93] | 0.001 |
| MPV | Mean platelet volume, mean (SD) | fL | 10.29 (0.83) | 10.41 (1.19) | 0.387 |
| GLU | Glucose, median [IQR] | mmol/L | 4.87 [4.65, 5.21] | 5.49 [4.91, 6.40] | <0.001 |
| GLOB | Globulin, mean (SD) | g/L | 25.85 (3.15) | 28.44 (5.04) | <0.001 |
| LDH | Lactate dehydrogenase, median [IQR] | U/L | 201.00 [185.25, 224.25] | 210.00 [182.75, 271.25] | 0.061 |
| EOS | Eosinophil count, median [IQR] | 10^9/L | 0.08 [0.05, 0.14] | 0.06 [0.03, 0.13] | 0.004 |
| CA125 | Cancer antigen 125, median [IQR] | U/mL | 11.98 [8.33, 16.27] | 69.10 [13.85, 337.00] | <0.001 |
| CA15-3 | Cancer antigen 15-3, median [IQR] | U/mL | 8.55 [6.28, 11.83] | 24.30 [13.35, 50.70] | <0.001 |
| DBIL | Direct bilirubin, median [IQR] | μmol/L | 3.54 [2.80, 4.74] | 3.20 [2.68, 4.12] | 0.115 |
| NEU | Neutrophil count, median [IQR] | 10^9/L | 3.20 [2.31, 3.86] | 3.27 [2.34, 5.41] | 0.058 |

| | | | | | |
|---|---|---|---|---|---|
| TBIL | Total bilirubin, median [IQR] | μmol/L | 9.30 [6.73, 13.33] | 8.25 [6.30, 11.83] | 0.126 |
| TP | Total protein, median [IQR] | g/L | 72.20 [69.47, 75.11] | 74.10 [68.50, 77.35] | 0.144 |

*Note: Values are presented as mean (SD), median [IQR], or n (%), as shown.*

*P values and FDR q values are reported as <0.001 when below 0.001; otherwise they are shown to three decimal places.*

**Supplementary Table S2. Clinical and laboratory characteristics of controls and ovarian cancer cases by FIGO stage group in dataset1.**

| Marker | Characteristic | Category | Unit | Control (n = 128) | FIGO stage I-II (n = 55) | FIGO stage III-IV (n = 73) | P value |
|---|---|---|---|---|---|---|---|
| N | No. of participants | | | 128 | 55 | 73 | |
| Age | Age, median [IQR] | | years | 54.00 [47.00, 61.25] | 56.00 [49.00, 62.50] | 57.00 [53.00, 66.00] | 0.061 |
| BMI | Body mass index, median [IQR] | | kg/m² | 22.68 [20.96, 24.48] | 22.43 [20.57, 25.04] | 23.05 [21.26, 25.39] | 0.745 |
| Hypertension | Hypertension | No | n (%) | 79 (61.7) | 44 (80.0) | 61 (83.6) | 0.001 |
| | | Yes | | 49 (38.3) | 11 (20.0) | 12 (16.4) | |
| CEA | Carcinoembryonic antigen, median [IQR] | | ng/mL | 1.21 [0.84, 1.55] | 1.62 [1.11, 3.67] | 1.89 [0.98, 2.85] | <0.001 |
| ALB | Albumin, median [IQR] | | g/L | 46.05 [44.08, 47.43] | 46.00 [43.45, 47.76] | 45.90 [42.90, 47.70] | 0.570 |
| WBC | White blood cell count, median [IQR] | | 10^9/L | 5.36 [4.42, 6.46] | 5.19 [4.01, 7.81] | 5.54 [4.31, 7.40] | 0.898 |
| P-LCR | Platelet large cell ratio, median [IQR] | | % | 26.88 [22.28, 32.02] | 28.10 [22.25, 31.90] | 27.30 [20.50, 35.00] | 0.752 |
| MON | Monocyte count, median [IQR] | | 10^9/L | 0.36 [0.30, 0.44] | 0.36 [0.27, 0.55] | 0.40 [0.32, 0.55] | 0.039 |
| TC | Total cholesterol, median [IQR] | | mmol/L | 5.04 [4.52, 5.58] | 5.00 [4.11, 5.30] | 5.38 [4.89, 6.48] | <0.001 |
| LDL-C | LDL cholesterol, median [IQR] | | mmol/L | 2.96 [2.44, 3.42] | 3.01 [2.43, 3.72] | 3.53 [2.73, 4.13] | <0.001 |
| TG | Triglycerides, median [IQR] | | mmol/L | 1.04 [0.68, 1.46] | 1.61 [1.06, 2.43] | 1.82 [1.33, 2.37] | <0.001 |
| HDL-C | HDL cholesterol, mean (SD) | | mmol/L | 1.62 (0.39) | 1.31 (0.28) | 1.32 (0.33) | <0.001 |

| | | | | | | |
|---|---|---|---|---|---|---|
| GGT | Gamma-glutamyl transferase, median [IQR] | U/L | 16.00 [11.00, 25.25] | 28.00 [20.00, 53.00] | 29.00 [18.00, 50.00] | <0.001 |
| ALT | Alanine aminotransferase, median [IQR] | U/L | 15.00 [12.00, 21.00] | 18.00 [12.50, 28.50] | 19.00 [13.00, 31.00] | 0.008 |
| AST | Aspartate aminotransferase, median [IQR] | U/L | 19.00 [16.00, 22.00] | 21.00 [17.50, 27.50] | 24.00 [18.00, 33.00] | <0.001 |
| RDW-SD | Red cell distribution width-standard deviation, median [IQR] | fL | 42.45 [40.50, 44.05] | 50.20 [45.75, 57.35] | 53.73 [46.20, 58.90] | <0.001 |
| AFP | Alpha-fetoprotein, median [IQR] | ng/mL | 2.52 [1.49, 4.08] | 2.44 [1.93, 3.49] | 2.80 [1.84, 4.11] | 0.329 |
| ALP | Alkaline phosphatase, median [IQR] | U/L | 75.00 [62.75, 89.00] | 93.00 [76.00, 108.00] | 89.00 [76.00, 113.00] | <0.001 |
| LYM | Lymphocyte count, median [IQR] | 10^9/L | 1.77 [1.51, 2.18] | 1.49 [1.04, 1.80] | 1.29 [1.08, 1.58] | <0.001 |
| MCH | Mean corpuscular hemoglobin, median [IQR] | pg | 30.40 [29.70, 31.10] | 31.10 [29.50, 33.00] | 31.60 [29.50, 34.10] | <0.001 |
| MCHC | Mean corpuscular hemoglobin concentration, median [IQR] | g/L | 326.00 [321.00, 332.00] | 331.00 [323.00, 335.00] | 331.00 [326.00, 336.00] | 0.001 |
| MCV | Mean corpuscular volume, median [IQR] | fL | 92.85 [90.07, 94.90] | 93.40 [89.45, 97.63] | 96.20 [91.20, 103.40] | <0.001 |
| MPV | Mean platelet volume, median [IQR] | fL | 10.26 [9.70, 10.90] | 10.50 [9.60, 10.90] | 10.30 [9.50, 11.40] | 0.732 |
| GLU | Glucose, median [IQR] | mmol/L | 4.87 [4.65, 5.21] | 5.47 [4.84, 6.28] | 5.59 [4.92, 6.40] | <0.001 |
| GLOB | Globulin, mean (SD) | g/L | 25.85 (3.15) | 28.29 (4.82) | 28.56 (5.22) | <0.001 |
| LDH | Lactate dehydrogenase, median [IQR] | U/L | 201.00 [185.25, 224.25] | 207.00 [176.50, 291.50] | 216.00 [186.00, 262.00] | 0.148 |

| | | | | | | |
|---|---|---|---|---|---|---|
| EOS | Eosinophil count, median [IQR] | 10^9/L | 0.08 [0.05, 0.14] | 0.07 [0.04, 0.15] | 0.05 [0.02, 0.13] | 0.010 |
| CA125 | Cancer antigen 125, median [IQR] | U/mL | 11.98 [8.33, 16.27] | 35.70 [10.55, 308.87] | 93.10 [19.80, 376.00] | <0.001 |
| CA15-3 | Cancer antigen 15-3, median [IQR] | U/mL | 8.55 [6.28, 11.83] | 19.10 [11.70, 38.60] | 27.40 [16.60, 52.50] | <0.001 |
| DBIL | Direct bilirubin, median [IQR] | μmol/L | 3.54 [2.80, 4.74] | 3.40 [2.55, 4.50] | 3.20 [2.70, 3.90] | 0.212 |
| NEU | Neutrophil count, median [IQR] | 10^9/L | 3.20 [2.31, 3.86] | 3.05 [2.32, 5.39] | 3.43 [2.51, 5.40] | 0.117 |
| TBIL | Total bilirubin, median [IQR] | μmol/L | 9.30 [6.73, 13.33] | 9.35 [6.10, 12.07] | 8.00 [6.40, 11.00] | 0.201 |
| TP | Total protein, median [IQR] | g/L | 72.20 [69.47, 75.11] | 74.20 [67.80, 78.85] | 74.10 [70.00, 76.70] | 0.307 |

*Note: Values are presented as mean (SD), median [IQR], or n (%), as shown.*

*P values and FDR q values are reported as <0.001 when below 0.001; otherwise they are shown to three decimal places.*

**Supplementary Table S3. Clinical and laboratory characteristics by FIGO stage group among ovarian cancer cases in dataset1.**

| Marker | Characteristic | Category | Unit | FIGO stage I-II (n = 55) | FIGO stage III-IV (n = 73) | P value |
|---|---|---|---|---|---|---|
| N | No. of participants | | | 55 | 73 | |
| Age | Age, mean (SD) | | years | 55.33 (11.18) | 58.51 (9.44) | 0.084 |
| BMI | Body mass index, median [IQR] | | kg/m² | 22.43 [20.57, 25.04] | 23.05 [21.26, 25.39] | 0.511 |
| Hypertension | Hypertension | No | n (%) | 44 (80.0) | 61 (83.6) | 0.774 |
| | | Yes | | 11 (20.0) | 12 (16.4) | |
| CEA | Carcinoembryonic antigen, median [IQR] | | ng/mL | 1.62 [1.11, 3.67] | 1.89 [0.98, 2.85] | 0.975 |
| ALB | Albumin, median [IQR] | | g/L | 46.00 [43.45, 47.76] | 45.90 [42.90, 47.70] | 0.941 |
| WBC | White blood cell count, median [IQR] | | 10^9/L | 5.19 [4.01, 7.81] | 5.54 [4.31, 7.40] | 0.810 |
| P-LCR | Platelet large cell ratio, median [IQR] | | % | 28.10 [22.25, 31.90] | 27.30 [20.50, 35.00] | 0.840 |
| MON | Monocyte count, median [IQR] | | 10^9/L | 0.36 [0.27, 0.55] | 0.40 [0.32, 0.55] | 0.162 |
| TC | Total cholesterol, mean (SD) | | mmol/L | 4.94 (1.04) | 5.59 (1.13) | 0.001 |
| LDL-C | LDL cholesterol, mean (SD) | | mmol/L | 3.11 (1.01) | 3.55 (1.07) | 0.020 |
| TG | Triglycerides, median [IQR] | | mmol/L | 1.61 [1.06, 2.43] | 1.82 [1.33, 2.37] | 0.277 |
| HDL-C | HDL cholesterol, mean (SD) | | mmol/L | 1.31 (0.28) | 1.32 (0.33) | 0.850 |
| GGT | Gamma-glutamyl transferase, median [IQR] | | U/L | 28.00 [20.00, 53.00] | 29.00 [18.00, 50.00] | 0.900 |
| ALT | Alanine aminotransferase, median [IQR] | | U/L | 18.00 [12.50, 28.50] | 19.00 [13.00, 31.00] | 0.981 |

| | | | | | |
|---|---|---|---|---|---|
| AST | Aspartate aminotransferase, median [IQR] | U/L | 21.00 [17.50, 27.50] | 24.00 [18.00, 33.00] | 0.191 |
| RDW-SD | Red cell distribution width-standard deviation, median [IQR] | fL | 50.20 [45.75, 57.35] | 53.73 [46.20, 58.90] | 0.115 |
| AFP | Alpha-fetoprotein, median [IQR] | ng/mL | 2.44 [1.93, 3.49] | 2.80 [1.84, 4.11] | 0.393 |
| ALP | Alkaline phosphatase, median [IQR] | U/L | 93.00 [76.00, 108.00] | 89.00 [76.00, 113.00] | 0.522 |
| LYM | Lymphocyte count, median [IQR] | 10^9/L | 1.49 [1.04, 1.80] | 1.29 [1.08, 1.58] | 0.443 |
| MCH | Mean corpuscular hemoglobin, mean (SD) | pg | 31.26 (3.02) | 31.88 (3.30) | 0.278 |
| MCHC | Mean corpuscular hemoglobin concentration, median [IQR] | g/L | 331.00 [323.00, 335.00] | 331.00 [326.00, 336.00] | 0.709 |
| MCV | Mean corpuscular volume, mean (SD) | fL | 93.63 (7.72) | 97.15 (8.21) | 0.015 |
| MPV | Mean platelet volume, median [IQR] | fL | 10.50 [9.60, 10.90] | 10.30 [9.50, 11.40] | 0.464 |
| GLU | Glucose, median [IQR] | mmol/L | 5.47 [4.84, 6.28] | 5.59 [4.92, 6.40] | 0.460 |
| GLOB | Globulin, mean (SD) | g/L | 28.29 (4.82) | 28.56 (5.22) | 0.761 |
| LDH | Lactate dehydrogenase, median [IQR] | U/L | 207.00 [176.50, 291.50] | 216.00 [186.00, 262.00] | 0.698 |
| EOS | Eosinophil count, median [IQR] | 10^9/L | 0.07 [0.04, 0.15] | 0.05 [0.02, 0.13] | 0.394 |
| CA125 | Cancer antigen 125, median [IQR] | U/mL | 35.70 [10.55, 308.87] | 93.10 [19.80, 376.00] | 0.160 |
| CA15-3 | Cancer antigen 15-3, median [IQR] | U/mL | 19.10 [11.70, 38.60] | 27.40 [16.60, 52.50] | 0.068 |
| DBIL | Direct bilirubin, median [IQR] | μmol/L | 3.40 [2.55, 4.50] | 3.20 [2.70, 3.90] | 0.520 |
| NEU | Neutrophil count, median [IQR] | 10^9/L | 3.05 [2.32, 5.39] | 3.43 [2.51, 5.40] | 0.490 |

| | | | | | |
|---|---|---|---|---|---|
| TBIL | Total bilirubin, median [IQR] | μmol/L | 9.35 [6.10, 12.07] | 8.00 [6.40, 11.00] | 0.391 |
| TP | Total protein, median [IQR] | g/L | 74.20 [67.80, 78.85] | 74.10 [70.00, 76.70] | 0.950 |

*Note: Values are presented as mean (SD), median [IQR], or n (%), as shown.*

*P values and FDR q values are reported as <0.001 when below 0.001; otherwise they are shown to three decimal places.*

**Supplementary Table S4. Clinical and laboratory characteristics by case–control status in dataset2.**

| Marker | Characteristic | Category | Unit | Control (n = 100) | Case (n = 100) | P value |
|---|---|---|---|---|---|---|
| N | No. of participants | | | 100 | 100 | |
| Age | Age, median [IQR] | | years | 55.00 [46.00, 60.00] | 56.00 [51.00, 63.00] | 0.056 |
| BMI | Body mass index, median [IQR] | | kg/m² | 22.82 [21.16, 24.92] | 23.32 [21.26, 25.59] | 0.649 |
| Hypertension | Hypertension | No | n (%) | 72 (72.0) | 68 (68.0) | 0.643 |
| | | Yes | | 28 (28.0) | 32 (32.0) | |
| urine_cr_mmol_l | Urinary creatinine concentration, median [IQR] | | mmol/L | 8.96 [5.61, 13.31] | 7.99 [4.25, 12.20] | 0.074 |
| EOS | Eosinophil count, median [IQR] | | 10^9/L | 0.10 [0.05, 0.16] | 0.06 [0.03, 0.12] | <0.001 |
| HGB | Hemoglobin, median [IQR] | | g/L | 133.50 [126.00, 138.25] | 117.00 [107.00, 127.00] | <0.001 |
| LYM | Lymphocyte count, median [IQR] | | 10^9/L | 1.77 [1.46, 2.16] | 1.21 [0.93, 1.74] | <0.001 |
| MCH | Mean corpuscular hemoglobin, median [IQR] | | pg | 30.25 [29.20, 30.90] | 30.90 [29.37, 32.60] | 0.005 |
| MCHC | Mean corpuscular hemoglobin concentration, median [IQR] | | g/L | 326.50 [319.75, 332.00] | 329.00 [321.75, 334.00] | 0.073 |
| MCV | Mean corpuscular volume, median [IQR] | | fL | 91.80 [89.88, 94.42] | 94.40 [89.88, 98.20] | 0.005 |
| MON | Monocyte count, median [IQR] | | 10^9/L | 0.35 [0.28, 0.44] | 0.41 [0.32, 0.56] | 0.028 |
| MPV | Mean platelet volume, median [IQR] | | fL | 10.10 [9.67, 10.60] | 10.15 [9.30, 11.50] | 0.524 |

| | | | | | |
|---|---|---|---|---|---|
| NEU | Neutrophil count, median [IQR] | 10^9/L | 3.22 [2.46, 4.09] | 3.84 [2.42, 5.93] | 0.043 |
| P-LCR | Platelet large cell ratio, median [IQR] | % | 26.30 [21.35, 29.50] | 26.10 [20.20, 36.30] | 0.381 |
| RDW-SD | Red cell distribution width-standard deviation, median [IQR] | fL | 42.80 [41.10, 45.32] | 48.35 [42.50, 57.47] | <0.001 |
| WBC | White blood cell count, median [IQR] | 10^9/L | 5.62 [4.73, 6.49] | 5.64 [4.15, 7.66] | 0.661 |
| ALB | Albumin, median [IQR] | g/L | 45.40 [44.30, 46.92] | 45.20 [41.32, 48.08] | 0.433 |
| ALP | Alkaline phosphatase, median [IQR] | U/L | 72.00 [61.00, 79.25] | 87.50 [74.50, 112.00] | <0.001 |
| ALT | Alanine aminotransferase, median [IQR] | U/L | 14.00 [11.00, 20.00] | 18.00 [12.00, 29.25] | 0.006 |
| AST | Aspartate aminotransferase, median [IQR] | U/L | 19.00 [16.00, 22.00] | 23.00 [19.00, 29.00] | <0.001 |
| DBIL | Direct bilirubin, median [IQR] | μmol/L | 3.80 [2.81, 4.95] | 3.30 [2.68, 4.50] | 0.199 |
| GGT | Gamma-glutamyl transferase, median [IQR] | U/L | 15.00 [11.00, 21.25] | 27.00 [17.00, 51.25] | <0.001 |
| GLOB | Globulin, mean (SD) | g/L | 25.90 (4.06) | 27.73 (4.52) | 0.003 |
| GLU | Glucose, median [IQR] | mmol/L | 4.88 [4.59, 5.26] | 5.59 [5.13, 6.27] | <0.001 |
| HDL-C | HDL cholesterol, median [IQR] | mmol/L | 1.52 [1.33, 1.80] | 1.26 [1.03, 1.57] | <0.001 |
| LDH | Lactate dehydrogenase, median [IQR] | U/L | 206.00 [185.00, 223.25] | 201.50 [174.00, 234.00] | 0.821 |
| LDL-C | LDL cholesterol, mean (SD) | mmol/L | 2.80 (0.71) | 3.11 (0.94) | 0.009 |
| TC | Total cholesterol, median [IQR] | mmol/L | 4.99 [4.23, 5.69] | 5.12 [4.32, 5.78] | 0.443 |

| | | | | | | |
|---|---|---|---|---|---|---|
| TG | Triglycerides, median [IQR] | | mmol/L | 1.06 [0.76, 1.42] | 1.67 [1.27, 2.31] | <0.001 |
| TP | Total protein, median [IQR] | | g/L | 70.55 [67.80, 73.85] | 72.15 [67.60, 77.05] | 0.084 |
| UREA | Urea, median [IQR] | | mmol/L | 4.85 [4.10, 5.73] | 5.21 [4.42, 6.34] | 0.031 |
| UA | Uric acid, median [IQR] | | μmol/L | 265.50 [235.00, 317.00] | 289.50 [235.75, 357.25] | 0.236 |
| AFP | Alpha-fetoprotein, median [IQR] | | ng/mL | 2.49 [1.83, 3.32] | 2.86 [2.09, 4.17] | 0.019 |
| CA125 | Cancer antigen 125, median [IQR] | | U/mL | 10.85 [7.58, 15.53] | 45.30 [15.28, 161.25] | <0.001 |
| CA15-3 | Cancer antigen 15-3, median [IQR] | | U/mL | 10.30 [6.89, 14.35] | 22.55 [15.10, 39.80] | <0.001 |
| CEA | Carcinoembryonic antigen, median [IQR] | | ng/mL | 1.29 [0.91, 1.71] | 1.77 [1.22, 2.93] | <0.001 |
| BIL_CODE | Urine bilirubin | Negative | n (%) | 100 (100.0) | 99 (99.0) | 1.000 |
| | | 1+ | | 0 (0.0) | 1 (1.0) | |
| BLD_CODE | Urine occult blood | Negative | n (%) | 34 (34.7) | 63 (63.6) | <0.001 |
| | | Trace | | 32 (32.7) | 18 (18.2) | |
| | | 1+ | | 14 (14.3) | 12 (12.1) | |
| | | 2+ | | 14 (14.3) | 2 (2.0) | |
| | | 3+ | | 4 (4.1) | 4 (4.0) | |
| URINE_GLU_CODE | Urine glucose | Negative | n (%) | 92 (92.0) | 95 (95.0) | 0.296 |
| | | Trace | | 1 (1.0) | 0 (0.0) | |

| | | | | | | |
|---|---|---|---|---|---|---|
| | | 1+ | | 0 (0.0) | 1 (1.0) | |
| | | 3+ | | 5 (5.0) | 1 (1.0) | |
| | | 4+ | | 2 (2.0) | 3 (3.0) | |
| KET_CODE | Urine ketones | Negative | n (%) | 94 (94.0) | 95 (95.0) | 0.472 |
| | | Trace | | 2 (2.0) | 0 (0.0) | |
| | | 1+ | | 3 (3.0) | 2 (2.0) | |
| | | 2+ | | 1 (1.0) | 2 (2.0) | |
| | | 3+ | | 0 (0.0) | 1 (1.0) | |
| NIT_CODE | Urine nitrite | Negative | n (%) | 100 (100.0) | 93 (93.0) | 0.021 |
| | | Positive | | 0 (0.0) | 7 (7.0) | |
| PRO_CODE | Urine protein | Negative | n (%) | 100 (100.0) | 75 (75.0) | <0.001 |
| | | Trace | | 0 (0.0) | 15 (15.0) | |
| | | 1+ | | 0 (0.0) | 9 (9.0) | |
| | | 3+ | | 0 (0.0) | 1 (1.0) | |
| UBG_CODE | Urine urobilinogen | Negative | n (%) | 100 (100.0) | 100 (100.0) | |

*Note: Values are presented as mean (SD), median [IQR], or n (%), as shown.*

*P values and FDR q values are reported as <0.001 when below 0.001; otherwise they are shown to three decimal places.*

**Supplementary Table S5. Clinical and laboratory characteristics of controls and ovarian cancer cases by FIGO stage group in dataset2.**

| Marker | Characteristic | Category | Unit | Control (n = 100) | FIGO stage I-II (n = 42) | FIGO stage III-IV (n = 58) | P value |
|---|---|---|---|---|---|---|---|
| N | No. of participants | | | 100 | 42 | 58 | |
| Age | Age, median [IQR] | | years | 55.00 [46.00, 60.00] | 54.00 [50.00, 60.75] | 57.50 [52.25, 65.00] | 0.050 |
| BMI | Body mass index, median [IQR] | | kg/m² | 22.82 [21.16, 24.92] | 24.14 [21.54, 26.14] | 22.77 [21.15, 24.92] | 0.326 |
| Hypertension | Hypertension | No | n (%) | 72 (72.0) | 29 (69.0) | 39 (67.2) | 0.811 |
| | | Yes | | 28 (28.0) | 13 (31.0) | 19 (32.8) | |
| urine_cr_mmol_l | Urinary creatinine concentration, median [IQR] | | mmol/L | 8.96 [5.61, 13.31] | 8.33 [3.83, 12.21] | 7.64 [4.38, 11.91] | 0.199 |
| EOS | Eosinophil count, median [IQR] | | 10^9/L | 0.10 [0.05, 0.16] | 0.06 [0.04, 0.14] | 0.05 [0.02, 0.09] | 0.001 |
| HGB | Hemoglobin, median [IQR] | | g/L | 133.50 [126.00, 138.25] | 116.50 [107.25, 127.00] | 117.00 [106.25, 126.00] | <0.001 |
| LYM | Lymphocyte count, median [IQR] | | 10^9/L | 1.77 [1.46, 2.16] | 1.39 [1.04, 1.88] | 1.12 [0.85, 1.55] | <0.001 |
| MCH | Mean corpuscular hemoglobin, median [IQR] | | pg | 30.25 [29.20, 30.90] | 30.95 [28.82, 32.65] | 30.90 [29.50, 32.53] | 0.020 |
| MCHC | Mean corpuscular hemoglobin concentration, median [IQR] | | g/L | 326.50 [319.75, 332.00] | 331.00 [322.00, 334.75] | 328.50 [320.50, 332.00] | 0.110 |
| MCV | Mean corpuscular volume, median [IQR] | | fL | 91.80 [89.88, 94.42] | 93.45 [89.15, 98.33] | 94.70 [90.82, 98.07] | 0.012 |
| MON | Monocyte count, median [IQR] | | 10^9/L | 0.35 [0.28, 0.44] | 0.35 [0.32, 0.54] | 0.42 [0.30, 0.65] | 0.054 |

| | | | | | | |
|---|---|---|---|---|---|---|
| MPV | Mean platelet volume, median [IQR] | fL | 10.10 [9.67, 10.60] | 10.25 [9.38, 11.17] | 10.00 [9.30, 11.50] | 0.745 |
| NEU | Neutrophil count, median [IQR] | 10^9/L | 3.22 [2.46, 4.09] | 3.83 [2.70, 6.12] | 3.88 [2.31, 5.66] | 0.124 |
| P-LCR | Platelet large cell ratio, median [IQR] | % | 26.30 [21.35, 29.50] | 26.75 [20.70, 35.05] | 25.20 [19.60, 36.60] | 0.651 |
| RDW-SD | Red cell distribution width-standard deviation, median [IQR] | fL | 42.80 [41.10, 45.32] | 48.25 [42.58, 55.80] | 48.35 [42.67, 58.93] | <0.001 |
| WBC | White blood cell count, median [IQR] | 10^9/L | 5.62 [4.73, 6.49] | 5.56 [4.95, 7.60] | 5.77 [3.97, 7.66] | 0.727 |
| ALB | Albumin, median [IQR] | g/L | 45.40 [44.30, 46.92] | 44.60 [40.52, 47.85] | 45.45 [43.32, 48.45] | 0.247 |
| ALP | Alkaline phosphatase, median [IQR] | U/L | 72.00 [61.00, 79.25] | 86.00 [71.50, 104.75] | 92.00 [76.00, 113.75] | <0.001 |
| ALT | Alanine aminotransferase, median [IQR] | U/L | 14.00 [11.00, 20.00] | 17.00 [12.25, 27.25] | 19.50 [12.25, 29.75] | 0.018 |
| AST | Aspartate aminotransferase, median [IQR] | U/L | 19.00 [16.00, 22.00] | 20.50 [18.00, 28.75] | 23.00 [19.00, 28.75] | <0.001 |
| DBIL | Direct bilirubin, median [IQR] | μmol/L | 3.80 [2.81, 4.95] | 3.35 [2.70, 4.75] | 3.30 [2.62, 4.47] | 0.423 |
| GGT | Gamma-glutamyl transferase, median [IQR] | U/L | 15.00 [11.00, 21.25] | 25.50 [17.25, 49.50] | 27.00 [17.25, 51.75] | <0.001 |
| GLOB | Globulin, mean (SD) | g/L | 25.90 (4.06) | 27.21 (4.62) | 28.11 (4.45) | 0.007 |
| GLU | Glucose, median [IQR] | mmol/L | 4.88 [4.59, 5.26] | 5.43 [5.12, 6.16] | 5.64 [5.16, 6.38] | <0.001 |
| HDL-C | HDL cholesterol, median [IQR] | mmol/L | 1.52 [1.33, 1.80] | 1.16 [0.99, 1.46] | 1.37 [1.04, 1.61] | <0.001 |

| | | | | | | | |
|---|---|---|---|---|---|---|---|
| LDH | Lactate dehydrogenase, median [IQR] | | U/L | 206.00 [185.00, 223.25] | 199.00 [171.00, 226.75] | 205.00 [176.50, 243.50] | 0.382 |
| LDL-C | LDL cholesterol, mean (SD) | | mmol/L | 2.80 (0.71) | 2.86 (0.92) | 3.30 (0.91) | 0.001 |
| TC | Total cholesterol, median [IQR] | | mmol/L | 4.99 [4.23, 5.69] | 4.63 [4.12, 5.35] | 5.34 [4.66, 5.91] | 0.035 |
| TG | Triglycerides, median [IQR] | | mmol/L | 1.06 [0.76, 1.42] | 1.90 [1.12, 2.34] | 1.62 [1.33, 2.18] | <0.001 |
| TP | Total protein, mean (SD) | | g/L | 70.97 (4.08) | 70.62 (8.05) | 72.80 (7.51) | 0.129 |
| UREA | Urea, median [IQR] | | mmol/L | 4.85 [4.10, 5.73] | 5.26 [3.94, 6.04] | 5.18 [4.50, 6.76] | 0.056 |
| UA | Uric acid, median [IQR] | | μmol/L | 265.50 [235.00, 317.00] | 286.00 [242.25, 354.00] | 289.50 [226.25, 358.75] | 0.391 |
| AFP | Alpha-fetoprotein, median [IQR] | | ng/mL | 2.49 [1.83, 3.32] | 2.78 [2.05, 3.84] | 3.20 [2.12, 4.27] | 0.039 |
| CA125 | Cancer antigen 125, median [IQR] | | U/mL | 10.85 [7.58, 15.53] | 45.30 [16.95, 148.00] | 48.25 [14.82, 165.25] | <0.001 |
| CA15-3 | Cancer antigen 15-3, median [IQR] | | U/mL | 10.30 [6.89, 14.35] | 22.50 [14.03, 33.27] | 24.60 [17.10, 43.90] | <0.001 |
| CEA | Carcinoembryonic antigen, median [IQR] | | ng/mL | 1.29 [0.91, 1.71] | 1.57 [1.15, 3.03] | 1.90 [1.28, 2.82] | <0.001 |
| BIL_CODE | Urine bilirubin | Negative | n (%) | 100 (100.0) | 42 (100.0) | 57 (98.3) | 0.292 |
| | | 1+ | | 0 (0.0) | 0 (0.0) | 1 (1.7) | |
| BLD_CODE | Urine occult blood | Negative | n (%) | 34 (34.7) | 26 (61.9) | 37 (64.9) | 0.003 |
| | | Trace | | 32 (32.7) | 10 (23.8) | 8 (14.0) | |
| | | 1+ | | 14 (14.3) | 5 (11.9) | 7 (12.3) | |

| | | | | | | | |
|---|---|---|---|---|---|---|---|
| | | 2+ | | 14 (14.3) | 0 (0.0) | 2 (3.5) | |
| | | 3+ | | 4 (4.1) | 1 (2.4) | 3 (5.3) | |
| URINE_GLU_CODE | Urine glucose | Negative | n (%) | 92 (92.0) | 40 (95.2) | 55 (94.8) | 0.544 |
| | | Trace | | 1 (1.0) | 0 (0.0) | 0 (0.0) | |
| | | 1+ | | 0 (0.0) | 0 (0.0) | 1 (1.7) | |
| | | 3+ | | 5 (5.0) | 1 (2.4) | 0 (0.0) | |
| | | 4+ | | 2 (2.0) | 1 (2.4) | 2 (3.4) | |
| KET_CODE | Urine ketones | Negative | n (%) | 94 (94.0) | 38 (90.5) | 57 (98.3) | 0.365 |
| | | Trace | | 2 (2.0) | 0 (0.0) | 0 (0.0) | |
| | | 1+ | | 3 (3.0) | 2 (4.8) | 0 (0.0) | |
| | | 2+ | | 1 (1.0) | 1 (2.4) | 1 (1.7) | |
| | | 3+ | | 0 (0.0) | 1 (2.4) | 0 (0.0) | |
| NIT_CODE | Urine nitrite | Negative | n (%) | 100 (100.0) | 39 (92.9) | 54 (93.1) | 0.027 |
| | | Positive | | 0 (0.0) | 3 (7.1) | 4 (6.9) | |
| PRO_CODE | Urine protein | Negative | n (%) | 100 (100.0) | 33 (78.6) | 42 (72.4) | <0.001 |
| | | Trace | | 0 (0.0) | 5 (11.9) | 10 (17.2) | |
| | | 1+ | | 0 (0.0) | 4 (9.5) | 5 (8.6) | |
| | | 3+ | | 0 (0.0) | 0 (0.0) | 1 (1.7) | |

| | | | | | | |
|---|---|---|---|---|---|---|
| UBG_CODE | Urine urobilinogen | Negative | n (%) | 100 (100.0) | 42 (100.0) | 58 (100.0) |

*Note: Values are presented as mean (SD), median [IQR], or n (%), as shown.*

*P values and FDR q values are reported as <0.001 when below 0.001; otherwise they are shown to three decimal places.*

**Supplementary Table S6. Clinical and laboratory characteristics by FIGO stage group among ovarian cancer cases in dataset2.**

| Marker | Characteristic | Category | Unit | FIGO stage I-II (n = 42) | FIGO stage III-IV (n = 58) | P value |
|---|---|---|---|---|---|---|
| N | No. of participants | | | 42 | 58 | |
| Age | Age, mean (SD) | | years | 55.57 (10.76) | 57.93 (8.55) | 0.225 |
| BMI | Body mass index, mean (SD) | | kg/m² | 24.08 (3.89) | 23.20 (3.02) | 0.206 |
| Hypertension | Hypertension | No | n (%) | 29 (69.0) | 39 (67.2) | 1.000 |
| | | Yes | | 13 (31.0) | 19 (32.8) | |
| urine_cr_mmol_l | Urinary creatinine concentration, median [IQR] | | mmol/L | 8.33 [3.83, 12.21] | 7.64 [4.38, 11.91] | 0.772 |
| EOS | Eosinophil count, median [IQR] | | 10^9/L | 0.06 [0.04, 0.14] | 0.05 [0.02, 0.09] | 0.181 |
| HGB | Hemoglobin, mean (SD) | | g/L | 116.38 (14.78) | 115.59 (15.35) | 0.796 |
| LYM | Lymphocyte count, median [IQR] | | 10^9/L | 1.39 [1.04, 1.88] | 1.12 [0.85, 1.55] | 0.066 |
| MCH | Mean corpuscular hemoglobin, mean (SD) | | pg | 30.89 (2.72) | 31.06 (2.74) | 0.762 |
| MCHC | Mean corpuscular hemoglobin concentration, median [IQR] | | g/L | 331.00 [322.00, 334.75] | 328.50 [320.50, 332.00] | 0.243 |
| MCV | Mean corpuscular volume, median [IQR] | | fL | 93.45 [89.15, 98.33] | 94.70 [90.82, 98.07] | 0.424 |
| MON | Monocyte count, median [IQR] | | 10^9/L | 0.35 [0.32, 0.54] | 0.42 [0.30, 0.65] | 0.299 |
| MPV | Mean platelet volume, median [IQR] | | fL | 10.25 [9.38, 11.17] | 10.00 [9.30, 11.50] | 0.719 |
| NEU | Neutrophil count, median [IQR] | | 10^9/L | 3.83 [2.70, 6.12] | 3.88 [2.31, 5.66] | 0.944 |

| | | | | | |
|---|---|---|---|---|---|
| P-LCR | Platelet large cell ratio, median [IQR] | % | 26.75 [20.70, 35.05] | 25.20 [19.60, 36.60] | 0.799 |
| RDW-SD | Red cell distribution width-standard deviation, median [IQR] | fL | 48.25 [42.58, 55.80] | 48.35 [42.67, 58.93] | 0.557 |
| WBC | White blood cell count, median [IQR] | 10^9/L | 5.56 [4.95, 7.60] | 5.77 [3.97, 7.66] | 0.600 |
| ALB | Albumin, median [IQR] | g/L | 44.60 [40.52, 47.85] | 45.45 [43.32, 48.45] | 0.199 |
| ALP | Alkaline phosphatase, median [IQR] | U/L | 86.00 [71.50, 104.75] | 92.00 [76.00, 113.75] | 0.252 |
| ALT | Alanine aminotransferase, median [IQR] | U/L | 17.00 [12.25, 27.25] | 19.50 [12.25, 29.75] | 0.461 |
| AST | Aspartate aminotransferase, median [IQR] | U/L | 20.50 [18.00, 28.75] | 23.00 [19.00, 28.75] | 0.413 |
| DBIL | Direct bilirubin, median [IQR] | µmol/L | 3.35 [2.70, 4.75] | 3.30 [2.62, 4.47] | 0.845 |
| GGT | Gamma-glutamyl transferase, median [IQR] | U/L | 25.50 [17.25, 49.50] | 27.00 [17.25, 51.75] | 0.714 |
| GLOB | Globulin, mean (SD) | g/L | 27.21 (4.62) | 28.11 (4.45) | 0.331 |
| GLU | Glucose, median [IQR] | mmol/L | 5.43 [5.12, 6.16] | 5.64 [5.16, 6.38] | 0.543 |
| HDL-C | HDL cholesterol, median [IQR] | mmol/L | 1.16 [0.99, 1.46] | 1.37 [1.04, 1.61] | 0.141 |
| LDH | Lactate dehydrogenase, median [IQR] | U/L | 199.00 [171.00, 226.75] | 205.00 [176.50, 243.50] | 0.199 |
| LDL-C | LDL cholesterol, mean (SD) | mmol/L | 2.86 (0.92) | 3.30 (0.91) | 0.020 |
| TC | Total cholesterol, mean (SD) | mmol/L | 4.74 (1.05) | 5.25 (1.16) | 0.025 |
| TG | Triglycerides, median [IQR] | mmol/L | 1.90 [1.12, 2.34] | 1.62 [1.33, 2.18] | 0.831 |
| TP | Total protein, mean (SD) | g/L | 70.62 (8.05) | 72.80 (7.51) | 0.167 |

| | | | | | | |
|---|---|---|---|---|---|---|
| UREA | Urea, median [IQR] | | mmol/L | 5.26 [3.94, 6.04] | 5.18 [4.50, 6.76] | 0.313 |
| UA | Uric acid, mean (SD) | | μmol/L | 299.19 (71.70) | 292.64 (86.01) | 0.688 |
| AFP | Alpha-fetoprotein, median [IQR] | | ng/mL | 2.78 [2.05, 3.84] | 3.20 [2.12, 4.27] | 0.330 |
| CA125 | Cancer antigen 125, median [IQR] | | U/mL | 45.30 [16.95, 148.00] | 48.25 [14.82, 165.25] | 1.000 |
| CA15-3 | Cancer antigen 15-3, median [IQR] | | U/mL | 22.50 [14.03, 33.27] | 24.60 [17.10, 43.90] | 0.300 |
| CEA | Carcinoembryonic antigen, median [IQR] | | ng/mL | 1.57 [1.15, 3.03] | 1.90 [1.28, 2.82] | 0.772 |
| BIL_CODE | Urine bilirubin | Negative | n (%) | 42 (100.0) | 57 (98.3) | 1.000 |
| | | 1+ | | 0 (0.0) | 1 (1.7) | |
| BLD_CODE | Urine occult blood | Negative | n (%) | 26 (61.9) | 37 (64.9) | 0.512 |
| | | Trace | | 10 (23.8) | 8 (14.0) | |
| | | 1+ | | 5 (11.9) | 7 (12.3) | |
| | | 2+ | | 0 (0.0) | 2 (3.5) | |
| | | 3+ | | 1 (2.4) | 3 (5.3) | |
| URINE_GLU_CODE | Urine glucose | Negative | n (%) | 40 (95.2) | 55 (94.8) | 0.532 |
| | | 1+ | | 0 (0.0) | 1 (1.7) | |
| | | 3+ | | 1 (2.4) | 0 (0.0) | |
| | | 4+ | | 1 (2.4) | 2 (3.4) | |
| KET_CODE | Urine ketones | Negative | n (%) | 38 (90.5) | 57 (98.3) | 0.226 |

| | | | | | | |
|---|---|---|---|---|---|---|
| | | 1+ | | 2 (4.8) | 0 (0.0) | |
| | | 2+ | | 1 (2.4) | 1 (1.7) | |
| | | 3+ | | 1 (2.4) | 0 (0.0) | |
| NIT_CODE | Urine nitrite | Negative | n (%) | 39 (92.9) | 54 (93.1) | 1.000 |
| | | Positive | | 3 (7.1) | 4 (6.9) | |
| PRO_CODE | Urine protein | Negative | n (%) | 33 (78.6) | 42 (72.4) | 0.722 |
| | | Trace | | 5 (11.9) | 10 (17.2) | |
| | | 1+ | | 4 (9.5) | 5 (8.6) | |
| | | 3+ | | 0 (0.0) | 1 (1.7) | |
| UBG_CODE | Urine urobilinogen | Negative | n (%) | 42 (100.0) | 58 (100.0) | |

*Note: Values are presented as mean (SD), median [IQR], or n (%), as shown.*

*P values and FDR q values are reported as <0.001 when below 0.001; otherwise they are shown to three decimal places.*

**Supplementary Table S7. Logistic regression adjustment models for clinical laboratory marker associations with ovarian cancer case status.**

*Note: Logistic regression modeled ovarian cancer case status (case = 1, control = 0).*

| Model | Formal table label | Adjustment variables | Role |
|---|---|---|---|
| Model 1 | Unadjusted | None | Crude association between each marker and ovarian cancer case status. |
| Model 2 | Age-adjusted | Age | Age-adjusted association. |
| Model 3 | Age- and BMI-adjusted | Age and BMI | Recommended primary adjusted model for the main table. |
| Model 4 | Sensitivity model | Age, BMI, and hypertension | Sensitivity model assessing robustness after comorbidity adjustment. Diabetes was not listed as an observed covariate in the available result sheet and is therefore not named in the formal table. |

**Supplementary Table S8. Associations of clinical laboratory markers with ovarian cancer case status in dataset1.**

*Note: Logistic regression modeled ovarian cancer case status (case = 1, control = 0).*

| Marker category | Marker | Unit | Effect scale | Primary model OR (95% CI) | P | FDR q value | Sensitivity model OR (95% CI) | P | FDR q value |
|---|---|---|---|---|---|---|---|---|---|
| CBC and leukocyte differential | EOS | *10^9/L | Per 1-unit increase | 0.112 (0.007 to 1.848) | 0.126 | 0.151 | 0.130 (0.008 to 2.249) | 0.161 | 0.193 |
| CBC and leukocyte differential | LYM | *10^9/L | Per 1-unit increase | 0.202 (0.116 to 0.354) | <0.001 | <0.001 | 0.199 (0.111 to 0.356) | <0.001 | <0.001 |
| CBC and leukocyte differential | MON | *10^9/L | Per 1-unit increase | 9.885 (2.210 to 44.215) | 0.003 | 0.004 | 10.590 (2.088 to 53.708) | 0.004 | 0.007 |
| CBC and leukocyte differential | NEU | *10^9/L | Per 1-unit increase | 1.255 (1.090 to 1.444) | 0.002 | 0.003 | 1.291 (1.108 to 1.503) | 0.001 | 0.002 |
| CBC and leukocyte differential | WBC | *10^9/L | Per 1-unit increase | 1.117 (1.019 to 1.224) | 0.018 | 0.025 | 1.123 (1.021 to 1.234) | 0.017 | 0.024 |
| Erythrocyte and hemoglobin indices | MCH | pg | Per 1-unit increase | 1.262 (1.127 to 1.413) | <0.001 | <0.001 | 1.282 (1.139 to 1.444) | <0.001 | <0.001 |
| Erythrocyte and hemoglobin indices | MCHC | g/L | Per 1-unit increase | 1.045 (1.018 to 1.073) | <0.001 | 0.002 | 1.052 (1.023 to 1.082) | <0.001 | <0.001 |
| Erythrocyte and hemoglobin indices | MCV | fL | Per 1-unit increase | 1.081 (1.037 to 1.127) | <0.001 | <0.001 | 1.080 (1.035 to 1.128) | <0.001 | <0.001 |
| Erythrocyte and hemoglobin indices | RDW-SD | fL | Per 1-unit increase | 1.405 (1.282 to 1.541) | <0.001 | <0.001 | 1.404 (1.277 to 1.543) | <0.001 | <0.001 |
| Platelet indices | MPV | fL | Per 1-unit increase | 1.127 (0.884 to 1.436) | 0.334 | 0.353 | 1.094 (0.849 to 1.408) | 0.488 | 0.523 |
| Platelet indices | P-LCR | % | Per 1-unit increase | 1.021 (0.989 to 1.053) | 0.198 | 0.228 | 1.020 (0.987 to 1.054) | 0.241 | 0.278 |

| | | | | | | | | | | |
|---|---|---|---|---|---|---|---|---|---|---|
| Glucose and lipid metabolism | GLU | mmol/L | Per increase | 2-fold | 12.477 (4.177 to 37.273) | <0.001 | <0.001 | 14.392 (4.552 to 45.506) | <0.001 | <0.001 |
| Glucose and lipid metabolism | HDL-C | mmol/L | Per increase | 1-unit | 0.084 (0.037 to 0.192) | <0.001 | <0.001 | 0.075 (0.031 to 0.178) | <0.001 | <0.001 |
| Glucose and lipid metabolism | LDL-C | mmol/L | Per increase | 1-unit | 1.575 (1.191 to 2.081) | 0.001 | 0.002 | 1.522 (1.140 to 2.032) | 0.004 | 0.007 |
| Glucose and lipid metabolism | TC | mmol/L | Per increase | 1-unit | 1.250 (0.985 to 1.587) | 0.066 | 0.083 | 1.219 (0.953 to 1.560) | 0.115 | 0.144 |
| Glucose and lipid metabolism | TG | mmol/L | Per increase | 2-fold | 4.398 (2.810 to 6.883) | <0.001 | <0.001 | 4.660 (2.936 to 7.397) | <0.001 | <0.001 |
| Hepatobiliary and liver function | ALP | U/L | Per increase | 2-fold | 8.042 (3.870 to 16.709) | <0.001 | <0.001 | 8.675 (4.052 to 18.573) | <0.001 | <0.001 |
| Hepatobiliary and liver function | ALT | U/L | Per increase | 2-fold | 1.765 (1.260 to 2.471) | <0.001 | 0.002 | 1.939 (1.354 to 2.778) | <0.001 | <0.001 |
| Hepatobiliary and liver function | AST | U/L | Per increase | 2-fold | 3.736 (2.106 to 6.627) | <0.001 | <0.001 | 4.107 (2.262 to 7.456) | <0.001 | <0.001 |
| Hepatobiliary and liver function | DBIL | umol/L | Per increase | 2-fold | 0.852 (0.545 to 1.331) | 0.482 | 0.482 | 0.912 (0.576 to 1.444) | 0.693 | 0.717 |
| Hepatobiliary and liver function | GGT | U/L | Per increase | 2-fold | 2.639 (1.927 to 3.614) | <0.001 | <0.001 | 2.595 (1.888 to 3.566) | <0.001 | <0.001 |
| Hepatobiliary and liver function | LDH | U/L | Per increase | 2-fold | 2.863 (1.365 to 6.001) | 0.005 | 0.008 | 3.157 (1.418 to 7.026) | 0.005 | 0.007 |
| Hepatobiliary and liver function | TBIL | umol/L | Per increase | 2-fold | 0.840 (0.587 to 1.203) | 0.341 | 0.353 | 0.935 (0.645 to 1.355) | 0.722 | 0.722 |
| Protein nutrition and hepatic synthesis | ALB | g/L | Per increase | 1-unit | 0.924 (0.852 to 1.001) | 0.053 | 0.072 | 0.923 (0.849 to 1.003) | 0.059 | 0.081 |

| | | | | | | | | | | |
|---|---|---|---|---|---|---|---|---|---|---|
| Protein nutrition and hepatic synthesis | GLOB | g/L | Per increase | 1-unit | 1.161 (1.087 to 1.239) | <0.001 | <0.001 | 1.164 (1.087 to 1.246) | <0.001 | <0.001 |
| Protein nutrition and hepatic synthesis | TP | g/L | Per increase | 1-unit | 1.026 (0.979 to 1.075) | 0.280 | 0.311 | 1.025 (0.976 to 1.076) | 0.320 | 0.356 |
| Tumor markers | AFP | ng/mL | Per increase | 2-fold | 1.297 (0.982 to 1.713) | 0.066 | 0.083 | 1.280 (0.959 to 1.709) | 0.093 | 0.122 |
| Tumor markers | CA125 | U/mL | Per increase | 2-fold | 2.600 (1.970 to 3.432) | <0.001 | <0.001 | 2.541 (1.927 to 3.352) | <0.001 | <0.001 |
| Tumor markers | CA15-3 | U/mL | Per increase | 2-fold | 7.479 (4.456 to 12.550) | <0.001 | <0.001 | 7.845 (4.588 to 13.415) | <0.001 | <0.001 |
| Tumor markers | CEA | ng/mL | Per increase | 2-fold | 2.075 (1.547 to 2.784) | <0.001 | <0.001 | 2.152 (1.574 to 2.942) | <0.001 | <0.001 |

*Note:P values and FDR q values are reported as <0.001 when below 0.001; otherwise they are shown to three decimal places.*

**Supplementary Table S9. Associations of clinical laboratory markers with ovarian cancer case status in dataset2.**

*Note: Logistic regression modeled ovarian cancer case status (case = 1, control = 0).*

| Marker category | Marker | Unit | Effect scale | Primary model OR (95% CI) | P | FDR q value | Sensitivity model OR (95% CI) | P | FDR q value |
|---|---|---|---|---|---|---|---|---|---|
| CBC and leukocyte differential | EOS | *10^9/L | Per 1-unit increase | 0.259 (0.019 to 3.463) | 0.307 | 0.351 | 0.263 (0.020 to 3.516) | 0.313 | 0.357 |
| CBC and leukocyte differential | LYM | *10^9/L | Per 1-unit increase | 0.209 (0.116 to 0.376) | <0.001 | <0.001 | 0.207 (0.115 to 0.374) | <0.001 | <0.001 |
| CBC and leukocyte differential | MON | *10^9/L | Per 1-unit increase | 8.889 (1.877 to 42.089) | 0.006 | 0.010 | 8.894 (1.877 to 42.145) | 0.006 | 0.010 |
| CBC and leukocyte differential | NEU | *10^9/L | Per 1-unit increase | 1.225 (1.071 to 1.401) | 0.003 | 0.007 | 1.228 (1.072 to 1.406) | 0.003 | 0.007 |
| CBC and leukocyte differential | WBC | *10^9/L | Per 1-unit increase | 1.120 (1.020 to 1.229) | 0.017 | 0.026 | 1.120 (1.020 to 1.230) | 0.018 | 0.027 |
| Erythrocyte and hemoglobin indices | HGB | g/L | Per 1-unit increase | 0.927 (0.904 to 0.951) | <0.001 | <0.001 | 0.927 (0.904 to 0.951) | <0.001 | <0.001 |
| Erythrocyte and hemoglobin indices | MCH | pg | Per 1-unit increase | 1.201 (1.058 to 1.362) | 0.005 | 0.009 | 1.202 (1.059 to 1.364) | 0.004 | 0.009 |
| Erythrocyte and hemoglobin indices | MCHC | g/L | Per 1-unit increase | 1.031 (1.004 to 1.059) | 0.024 | 0.035 | 1.031 (1.004 to 1.059) | 0.024 | 0.034 |
| Erythrocyte and hemoglobin indices | MCV | fL | Per 1-unit increase | 1.071 (1.021 to 1.123) | 0.005 | 0.009 | 1.071 (1.021 to 1.123) | 0.005 | 0.009 |
| Erythrocyte and hemoglobin indices | RDW-SD | fL | Per 1-unit increase | 1.225 (1.135 to 1.321) | <0.001 | <0.001 | 1.228 (1.138 to 1.325) | <0.001 | <0.001 |
| Platelet indices | MPV | fL | Per 1-unit increase | 1.232 (0.972 to 1.560) | 0.084 | 0.104 | 1.233 (0.972 to 1.563) | 0.084 | 0.104 |

| | | | | | | | | | |
|---|---|---|---|---|---|---|---|---|---|
| Platelet indices | P-LCR | % | Per 1-unit increase | 1.035 (1.002 to 1.070) | 0.038 | 0.052 | 1.035 (1.002 to 1.070) | 0.037 | 0.052 |
| Glucose and lipid metabolism | GLU | mmol/L | Per 2-fold increase | 17.861 (4.415 to 72.266) | <0.001 | <0.001 | 22.177 (5.126 to 95.948) | <0.001 | <0.001 |
| Glucose and lipid metabolism | HDL-C | mmol/L | Per 1-unit increase | 0.147 (0.063 to 0.343) | <0.001 | <0.001 | 0.146 (0.063 to 0.342) | <0.001 | <0.001 |
| Glucose and lipid metabolism | LDL-C | mmol/L | Per 1-unit increase | 1.550 (1.097 to 2.192) | 0.013 | 0.021 | 1.565 (1.104 to 2.218) | 0.012 | 0.019 |
| Glucose and lipid metabolism | TC | mmol/L | Per 1-unit increase | 1.076 (0.821 to 1.411) | 0.593 | 0.593 | 1.080 (0.822 to 1.420) | 0.580 | 0.580 |
| Glucose and lipid metabolism | TG | mmol/L | Per 2-fold increase | 3.402 (2.133 to 5.424) | <0.001 | <0.001 | 3.403 (2.134 to 5.428) | <0.001 | <0.001 |
| Hepatobiliary and liver function | ALP | U/L | Per 2-fold increase | 8.609 (3.696 to 20.055) | <0.001 | <0.001 | 8.610 (3.696 to 20.055) | <0.001 | <0.001 |
| Hepatobiliary and liver function | ALT | U/L | Per 2-fold increase | 1.846 (1.246 to 2.734) | 0.002 | 0.005 | 1.857 (1.250 to 2.759) | 0.002 | 0.005 |
| Hepatobiliary and liver function | AST | U/L | Per 2-fold increase | 3.650 (1.816 to 7.337) | <0.001 | <0.001 | 3.697 (1.828 to 7.475) | <0.001 | <0.001 |
| Hepatobiliary and liver function | DBIL | umol/L | Per 2-fold increase | 0.792 (0.493 to 1.275) | 0.338 | 0.372 | 0.786 (0.487 to 1.271) | 0.327 | 0.361 |
| Hepatobiliary and liver function | GGT | U/L | Per 2-fold increase | 2.824 (1.922 to 4.149) | <0.001 | <0.001 | 2.829 (1.925 to 4.159) | <0.001 | <0.001 |
| Hepatobiliary and liver function | LDH | U/L | Per 2-fold increase | 1.276 (0.577 to 2.819) | 0.547 | 0.565 | 1.277 (0.578 to 2.820) | 0.546 | 0.563 |
| Protein nutrition and hepatic synthesis | ALB | g/L | Per 1-unit increase | 0.934 (0.870 to 1.003) | 0.060 | 0.080 | 0.934 (0.870 to 1.003) | 0.060 | 0.081 |

| | | | | | | | | | |
|---|---|---|---|---|---|---|---|---|---|
| Protein nutrition and hepatic synthesis | GLOB | g/L | Per 1-unit increase | 1.102 (1.029 to 1.180) | 0.005 | 0.010 | 1.102 (1.030 to 1.180) | 0.005 | 0.010 |
| Protein nutrition and hepatic synthesis | TP | g/L | Per 1-unit increase | 1.027 (0.981 to 1.075) | 0.262 | 0.310 | 1.027 (0.981 to 1.075) | 0.262 | 0.311 |
| Tumor markers | AFP | ng/mL | Per 2-fold increase | 1.697 (1.154 to 2.496) | 0.007 | 0.012 | 1.699 (1.155 to 2.499) | 0.007 | 0.012 |
| Tumor markers | CA125 | U/mL | Per 2-fold increase | 2.990 (2.093 to 4.271) | <0.001 | <0.001 | 2.990 (2.093 to 4.273) | <0.001 | <0.001 |
| Tumor markers | CA15-3 | U/mL | Per 2-fold increase | 5.751 (3.409 to 9.702) | <0.001 | <0.001 | 5.747 (3.408 to 9.692) | <0.001 | <0.001 |
| Tumor markers | CEA | ng/mL | Per 2-fold increase | 1.940 (1.387 to 2.713) | <0.001 | <0.001 | 1.957 (1.397 to 2.743) | <0.001 | <0.001 |
| Other clinical markers | UA | umol/L | Per 2-fold increase | 1.350 (0.614 to 2.967) | 0.455 | 0.486 | 1.351 (0.615 to 2.970) | 0.454 | 0.484 |
| Other clinical markers | Urea | mmol/L | Per 2-fold increase | 1.819 (0.928 to 3.563) | 0.081 | 0.104 | 1.830 (0.929 to 3.605) | 0.081 | 0.103 |

*Note: P values and FDR q values are reported as <0.001 when below 0.001; otherwise they are shown to three decimal places.*

**Supplementary Table S10. PFAS mixture set definitions.**

| Mixture set | No. of PFAS | PFAS included |
|---|---|---|
| All individual PFAS | 20 | DONA, 6:2CI−PFESA, 8:2CI−PFESA, FBSA, 6:2FTSA, GenX, PFBS, PFDA, PFDoA, PFDS, PFHpA, PFHpS, PFHxS, PFNA, PFNS, PFOA, PFOS, PFPeS, PFTrDA, PFUdA |
| PFCA mixture | 7 | PFHpA, PFOA, PFNA, PFDA, PFUdA, PFDoA, PFTrDA |
| PFSA mixture | 7 | PFBS, PFPeS, PFHxS, PFHpS, PFOS, PFNS, PFDS |
| Alternative PFAS mixture | 6 | DONA, GenX, 6:2CI−PFESA, 8:2CI−PFESA, FBSA, 6:2FTSA |
| Legacy PFAS mixture | 14 | PFOA, PFOS, PFHxS, PFNA, PFDA, PFHpA, PFHpS, PFUdA, PFDoA, PFTrDA, PFBS, PFPeS, PFDS, PFNS |
| Emerging alternative PFAS mixture | 5 | GenX, DONA, 6:2CI−PFESA, 8:2CI−PFESA, 6:2FTSA |
| Disease-priority PFAS mixture | 6 | DONA, GenX, PFPeS, PFNA, PFUdA, PFTrDA |

**Supplementary Table S11. Associations between PFAS group burdens and ovarian cancer case status.**

*Note: Logistic regression modeled ovarian cancer case status (case = 1, control = 0).*

| Exposure group | Model | OR (95% CI) | P | FDR q value |
|---|---|---|---|---|
| All individual PFAS | Model 1 | 2.033 (1.523 to 2.715) | <0.001 | <0.001 |
| All individual PFAS | Model 2 | 2.010 (1.503 to 2.687) | <0.001 | <0.001 |
| All individual PFAS | Model 3 | 2.000 (1.496 to 2.674) | <0.001 | <0.001 |
| All individual PFAS | Model 4 | 1.932 (1.430 to 2.611) | <0.001 | <0.001 |
| PFCA | Model 1 | 1.828 (1.382 to 2.416) | <0.001 | <0.001 |
| PFCA | Model 2 | 1.804 (1.362 to 2.389) | <0.001 | <0.001 |
| PFCA | Model 3 | 1.800 (1.359 to 2.383) | <0.001 | <0.001 |
| PFCA | Model 4 | 1.754 (1.309 to 2.350) | <0.001 | <0.001 |
| PFSA | Model 1 | 1.513 (1.164 to 1.967) | 0.002 | 0.002 |
| PFSA | Model 2 | 1.488 (1.142 to 1.939) | 0.003 | 0.003 |
| PFSA | Model 3 | 1.479 (1.135 to 1.927) | 0.004 | 0.004 |
| PFSA | Model 4 | 1.416 (1.074 to 1.866) | 0.014 | 0.014 |
| Alternative PFAS | Model 1 | 2.664 (1.936 to 3.666) | <0.001 | <0.001 |
| Alternative PFAS | Model 2 | 2.690 (1.951 to 3.708) | <0.001 | <0.001 |
| Alternative PFAS | Model 3 | 2.675 (1.940 to 3.687) | <0.001 | <0.001 |
| Alternative PFAS | Model 4 | 2.564 (1.844 to 3.566) | <0.001 | <0.001 |
| Legacy PFAS | Model 1 | 1.711 (1.302 to 2.248) | <0.001 | <0.001 |
| Legacy PFAS | Model 2 | 1.687 (1.281 to 2.221) | <0.001 | <0.001 |
| Legacy PFAS | Model 3 | 1.680 (1.276 to 2.212) | <0.001 | <0.001 |
| Legacy PFAS | Model 4 | 1.624 (1.220 to 2.163) | <0.001 | 0.001 |
| Emerging alternative PFAS | Model 1 | 2.473 (1.818 to 3.365) | <0.001 | <0.001 |
| Emerging alternative PFAS | Model 2 | 2.458 (1.806 to 3.346) | <0.001 | <0.001 |

| | | | | |
|---|---|---|---|---|
| Emerging alternative PFAS | Model 3 | 2.442 (1.794 to 3.326) | <0.001 | <0.001 |
| Emerging alternative PFAS | Model 4 | 2.278 (1.663 to 3.119) | <0.001 | <0.001 |

*Note: P values and FDR q values are reported as <0.001 when below 0.001; otherwise they are shown to three decimal places.*

**Supplementary Table S12. Associations between individual PFAS and ovarian cancer case status.**

*Note: Logistic regression modeled ovarian cancer case status (case = 1, control = 0).*

| PFAS class | PFAS | Model | OR (95% CI) | P | FDR q value |
|---|---|---|---|---|---|
| PFCA | PFDA | Model 1 | 2.241 (1.653 to 3.039) | <0.001 | <0.001 |
| PFCA | PFDA | Model 2 | 2.310 (1.697 to 3.143) | <0.001 | <0.001 |
| PFCA | PFDA | Model 3 | 2.302 (1.692 to 3.133) | <0.001 | <0.001 |
| PFCA | PFDA | Model 4 | 2.188 (1.595 to 3.000) | <0.001 | <0.001 |
| PFCA | PFDoA | Model 1 | 1.570 (1.189 to 2.074) | 0.001 | 0.003 |
| PFCA | PFDoA | Model 2 | 1.558 (1.179 to 2.058) | 0.002 | 0.003 |
| PFCA | PFDoA | Model 3 | 1.577 (1.190 to 2.090) | 0.002 | 0.003 |
| PFCA | PFDoA | Model 4 | 1.640 (1.216 to 2.212) | 0.001 | 0.002 |
| PFCA | PFHpA | Model 1 | 1.465 (1.126 to 1.905) | 0.004 | 0.007 |
| PFCA | PFHpA | Model 2 | 1.444 (1.107 to 1.882) | 0.007 | 0.010 |
| PFCA | PFHpA | Model 3 | 1.434 (1.100 to 1.869) | 0.008 | 0.012 |
| PFCA | PFHpA | Model 4 | 1.520 (1.138 to 2.031) | 0.005 | 0.008 |
| PFCA | PFNA | Model 1 | 3.632 (2.502 to 5.272) | <0.001 | <0.001 |
| PFCA | PFNA | Model 2 | 3.606 (2.480 to 5.241) | <0.001 | <0.001 |
| PFCA | PFNA | Model 3 | 3.586 (2.466 to 5.214) | <0.001 | <0.001 |
| PFCA | PFNA | Model 4 | 3.691 (2.495 to 5.462) | <0.001 | <0.001 |
| PFCA | PFOA | Model 1 | 1.362 (1.054 to 1.759) | 0.018 | 0.026 |
| PFCA | PFOA | Model 2 | 1.329 (1.020 to 1.733) | 0.035 | 0.047 |
| PFCA | PFOA | Model 3 | 1.328 (1.019 to 1.730) | 0.036 | 0.045 |
| PFCA | PFOA | Model 4 | 1.319 (0.998 to 1.744) | 0.052 | 0.065 |
| PFCA | PFTrDA | Model 1 | 2.615 (1.868 to 3.660) | <0.001 | <0.001 |
| PFCA | PFTrDA | Model 2 | 2.762 (1.943 to 3.927) | <0.001 | <0.001 |
| PFCA | PFTrDA | Model 3 | 2.778 (1.952 to 3.953) | <0.001 | <0.001 |

| | | | | | |
|---|---|---|---|---|---|
| PFCA | PFTrDA | Model 4 | 2.558 (1.795 to 3.644) | <0.001 | <0.001 |
| PFCA | PFUdA | Model 1 | 3.977 (2.360 to 6.702) | <0.001 | <0.001 |
| PFCA | PFUdA | Model 2 | 5.248 (2.904 to 9.484) | <0.001 | <0.001 |
| PFCA | PFUdA | Model 3 | 5.221 (2.890 to 9.429) | <0.001 | <0.001 |
| PFCA | PFUdA | Model 4 | 4.812 (2.663 to 8.696) | <0.001 | <0.001 |
| PFSA | PFBS | Model 1 | 2.322 (1.700 to 3.172) | <0.001 | <0.001 |
| PFSA | PFBS | Model 2 | 2.338 (1.708 to 3.199) | <0.001 | <0.001 |
| PFSA | PFBS | Model 3 | 2.326 (1.698 to 3.185) | <0.001 | <0.001 |
| PFSA | PFBS | Model 4 | 2.257 (1.641 to 3.105) | <0.001 | <0.001 |
| PFSA | PFDS | Model 1 | 1.284 (1.000 to 1.649) | 0.050 | 0.062 |
| PFSA | PFDS | Model 2 | 1.282 (0.997 to 1.647) | 0.053 | 0.062 |
| PFSA | PFDS | Model 3 | 1.299 (1.009 to 1.672) | 0.043 | 0.050 |
| PFSA | PFDS | Model 4 | 1.300 (0.999 to 1.693) | 0.051 | 0.065 |
| PFSA | PFHpS | Model 1 | 1.708 (1.304 to 2.238) | <0.001 | <0.001 |
| PFSA | PFHpS | Model 2 | 1.689 (1.282 to 2.225) | <0.001 | <0.001 |
| PFSA | PFHpS | Model 3 | 1.684 (1.279 to 2.218) | <0.001 | <0.001 |
| PFSA | PFHpS | Model 4 | 1.692 (1.276 to 2.243) | <0.001 | <0.001 |
| PFSA | PFHxS | Model 1 | 0.931 (0.728 to 1.191) | 0.569 | 0.576 |
| PFSA | PFHxS | Model 2 | 0.850 (0.647 to 1.117) | 0.244 | 0.271 |
| PFSA | PFHxS | Model 3 | 0.850 (0.647 to 1.117) | 0.243 | 0.270 |
| PFSA | PFHxS | Model 4 | 0.839 (0.629 to 1.119) | 0.232 | 0.258 |
| PFSA | PFNS | Model 1 | 0.929 (0.726 to 1.188) | 0.556 | 0.576 |
| PFSA | PFNS | Model 2 | 0.941 (0.735 to 1.206) | 0.632 | 0.632 |
| PFSA | PFNS | Model 3 | 0.945 (0.738 to 1.211) | 0.654 | 0.654 |
| PFSA | PFNS | Model 4 | 0.932 (0.719 to 1.208) | 0.592 | 0.624 |
| PFSA | PFOS | Model 1 | 1.562 (1.198 to 2.035) | <0.001 | 0.002 |

| | | | | | |
|---|---|---|---|---|---|
| PFSA | PFOS | Model 2 | 1.544 (1.184 to 2.012) | 0.001 | 0.003 |
| PFSA | PFOS | Model 3 | 1.534 (1.176 to 2.000) | 0.002 | 0.003 |
| PFSA | PFOS | Model 4 | 1.464 (1.109 to 1.932) | 0.007 | 0.012 |
| PFSA | PFPeS | Model 1 | 2.898 (2.106 to 3.988) | <0.001 | <0.001 |
| PFSA | PFPeS | Model 2 | 2.923 (2.119 to 4.034) | <0.001 | <0.001 |
| PFSA | PFPeS | Model 3 | 2.927 (2.117 to 4.046) | <0.001 | <0.001 |
| PFSA | PFPeS | Model 4 | 2.931 (2.093 to 4.104) | <0.001 | <0.001 |
| Alternative PFAS | DONA | Model 1 | 7.979 (5.167 to 12.322) | <0.001 | <0.001 |
| Alternative PFAS | DONA | Model 2 | 8.594 (5.428 to 13.606) | <0.001 | <0.001 |
| Alternative PFAS | DONA | Model 3 | 8.571 (5.414 to 13.569) | <0.001 | <0.001 |
| Alternative PFAS | DONA | Model 4 | 9.881 (5.896 to 16.560) | <0.001 | <0.001 |
| Alternative PFAS | 6:2CI−PFESA | Model 1 | 1.471 (1.134 to 1.907) | 0.004 | 0.006 |
| Alternative PFAS | 6:2CI−PFESA | Model 2 | 1.451 (1.118 to 1.884) | 0.005 | 0.009 |
| Alternative PFAS | 6:2CI−PFESA | Model 3 | 1.442 (1.110 to 1.872) | 0.006 | 0.010 |
| Alternative PFAS | 6:2CI−PFESA | Model 4 | 1.372 (1.044 to 1.804) | 0.023 | 0.036 |
| Alternative PFAS | 8:2CI−PFESA | Model 1 | 1.290 (1.004 to 1.657) | 0.046 | 0.062 |
| Alternative PFAS | 8:2CI−PFESA | Model 2 | 1.346 (1.041 to 1.740) | 0.023 | 0.033 |

| | | | | | |
|---|---|---|---|---|---|
| Alternative PFAS | 8:2CI−PFESA | Model 3 | 1.341 (1.037 to 1.734) | 0.025 | 0.036 |
| Alternative PFAS | 8:2CI−PFESA | Model 4 | 1.267 (0.970 to 1.654) | 0.082 | 0.097 |
| Alternative PFAS | FBSA | Model 1 | 1.272 (0.991 to 1.634) | 0.059 | 0.070 |
| Alternative PFAS | FBSA | Model 2 | 1.310 (1.015 to 1.691) | 0.038 | 0.048 |
| Alternative PFAS | FBSA | Model 3 | 1.319 (1.021 to 1.704) | 0.034 | 0.045 |
| Alternative PFAS | FBSA | Model 4 | 1.307 (0.999 to 1.710) | 0.051 | 0.065 |
| Alternative PFAS | 6:2FTSA | Model 1 | 1.073 (0.839 to 1.372) | 0.576 | 0.576 |
| Alternative PFAS | 6:2FTSA | Model 2 | 1.086 (0.848 to 1.391) | 0.514 | 0.541 |
| Alternative PFAS | 6:2FTSA | Model 3 | 1.077 (0.840 to 1.380) | 0.560 | 0.589 |
| Alternative PFAS | 6:2FTSA | Model 4 | 1.011 (0.778 to 1.313) | 0.937 | 0.937 |
| Alternative PFAS | GenX | Model 1 | 6.882 (4.261 to 11.116) | <0.001 | <0.001 |
| Alternative PFAS | GenX | Model 2 | 6.844 (4.234 to 11.063) | <0.001 | <0.001 |
| Alternative PFAS | GenX | Model 3 | 6.815 (4.212 to 11.027) | <0.001 | <0.001 |
| Alternative PFAS | GenX | Model 4 | 6.742 (4.112 to 11.053) | <0.001 | <0.001 |

*Note: P values and FDR q values are reported as <0.001 when below 0.001; otherwise they are shown to three decimal places.*

**Supplementary Table S13. Summary of individual PFAS associations by PFAS class in Model 4.**

| PFAS class | PFAS analyzed | Estimable PFAS | FDR-significant PFAS | Nominally significant PFAS | Median OR in Model 4 | Top PFAS by FDR | Top PFAS OR (95% CI) |
|---|---|---|---|---|---|---|---|
| PFCA | 7 | 7 | 6 | 6 | 2.188 | PFNA | 3.691 (2.495 to 5.462) |
| PFSA | 7 | 7 | 4 | 4 | 1.464 | PFPeS | 2.931 (2.093 to 4.104) |
| Alternative PFAS | 6 | 6 | 3 | 3 | 1.339 | DONA | 9.881 (5.896 to 16.560) |

**Supplementary Table S14. Cross-method priority components within the disease-priority PFAS mixture for ovarian cancer case status.**

| PFAS | PFAS class | qgcomp weight | gWQS positive weight | BKMR PIP |
|---|---|---|---|---|
| DONA | Alternative PFAS | 0.420 | 0.396 | 1.000 |
| GenX | Alternative PFAS | 0.076 | 0.035 | 1.000 |
| PFPeS | PFSA | 0.177 | 0.164 | 0.571 |
| PFNA | PFCA | 0.097 | 0.172 | 0.822 |
| PFUdA | PFCA | 0.151 | 0.196 | 0.987 |
| PFTrDA | PFCA | 0.080 | 0.037 | 1.000 |

*Note: The disease-priority mixture included DONA, GenX, PFPeS, PFNA, PFUdA, and PFTrDA. BKMR was fitted as an exploratory binary model with 20,000 iterations and variable selection.*

**Supplementary Table S15. PFAS mixture associations with ovarian cancer case status in qgcomp and gWQS models.**

| Mixture set | No. of PFAS | qgcomp OR (95% CI) | qgcomp P | gWQS positive OR (95% CI) | gWQS positive P | gWQS negative OR (95% CI) | Negative P | Negative fit |
|---|---|---|---|---|---|---|---|---|
| All individual PFAS | 20 | 34.12 (5.39, 215.96) | <0.001 | 43.12 (11.92, 155.92) | <0.001 | | | failed |
| PFCA mixture | 7 | 4.35 (2.47, 7.66) | <0.001 | 4.28 (2.40, 7.64) | <0.001 | | | failed |
| PFSA mixture | 7 | 3.91 (2.19, 6.97) | <0.001 | 4.84 (2.64, 8.88) | <0.001 | 0.79 (0.51, 1.22) | 0.295 | warning |
| Alternative PFAS mixture | 6 | 9.82 (4.46, 21.63) | <0.001 | 13.90 (6.01, 32.16) | <0.001 | | | failed |
| Legacy PFAS mixture | 14 | 9.28 (3.63, 23.72) | <0.001 | 11.33 (4.84, 26.55) | <0.001 | | | failed |
| Emerging alternative PFAS mixture | 5 | 10.38 (4.86, 22.19) | <0.001 | 13.64 (5.90, 31.54) | <0.001 | | | failed |
| Disease-priority PFAS mixture | 6 | 68.11 (22.14, 209.53) | <0.001 | 32.27 (10.17, 102.35) | <0.001 | | | failed |

*Note: ORs are estimated per simultaneous one-quantile increase in the mixture index. q=4; n=256 (128 cases and 128 controls).*

*Note: gWQS models used bootstrap=100 and validation=0.6.*

*Note: Failed gWQS negative-direction fits indicate that the specified direction was not supported among bootstrapped models. Warning fits were retained but should be interpreted cautiously.*

**Supplementary Table S16. Component weights in the all-individual PFAS mixture for ovarian cancer case status.**

| PFAS | PFAS class | qgcomp direction | qgcomp signed weight | gWQS positive weight |
|---|---|---|---|---|
| DONA | Alternative PFAS | Positive | 0.255 | 0.355 |
| GenX | Alternative PFAS | Positive | 0.043 | 0.008 |
| PFNA | PFCA | Positive | 0.190 | 0.086 |
| PFUdA | PFCA | Positive | 0.125 | 0.157 |
| PFPeS | PFSA | Positive | 0.095 | 0.155 |
| PFHpS | PFSA | Positive | 0.083 | 0.016 |
| PFTrDA | PFCA | Positive | 0.074 | 0.011 |
| PFBS | PFSA | Positive | 0.065 | 0.065 |
| 6:2CI−PFESA | Alternative PFAS | Positive | 0.059 | 0.006 |
| PFDS | PFSA | Positive | 0.011 | 0.004 |
| PFHxS | PFSA | Negative | -0.200 | 0.000 |
| PFOA | PFCA | Negative | -0.137 | 0.000 |
| PFDA | PFCA | Negative | -0.135 | 0.034 |
| PFHpA | PFCA | Negative | -0.128 | 0.000 |
| FBSA | Alternative PFAS | Negative | -0.105 | 0.005 |
| PFOS | PFSA | Negative | -0.098 | 0.000 |
| PFDoA | PFCA | Negative | -0.091 | 0.001 |
| 8:2CI−PFESA | Alternative PFAS | Negative | -0.056 | 0.025 |
| PFNS | PFSA | Negative | -0.032 | 0.058 |
| 6:2FTSA | Alternative PFAS | Negative | -0.017 | 0.015 |

*Note: qgcomp positive and negative weights each sum to 1 within direction; signed weights are shown only to distinguish direction. gWQS values are positive-direction mean WQS weights.*

Supplementary Table S17. Associations of individual PFAS with ovarian cancer case status and lipid-association burden in dataset1.

| PFAS | PFAS class | Ovarian cancer OR (95% CI) | Ovarian cancer FDR | FDR-associated lipids | Negative assoc. | Positive assoc. | Disease-differential and associated | Focus-class associated lipids | Bridge candidates | Pattern-consistent bridges | Strongest lipid | Lipid class | Strongest beta | Strongest FDR |
|---|---|---|---|---|---|---|---|---|---|---|---|---|---|---|
| DONA | Alternative PFAS | 9.881 (5.896 to 16.560) | 6.99E-17 | 513 | 402 | 111 | 325 | 407 | 258 | 258 | LPC(O-16:1) | LPC | -0.516 | 5.81E-15 |
| 6:2CI−PFESA | Alternative PFAS | 1.372 (1.044 to 1.804) | 3.60E-02 | 0 | 0 | 0 | 0 | 0 | 0 | 0 | PC(P-43:13) | PC | 0.232 | 1.87E-01 |
| 8:2CI−PFESA | Alternative PFAS | 1.267 (0.970 to 1.654) | 9.67E-02 | 2 | 1 | 1 | 0 | 1 | 0 | 0 | PI(16:0_18:3) | PI | 0.285 | 1.26E-02 |
| FBSA | Alternative PFAS | 1.307 (0.999 to 1.710) | 6.48E-02 | 0 | 0 | 0 | 0 | 0 | 0 | 0 | PE(18:0_22:2CHO) | PE | 0.244 | 1.90E-01 |

| | | | | | | | | | | | | | | |
|---|---|---|---|---|---|---|---|---|---|---|---|---|---|---|
| 6:2FTSA | Alternative PFAS | 1.011 (0.778 to 1.313) | 9.37E-01 | 0 | 0 | 0 | 0 | 0 | 0 | 0 | TG(10:0COOH_16:0_18:0) | TG | -0.177 | 8.75E-01 |
| GenX | Alternative PFAS | 6.742 (4.112 to 11.053) | 3.84E-13 | 242 | 188 | 54 | 209 | 191 | 165 | 165 | LPE(P-18:0) | LPE | -0.452 | 1.03E-10 |
| PFBS | PFSA | 2.257 (1.641 to 3.105) | 1.60E-06 | 0 | 0 | 0 | 0 | 0 | 0 | 0 | LPC(17:1) | LPC | -0.259 | 5.50E-02 |
| PFDA | PFCA | 2.188 (1.595 to 3.000) | 2.97E-06 | 1 | 1 | 0 | 1 | 1 | 0 | 0 | PE(O-20:1_18:2) | PE | -0.277 | 1.44E-02 |
| PFDoA | PFCA | 1.640 (1.216 to 2.212) | 2.36E-03 | 0 | 0 | 0 | 0 | 0 | 0 | 0 | SPH(d15:1) | SPH | -0.236 | 1.13E-01 |
| PFDS | PFSA | 1.300 (0.999 to 1.693) | 6.48E-02 | 0 | 0 | 0 | 0 | 0 | 0 | 0 | AcCa(18:2) | AcCa | -0.221 | 6.44E-01 |

| | | | | | | | | | | | | | | |
|---|---|---|---|---|---|---|---|---|---|---|---|---|---|---|
| PFHpA | PFCA | 1.520 (1.138 to 2.031) | 8.35E-03 | 0 | 0 | 0 | 0 | 0 | 0 | 0 | Cer(m18:2_16:0) | Cer | -0.233 | 1.83E-01 |
| PFHpS | PFSA | 1.692 (1.276 to 2.243) | 5.77E-04 | 4 | 2 | 2 | 3 | 2 | 0 | 0 | PG(18:2_20:5) | PG | 0.292 | 6.58E-03 |
| PFHxS | PFSA | 0.839 (0.629 to 1.119) | 2.58E-01 | 0 | 0 | 0 | 0 | 0 | 0 | 0 | PC(7:2COOH_30:1) | PC | 0.261 | 1.89E-01 |
| PFNA | PFCA | 3.691 (2.495 to 5.462) | 4.31E-10 | 120 | 97 | 23 | 96 | 103 | 83 | 83 | LPE(P-18:1) | LPE | -0.310 | 3.72E-04 |
| PFNS | PFSA | 0.932 (0.719 to 1.208) | 6.24E-01 | 0 | 0 | 0 | 0 | 0 | 0 | 0 | PE(22:5_19:5CHO) | PE | -0.178 | 9.99E-01 |
| PFOA | PFCA | 1.319 (0.998 to 1.744) | 6.48E-02 | 0 | 0 | 0 | 0 | 0 | 0 | 0 | AcCa(20:5) | AcCa | -0.223 | 5.40E-01 |
| PFOS | PFSA | 1.464 (1.109 | 1.19E-02 | 0 | 0 | 0 | 0 | 0 | 0 | 0 | PC(P-15:2_20:4) | PC | -0.217 | 4.98E-01 |

| | | | | | | | | | | | | | | |
|---|---|---|---|---|---|---|---|---|---|---|---|---|---|---|
| | | to 1.932) | | | | | | | | | | | | |
| PFPeS | PFSA | 2.931 (2.093 to 4.104) | 1.94E-09 | 95 | 83 | 12 | 83 | 81 | 71 | 71 | LPC(O-16:1) | LPC | -0.313 | 2.22E-04 |
| PFTrDA | PFCA | 2.558 (1.795 to 3.644) | 6.66E-07 | 43 | 38 | 5 | 39 | 35 | 0 | 0 | FA(22:4) | FA | 0.312 | 1.57E-03 |
| PFUdA | PFCA | 4.812 (2.663 to 8.696) | 6.66E-07 | 54 | 53 | 1 | 50 | 46 | 0 | 0 | LPC(O-16:1) | LPC | -0.293 | 5.76E-03 |

*Note: Logistic regression modeled ovarian cancer case status (case = 1, control = 0). Lipid-association counts are from Model A.*

*Model A: Covariate-adjusted PFAS-lipid association: lipid ~ PFAS + Age + BMI + Hypertension*

**Supplementary Table S18. Lipid-class summary of associations for disease-priority PFAS in dataset1.**

| PFAS | Lipid class | Lipids tested | Disease-differential lipids | FDR-associated lipids | Negative assoc. | Positive assoc. | Bridge candidates | Pattern-consistent bridges | Median beta | Minimum FDR | Strongest lipid | Strongest beta | Dominant direction |
|---|---|---|---|---|---|---|---|---|---|---|---|---|---|
| DONA | LPC | 62 | 47 | 46 | 46 | 0 | 44 | 44 | -0.331 | 5.81E-15 | LPC(O-16:1) | -0.516 | negative |
| DONA | PC | 573 | 161 | 164 | 144 | 20 | 93 | 93 | -0.075 | 1.19E-13 | PC(P-5:0_15:1) | -0.487 | negative |
| DONA | PE | 143 | 79 | 59 | 54 | 5 | 52 | 52 | -0.096 | 4.23E-06 | PE(7:2COOH_32:1) | -0.328 | negative |
| DONA | SM | 107 | 46 | 49 | 49 | 0 | 29 | 29 | -0.154 | 4.14E-07 | SM(d16:1_23:0) | -0.357 | negative |
| DONA | TG | 412 | 40 | 48 | 21 | 27 | 13 | 13 | 0.009 | 9.29E-06 | TG(29:4_2:0_4:0) | -0.317 | positive |
| DONA | DG | 158 | 29 | 26 | 19 | 7 | 14 | 14 | -0.031 | 4.56E-07 | DG(3:0_24:5) | -0.357 | negative |
| DONA | AcCa | 28 | 17 | 15 | 14 | 1 | 13 | 13 | -0.161 | 1.98E-09 | AcCa(20:4) | -0.409 | negative |
| GenX | LPC | 62 | 47 | 40 | 40 | 0 | 40 | 40 | -0.225 | 2.12E-08 | LPC(O-18:1) | -0.408 | negative |
| GenX | PC | 573 | 161 | 77 | 68 | 9 | 66 | 66 | -0.049 | 2.29E-06 | PC(P-5:0_15:1) | -0.354 | negative |
| GenX | PE | 143 | 79 | 27 | 24 | 3 | 27 | 27 | -0.104 | 1.09E-03 | PE(P-18:0_22:6) | -0.263 | negative |
| GenX | SM | 107 | 46 | 11 | 11 | 0 | 8 | 8 | -0.075 | 1.26E-03 | SM(d30:1) | -0.263 | negative |
| GenX | TG | 412 | 40 | 13 | 5 | 8 | 5 | 5 | -0.006 | 4.30E-03 | TG(30:4_16:1) | 0.239 | positive |
| GenX | DG | 158 | 29 | 13 | 6 | 7 | 10 | 10 | -0.021 | 7.08E-04 | DG(O-15:1_10:0) | -0.272 | positive |

| | | | | | | | | | | | | | |
|---|---|---|---|---|---|---|---|---|---|---|---|---|---|
| GenX | AcCa | 28 | 17 | 10 | 9 | 1 | 9 | 9 | -0.128 | 1.04E-03 | AcCa(20:5) | -0.265 | negative |
| PFNA | LPC | 62 | 47 | 23 | 23 | 0 | 23 | 23 | -0.152 | 5.44E-04 | LPC(P-16:0) | -0.297 | negative |
| PFNA | PC | 573 | 161 | 39 | 28 | 11 | 29 | 29 | -0.031 | 1.81E-03 | PC(O-15:1_22:5) | -0.271 | negative |
| PFNA | PE | 143 | 79 | 19 | 18 | 1 | 19 | 19 | -0.070 | 8.32E-04 | PE(O-20:1_18:2) | -0.285 | negative |
| PFNA | SM | 107 | 46 | 5 | 5 | 0 | 4 | 4 | -0.057 | 4.12E-03 | SM(t35:5) | -0.248 | negative |
| PFNA | TG | 412 | 40 | 10 | 5 | 5 | 3 | 3 | 0.000 | 1.30E-02 | TG(22:0_16:1_18:1) | 0.225 | mixed |
| PFNA | DG | 158 | 29 | 3 | 3 | 0 | 1 | 1 | -0.029 | 1.81E-03 | DG(3:0_30:0) | -0.274 | negative |
| PFNA | AcCa | 28 | 17 | 4 | 3 | 1 | 4 | 4 | -0.090 | 3.72E-04 | AcCa(20:5) | -0.308 | negative |
| PFPeS | LPC | 62 | 47 | 27 | 27 | 0 | 27 | 27 | -0.179 | 2.22E-04 | LPC(O-16:1) | -0.313 | negative |
| PFPeS | PC | 573 | 161 | 22 | 20 | 2 | 19 | 19 | -0.037 | 2.23E-04 | PC(P-5:0_15:1) | -0.302 | negative |
| PFPeS | PE | 143 | 79 | 8 | 7 | 1 | 8 | 8 | -0.066 | 2.70E-03 | PE(P-18:0_22:5) | -0.257 | negative |
| PFPeS | SM | 107 | 46 | 8 | 7 | 1 | 6 | 6 | -0.085 | 7.72E-03 | SM(d30:0) | -0.238 | negative |
| PFPeS | TG | 412 | 40 | 2 | 1 | 1 | 2 | 2 | 0.012 | 6.40E-04 | TG(29:4_2:0_4:0) | -0.280 | mixed |
| PFPeS | DG | 158 | 29 | 5 | 2 | 3 | 1 | 1 | -0.023 | 8.27E-03 | DG(O-15:1_10:0) | -0.234 | positive |
| PFPeS | AcCa | 28 | 17 | 9 | 9 | 0 | 8 | 8 | -0.136 | 2.22E-04 | AcCa(20:4) | -0.310 | negative |

Supplementary Table S19. Bridge-candidate counts by disease-priority PFAS and lipid class in dataset1.

| PFAS | LPC | PC | PE | SM | TG | DG | AcCa |
|---|---|---|---|---|---|---|---|
| DONA | 44 | 93 | 52 | 29 | 13 | 14 | 13 |
| GenX | 40 | 66 | 27 | 8 | 5 | 10 | 9 |
| PFNA | 23 | 29 | 19 | 4 | 3 | 1 | 4 |
| PFPeS | 27 | 19 | 8 | 6 | 2 | 1 | 8 |

Supplementary Table S20. Median covariate-adjusted PFAS-lipid association beta by disease-priority PFAS and lipid class in dataset1.

| PFAS | LPC | PC | PE | SM | TG | DG | AcCa |
|---|---|---|---|---|---|---|---|
| DONA | -0.331 | -0.075 | -0.096 | -0.154 | 0.009 | -0.031 | -0.161 |
| GenX | -0.225 | -0.049 | -0.104 | -0.075 | -0.006 | -0.021 | -0.128 |
| PFNA | -0.152 | -0.031 | -0.070 | -0.057 | 0.000 | -0.029 | -0.090 |
| PFPeS | -0.179 | -0.037 | -0.066 | -0.085 | 0.012 | -0.023 | -0.136 |

*Note: Model A: Covariate-adjusted PFAS-lipid association: lipid ~ PFAS + Age + BMI + Hypertension*

Supplementary Table S21. Shared small-molecule mediation summary by disease-priority PFAS in dataset1.

| PFAS | Candidate Pairs | Model Ok | Bootstrap FDR Supported | Bootstrap Nominal Supported | Screen FDR Supported | Screen Nominal Supported | Median Indirect Log Odds | Best bootstrap P | Best Mediator |
|---|---|---|---|---|---|---|---|---|---|
| DONA | 5 | 5 | 4.00E+00 | 0 | 0.00E+00 | 0 | 0.225 | 6.64E-03 | Hippuric acid |
| GenX | 5 | 5 | 4.00E+00 | 0 | 0.00E+00 | 0 | 0.144 | 6.64E-03 | Hippuric acid |
| PFNA | 5 | 5 | 2.00E+00 | 0 | 0.00E+00 | 1 | 0.133 | 6.64E-03 | Hippuric acid |
| PFPeS | 5 | 5 | 4.00E+00 | 1 | 0.00E+00 | 0 | 0.141 | 6.64E-03 | Hippuric acid |

*Note: dataset1 denotes the plasma subset; dataset2 denotes the urine subset when both datasets are shown.*

Supplementary Table S22. Shared small-molecule mediation summary by mediator in dataset1.

| Mediator | Priority Tier | Candidate Pairs | Model Ok | Bootstrap FDR Supported | Bootstrap Nominal Supported | Any Supported | Median Indirect Log Odds | Best bootstrap P | Best PFAS |
|---|---|---|---|---|---|---|---|---|---|
| Corticosterone | Tier 2 | 4 | 4 | 3.00E+00 | 0 | 4 | 0.143 | 6.64E-03 | DONA |
| Elaidic acid | Tier 1 | 4 | 4 | 4.00E+00 | 0 | 4 | 0.320 | 6.64E-03 | GenX |
| Hippuric acid | Tier 1 | 4 | 4 | 4.00E+00 | 0 | 4 | 0.285 | 6.64E-03 | GenX |
| Betaine | Tier 2 | 4 | 4 | 2.00E+00 | 0 | 2 | 0.097 | 6.64E-03 | GenX |
| Taurocholic acid | Tier 2 | 4 | 4 | 1.00E+00 | 1 | 2 | 0.076 | 6.64E-03 | DONA |

Supplementary Table S23. Supported PFAS-shared metabolite-ovarian cancer mediation candidates in dataset1.

| PFAS | Mediator | Priority Tier | Indirect Log Odds | Indirect OR (95% CI) | Sobel FDR | Bootstrap p indirect FDR | CI excludes zero | Direct OR (95% CI) | Total OR (95% CI) | Final Priority | dataset 1 Log2FC | dataset 2 Log2FC | dataset 1 Lasso Freq | dataset 2 Lasso Freq |
|---|---|---|---|---|---|---|---|---|---|---|---|---|---|---|
| GenX | Hippuric acid | Tier 1 | 0.337 | 1.432 (1.207 to 1.824) | 6.89E-03 | 1.53E-02 | Yes | 6.529 (3.900 to 12.172) | 7.163 (4.360 to 14.450) | A1: bootstrap FDR | -2.392 | -0.860 | 1.000 | 1 |
| GenX | Elaidic acid | Tier 1 | 0.322 | 1.386 (1.133 to 2.059) | 5.46E-02 | 1.53E-02 | Yes | 6.661 (4.005 to 13.885) | 7.121 (4.292 to 14.768) | A1: bootstrap FDR | 0.953 | 2.310 | 1.000 | 1 |
| DONA | Hippuric acid | Tier 1 | 0.317 | 1.395 (1.146 to 1.780) | 9.25E-03 | 1.53E-02 | Yes | 9.589 (5.798 to 22.176) | 11.156 (6.442 to 22.693) | A1: bootstrap FDR | -2.392 | -0.860 | 1.000 | 1 |
| PFNA | Hippuric acid | Tier 1 | 0.254 | 1.302 (1.129 to 1.545) | 9.25E-03 | 1.53E-02 | Yes | 3.473 (2.214 to 5.709) | 3.925 (2.570 to 6.197) | A1: bootstrap FDR | -2.392 | -0.860 | 1.000 | 1 |

| | | | | | | | | | | | | | | |
|---|---|---|---|---|---|---|---|---|---|---|---|---|---|---|
| DONA | Corticosterone | Tier 2 | 0.225 | 1.253 (1.093 to 1.526) | 2.41E-02 | 1.53E-02 | Yes | 9.837 (6.127 to 21.570) | 9.950 (6.500 to 20.005) | A1: bootstrap FDR | -2.043 | -0.643 | 0.000 | 1 |
| PFPeS | Hippuric acid | Tier 1 | 0.190 | 1.213 (1.058 to 1.456) | 4.05E-02 | 1.53E-02 | Yes | 3.201 (2.334 to 5.148) | 3.053 (2.216 to 4.451) | A1: bootstrap FDR | -2.392 | -0.860 | 1.000 | 1 |
| GenX | Corticosterone | Tier 2 | 0.144 | 1.159 (1.064 to 1.301) | 4.99E-02 | 1.53E-02 | Yes | 6.575 (4.132 to 13.160) | 6.777 (4.288 to 13.969) | A1: bootstrap FDR | -2.043 | -0.643 | 0.000 | 1 |
| DONA | Taurocholic acid | Tier 2 | 0.117 | 1.124 (1.023 to 1.332) | 6.19E-02 | 1.53E-02 | Yes | 11.898 (6.482 to 27.133) | 10.702 (6.385 to 20.862) | A1: bootstrap FDR | 1.664 | 0.623 | 1.000 | 0 |
| GenX | Betaine | Tier 2 | 0.105 | 1.110 (1.029 to 1.271) | 5.49E-02 | 1.53E-02 | Yes | 6.938 (4.316 to 13.733) | 6.987 (4.319 to 14.002) | A1: bootstrap FDR | 1.076 | 0.659 | 0.410 | 0 |

| | | | | | | | | | | | | | | |
|---|---|---|---|---|---|---|---|---|---|---|---|---|---|---|
| PFNA | Elaidic acid | Tier 1 | 0.325 | 1.348 (1.092 to 2.046) | 5.46E-02 | 1.81E-02 | Yes | 3.611 (2.413 to 6.552) | 3.748 (2.633 to 5.949) | A1: bootstrap FDR | 0.953 | 2.310 | 1.000 | 1 |
| PFPeS | Corticosterone | Tier 2 | 0.141 | 1.151 (1.052 to 1.319) | 4.99E-02 | 1.81E-02 | Yes | 2.942 (2.196 to 4.199) | 2.955 (2.241 to 4.380) | A1: bootstrap FDR | -2.043 | -0.643 | 0.000 | 1 |
| PFPeS | Betaine | Tier 2 | 0.101 | 1.103 (1.023 to 1.248) | 5.46E-02 | 1.81E-02 | Yes | 2.922 (2.167 to 4.392) | 3.018 (2.215 to 4.564) | A1: bootstrap FDR | 1.076 | 0.659 | 0.410 | 0 |
| PFPeS | Elaidic acid | Tier 1 | 0.166 | 1.189 (1.042 to 1.631) | 1.21E-01 | 4.33E-02 | Yes | 2.971 (2.190 to 4.518) | 3.004 (2.260 to 4.478) | A1: bootstrap FDR | 0.953 | 2.310 | 1.000 | 1 |
| DONA | Elaidic acid | Tier 1 | 0.318 | 1.382 (1.059 to 2.154) | 5.80E-02 | 4.98E-02 | Yes | 14.754 (8.052 to 29.747) | 10.489 (6.574 to 20.362) | A1: bootstrap FDR | 0.953 | 2.310 | 1.000 | 1 |
| PFPeS | Taurocholic acid | Tier 2 | 0.079 | 1.078 (1.004 to 1.202) | 8.56E-02 | 6.38E-02 | Yes | 3.002 (2.119 to 4.580) | 2.981 (2.156 to 4.290) | A2: bootstrap nominal | 1.664 | 0.623 | 1.000 | 0 |

| | | | | | | | | | | | | | | |
|---|---|---|---|---|---|---|---|---|---|---|---|---|---|---|
| PFNA | Corticosterone | Tier 2 | 0.133 | 1.138 (0.987 to 1.337) | 5.80E-02 | 9.20E-02 | No | 4.026 (2.557 to 6.873) | 3.738 (2.511 to 5.871) | B2: screen nominal | -2.043 | -0.643 | 0.000 | 1 |

Supplementary Table S24. Summary of PFAS-shared metabolite-clinical marker chains in dataset1.

| PFAS | Clinical Marker | N Chains | Primary Supported Chains | Disease Consistent Chains | Median Via Metabolite Effect | Best Chain Score | Top Mediator | Top Priority Tier |
|---|---|---|---|---|---|---|---|---|
| PFPeS | RDW-SD | 5 | 4 | 5 | 0.029 | 10.060 | Hippuric acid | Tier 1 |
| PFPeS | HDL-C | 5 | 4 | 5 | -0.024 | 7.335 | Elaidic acid | Tier 1 |
| DONA | HDL-C | 5 | 4 | 5 | -0.039 | 7.256 | Elaidic acid | Tier 1 |
| DONA | RDW-SD | 5 | 3 | 5 | 0.027 | 10.060 | Hippuric acid | Tier 1 |
| GenX | RDW-SD | 5 | 3 | 5 | 0.034 | 10.060 | Hippuric acid | Tier 1 |
| DONA | TG | 5 | 3 | 5 | 0.040 | 8.765 | Hippuric acid | Tier 1 |
| GenX | TG | 5 | 3 | 5 | 0.028 | 8.765 | Hippuric acid | Tier 1 |
| PFPeS | TG | 5 | 3 | 5 | 0.026 | 8.765 | Hippuric acid | Tier 1 |
| GenX | HDL-C | 5 | 3 | 5 | -0.025 | 8.034 | Elaidic acid | Tier 1 |
| PFNA | RDW-SD | 5 | 2 | 5 | 0.020 | 10.060 | Hippuric acid | Tier 1 |
| PFNA | TG | 5 | 2 | 5 | 0.022 | 8.765 | Hippuric acid | Tier 1 |
| GenX | GGT | 5 | 2 | 5 | 0.028 | 7.983 | Hippuric acid | Tier 1 |
| PFPeS | GGT | 5 | 2 | 5 | 0.015 | 7.983 | Hippuric acid | Tier 1 |

| | | | | | | | | |
|---|---|---|---|---|---|---|---|---|
| DONA | GLU | 5 | 2 | 5 | 0.020 | 7.979 | Hippuric acid | Tier 1 |
| GenX | GLU | 5 | 2 | 5 | 0.024 | 7.979 | Hippuric acid | Tier 1 |
| PFPeS | GLU | 5 | 2 | 5 | 0.015 | 7.979 | Hippuric acid | Tier 1 |
| PFNA | HDL-C | 5 | 2 | 5 | -0.021 | 7.733 | Elaidic acid | Tier 1 |
| GenX | MCH | 5 | 2 | 4 | 0.021 | 7.817 | Hippuric acid | Tier 1 |
| PFPeS | MCH | 5 | 2 | 4 | 0.011 | 7.817 | Hippuric acid | Tier 1 |
| DONA | ALP | 5 | 2 | 4 | 0.024 | 7.261 | Hippuric acid | Tier 1 |
| GenX | ALP | 5 | 2 | 4 | 0.029 | 7.261 | Hippuric acid | Tier 1 |
| PFPeS | ALP | 5 | 2 | 4 | 0.020 | 7.261 | Hippuric acid | Tier 1 |
| DONA | GGT | 5 | 1 | 5 | 0.023 | 7.983 | Hippuric acid | Tier 1 |
| PFNA | GGT | 5 | 1 | 5 | 0.023 | 7.983 | Hippuric acid | Tier 1 |
| PFNA | GLU | 5 | 1 | 5 | 0.020 | 7.979 | Hippuric acid | Tier 1 |
| DONA | MCH | 5 | 1 | 4 | 0.017 | 7.817 | Hippuric acid | Tier 1 |

| | | | | | | | | |
|---|---|---|---|---|---|---|---|---|
| PFNA | MCH | 5 | 1 | 4 | 0.018 | 7.817 | Hippuric acid | Tier 1 |
| PFNA | ALP | 5 | 1 | 4 | 0.025 | 7.261 | Hippuric acid | Tier 1 |

*Note: dataset1 denotes the plasma subset; dataset2 denotes the urine subset when both datasets are shown.*

Supplementary Table S25. Serial mediation summary by disease-priority PFAS and omics mediator block in dataset1.

| PFAS | Mediator Block | Screened Chains | Bootstrapped Chains | Serial FDR Supported | Serial Nominal Supported | Component Supported | Median Serial Indirect | Best serial bootstrap P | Best Chain |
|---|---|---|---|---|---|---|---|---|---|
| DONA | core_lipid | 1197 | 21 | 1.90E+01 | 0 | 221 | 0.020 | 6.64E-03 | PC(16:0_16:1) -> RDW-SD |
| DONA | shared_small_molecule | 35 | 7 | 7.00E+00 | 0 | 2 | 0.016 | 6.64E-03 | Hippuric acid -> RDW-SD |
| GenX | core_lipid | 1197 | 19 | 1.80E+01 | 1 | 384 | 0.019 | 6.64E-03 | LPC(P-18:0) -> RDW-SD |
| GenX | shared_small_molecule | 35 | 12 | 1.10E+01 | 1 | 0 | 0.016 | 6.64E-03 | Elaidic acid -> RDW-SD |
| PFNA | core_lipid | 1197 | 18 | 1.80E+01 | 0 | 470 | 0.020 | 6.64E-03 | LPC(P-18:0) -> RDW-SD |
| PFNA | shared_small_molecule | 35 | 11 | 1.00E+01 | 0 | 4 | 0.018 | 6.64E-03 | Hippuric acid -> RDW-SD |
| PFPeS | core_lipid | 1197 | 20 | 2.00E+01 | 0 | 425 | 0.019 | 6.64E-03 | LPC(P-18:0) -> RDW-SD |
| PFPeS | shared_small_molecule | 35 | 12 | 1.10E+01 | 0 | 3 | 0.017 | 6.64E-03 | Hippuric acid -> RDW-SD |

Supplementary Table S26. Serial mediation summary by clinical marker and omics mediator block in dataset1.

| Clinical Marker | Mediator Block | Screened Chains | Bootstrapped Chains | Serial FDR Supported | Supported Any | Median Serial Indirect | Best serial bootstrap P | Best Chain |
|---|---|---|---|---|---|---|---|---|
| ALP | core_lipid | 684 | 12 | 1.10E+01 | 148 | 0.014 | 6.64E-03 | PFNA -> LPC(P-18:0) |
| ALP | shared_small_molecule | 20 | 5 | 4.00E+00 | 5 | 0.018 | 1.33E-02 | PFPeS -> Hippuric acid |
| GGT | core_lipid | 684 | 12 | 1.20E+01 | 202 | 0.020 | 6.64E-03 | PFNA -> LPC(18:0) |
| GGT | shared_small_molecule | 20 | 5 | 5.00E+00 | 6 | 0.017 | 6.64E-03 | PFNA -> Hippuric acid |
| GLU | core_lipid | 684 | 12 | 1.20E+01 | 350 | 0.020 | 6.64E-03 | PFPeS -> PE(P-18:0_22:5) |
| GLU | shared_small_molecule | 20 | 5 | 5.00E+00 | 6 | 0.013 | 6.64E-03 | PFNA -> Hippuric acid |
| HDL-C | core_lipid | 684 | 12 | 1.20E+01 | 236 | 0.017 | 6.64E-03 | PFNA -> LPC(17:1) |
| HDL-C | shared_small_molecule | 20 | 5 | 5.00E+00 | 6 | 0.016 | 1.33E-02 | GenX -> Elaidic acid |
| MCH | core_lipid | 684 | 6 | 4.00E+00 | 14 | 0.001 | 1.33E-02 | PFPeS -> SM(t35:5) |
| MCH | shared_small_molecule | 20 | 6 | 4.00E+00 | 6 | 0.006 | 1.33E-02 | PFPeS -> Hippuric acid |
| RDW-SD | core_lipid | 684 | 12 | 1.20E+01 | 318 | 0.070 | 6.64E-03 | PFPeS -> LPC(P-18:0) |
| RDW-SD | shared_small_molecule | 20 | 9 | 9.00E+00 | 11 | 0.064 | 6.64E-03 | PFNA -> Hippuric acid |
| TG | core_lipid | 684 | 12 | 1.20E+01 | 308 | 0.027 | 6.64E-03 | PFPeS -> LPC(20:2) |
| TG | shared_small_molecule | 20 | 7 | 7.00E+00 | 9 | 0.024 | 6.64E-03 | PFNA -> Hippuric acid |

Supplementary Table S27. Network nodes for integrated PFAS-omics-clinical-ovarian cancer figures in dataset1.

| Network | Node Id | Label | Layer | Node Type | X | Y |
|---|---|---|---|---|---|---|
| aggregated_class | PFAS:DONA | DONA | PFAS | PFAS | 1 | 4 |
| aggregated_class | PFAS:GenX | GenX | PFAS | PFAS | 1 | 3 |
| aggregated_class | PFAS:PFNA | PFNA | PFAS | PFAS | 1 | 2 |
| aggregated_class | PFAS:PFPeS | PFPeS | PFAS | PFAS | 1 | 1 |
| aggregated_class | M1:LPC | LPC | Omics mediator class | Core lipid class | 2 | 8 |
| aggregated_class | M1:PC | PC | Omics mediator class | Core lipid class | 2 | 7 |
| aggregated_class | M1:PE | PE | Omics mediator class | Core lipid class | 2 | 6 |
| aggregated_class | M1:SM | SM | Omics mediator class | Core lipid class | 2 | 5 |
| aggregated_class | M1:TG | TG | Omics mediator class | Core lipid class | 2 | 4 |
| aggregated_class | M1:DG | DG | Omics mediator class | Core lipid class | 2 | 3 |
| aggregated_class | M1:AcCa | AcCa | Omics mediator class | Core lipid class | 2 | 2 |
| aggregated_class | M1:Shared SM | Shared SM | Omics mediator class | Supportive metabolomics | 2 | 1 |
| aggregated_class | Clinical:TG | TG | Clinical phenotypes | Clinical phenotype | 3 | 7 |
| aggregated_class | Clinical:HDL-C | HDL-C | Clinical phenotypes | Clinical phenotype | 3 | 6 |
| aggregated_class | Clinical:GGT | GGT | Clinical phenotypes | Clinical phenotype | 3 | 5 |
| aggregated_class | Clinical:ALP | ALP | Clinical phenotypes | Clinical phenotype | 3 | 4 |
| aggregated_class | Clinical:GLU | GLU | Clinical phenotypes | Clinical phenotype | 3 | 3 |
| aggregated_class | Clinical:RDW-SD | RDW-SD | Clinical phenotypes | Clinical phenotype | 3 | 2 |
| aggregated_class | Clinical:MCH | MCH | Clinical phenotypes | Clinical phenotype | 3 | 1 |
| aggregated_class | Disease:Disease risk | Disease risk | Outcome | Disease | 4 | 4 |
| bootstrap_fdr_supported | PFAS:DONA | DONA | PFAS | PFAS | 1 | 4 |
| bootstrap_fdr_supported | PFAS:GenX | GenX | PFAS | PFAS | 1 | 3 |
| bootstrap_fdr_supported | PFAS:PFNA | PFNA | PFAS | PFAS | 1 | 2 |
| bootstrap_fdr_supported | PFAS:PFPeS | PFPeS | PFAS | PFAS | 1 | 1 |

| | | | | | | |
|---|---|---|---|---|---|---|
| bootstrap_fdr_supported | M1:LPC | LPC | Omics mediators | Core lipid class | 2 | 10 |
| bootstrap_fdr_supported | M1:PC | PC | Omics mediators | Core lipid class | 2 | 9 |
| bootstrap_fdr_supported | M1:PE | PE | Omics mediators | Core lipid class | 2 | 8 |
| bootstrap_fdr_supported | M1:SM | SM | Omics mediators | Core lipid class | 2 | 7 |
| bootstrap_fdr_supported | M1:TG | TG | Omics mediators | Core lipid class | 2 | 6 |
| bootstrap_fdr_supported | M1:AcCa | AcCa | Omics mediators | Core lipid class | 2 | 5 |
| bootstrap_fdr_supported | M1:Hippuric acid | Hippuric acid | Omics mediators | Supportive metabolomics | 2 | 4 |
| bootstrap_fdr_supported | M1:Elaidic acid | Elaidic acid | Omics mediators | Supportive metabolomics | 2 | 3 |
| bootstrap_fdr_supported | M1:Corticosterone | Corticosterone | Omics mediators | Supportive metabolomics | 2 | 2 |
| bootstrap_fdr_supported | M1:Betaine | Betaine | Omics mediators | Supportive metabolomics | 2 | 1 |
| bootstrap_fdr_supported | Clinical:TG | TG | Clinical phenotypes | Clinical phenotype | 3 | 7 |
| bootstrap_fdr_supported | Clinical:HDL-C | HDL-C | Clinical phenotypes | Clinical phenotype | 3 | 6 |
| bootstrap_fdr_supported | Clinical:GGT | GGT | Clinical phenotypes | Clinical phenotype | 3 | 5 |
| bootstrap_fdr_supported | Clinical:ALP | ALP | Clinical phenotypes | Clinical phenotype | 3 | 4 |
| bootstrap_fdr_supported | Clinical:GLU | GLU | Clinical phenotypes | Clinical phenotype | 3 | 3 |
| bootstrap_fdr_supported | Clinical:RDW-SD | RDW-SD | Clinical phenotypes | Clinical phenotype | 3 | 2 |
| bootstrap_fdr_supported | Clinical:MCH | MCH | Clinical phenotypes | Clinical phenotype | 3 | 1 |
| bootstrap_fdr_supported | Disease:Disease risk | Disease risk | Outcome | Disease | 4 | 4 |

Supplementary Table S28. Feature sets used for shared-metabolite biomarker validation models using logistic regression.

| Dataset | Model Id | N Features | Feature Names | Interpretation |
|---|---|---|---|---|
| dataset1 | A_Basic_clinical | 3 | age; bmi; hypertension | Comparator model. |
| dataset1 | B_Routine_clinical | 10 | age; bmi; hypertension; TG; HDL-C; GLU; GGT; ALP; RDW-SD; MCH | Comparator model. |
| dataset1 | C_Cancer_markers_positive_reference | 4 | CA125; CEA; CA15-3; AFP | Positive reference model using conventional cancer markers. |
| dataset1 | D_Core5_shared_metabolites | 5 | Elaidic acid; Hippuric acid; Betaine; Corticosterone; Taurocholic acid | Shared plasma/urine metabolite signature alone. |
| dataset1 | E_Routine_plus_Core5 | 15 | age; bmi; hypertension; TG; HDL-C; GLU; GGT; ALP; RDW-SD; MCH; Elaidic acid; Hippuric acid; Betaine; Corticosterone; Taurocholic acid | Comparator model. |
| dataset1 | F_Cancer_markers_plus_Core5 | 9 | CA125; CEA; CA15-3; AFP; Elaidic acid; Hippuric acid; Betaine; Corticosterone; Taurocholic acid | Tests whether Core-5 adds to the cancer-marker positive reference. |
| dataset1 | G_Routine_plus_Cancer_reference | 14 | age; bmi; hypertension; TG; HDL-C; GLU; GGT; ALP; RDW-SD; MCH; CA125; CEA; CA15-3; AFP | Comparator model. |
| dataset1 | H_Routine_plus_Cancer_plus_Core5 | 19 | age; bmi; hypertension; TG; HDL-C; GLU; GGT; ALP; RDW-SD; MCH; CA125; CEA; CA15-3; AFP; Elaidic acid; Hippuric acid; Betaine; Corticosterone; Taurocholic acid | Strictest expanded model testing Core-5 beyond routine clinical plus cancer markers. |
| dataset2 | A_Basic_clinical | 3 | age; bmi; hypertension | Comparator model. |
| dataset2 | B_Routine_clinical | 10 | age; bmi; hypertension; TG; HDL-C; GLU; GGT; ALP; RDW-SD; MCH | Comparator model. |

| | | | | |
|---|---|---|---|---|
| dataset2 | C_Cancer_markers_positive_reference | 4 | CA125; CEA; CA15-3; AFP | Positive reference model using conventional cancer markers. |
| dataset2 | D_Core5_shared_metabolites | 5 | Elaidic acid; Hippuric acid; Betaine; Corticosterone; Taurocholic acid | Shared plasma/urine metabolite signature alone. |
| dataset2 | E_Routine_plus_Core5 | 15 | age; bmi; hypertension; TG; HDL-C; GLU; GGT; ALP; RDW-SD; MCH; Elaidic acid; Hippuric acid; Betaine; Corticosterone; Taurocholic acid | Comparator model. |
| dataset2 | F_Cancer_markers_plus_Core5 | 9 | CA125; CEA; CA15-3; AFP; Elaidic acid; Hippuric acid; Betaine; Corticosterone; Taurocholic acid | Tests whether Core-5 adds to the cancer-marker positive reference. |
| dataset2 | G_Routine_plus_Cancer_reference | 14 | age; bmi; hypertension; TG; HDL-C; GLU; GGT; ALP; RDW-SD; MCH; CA125; CEA; CA15-3; AFP | Comparator model. |
| dataset2 | H_Routine_plus_Cancer_plus_Core5 | 19 | age; bmi; hypertension; TG; HDL-C; GLU; GGT; ALP; RDW-SD; MCH; CA125; CEA; CA15-3; AFP; Elaidic acid; Hippuric acid; Betaine; Corticosterone; Taurocholic acid | Strictest expanded model testing Core-5 beyond routine clinical plus cancer markers. |

*Note: dataset1 denotes the plasma subset; dataset2 denotes the urine subset when both datasets are shown.*

Supplementary Table S29. Incremental performance comparisons for Core-5 shared-metabolite feature sets using logistic regression.

| Dataset | Algorithm | Comparison Id | Base Model | Expanded Model | N Fold Pairs | Delta AUC mean | Delta AUC median | Delta AUC 2.5th pct | Delta AUC 97.5th pct | % folds AUC improved | Delta Brier mean | % folds Brier improved | Conclusion Flag |
|---|---|---|---|---|---|---|---|---|---|---|---|---|---|
| dataset1 | glm_fixed | D_Core5_vs_C_Cancer_positive_reference | C_Cancer_markers_positive_reference | D_Core5_shared_metabolites | 100 | -0.035 | -0.041 | -0.132 | 0.065 | 29% | 0.022 | 32% | no_internal_incremental_value |
| dataset1 | glm_fixed | E_Routine_plus_Core5_vs_B_Routine | B_Routine_clinical | E_Routine_plus_Core5 | 100 | 0.021 | 0.029 | -0.083 | 0.080 | 75% | -0.028 | 85% | probable_internal_incremental_value |
| dataset1 | glm_fixed | F_Cancer_plus_Core5_vs_C_Cancer | C_Cancer_markers_positive_reference | F_Cancer_markers_plus_Core5 | 100 | 0.045 | 0.042 | -0.002 | 0.108 | 93% | -0.039 | 95% | probable_internal_incremental_value |
| dataset1 | glm_fixed | H_Full_plus_Core5_vs_G_Routine_plus_Cancer | G_Routine_plus_Cancer_reference | H_Routine_plus_Cancer_plus_Core5 | 100 | -0.027 | -0.025 | -0.096 | 0.032 | 18% | 0.011 | 37% | no_internal_incremental_value |
| dataset1 | glmnet_elastic_1se | D_Core5_vs_C_Cancer_positive_reference | C_Cancer_markers_positive_reference | D_Core5_shared_metabolites | 100 | -0.036 | -0.037 | -0.165 | 0.095 | 30% | -0.006 | 53% | no_internal_incremental_value |

| | | | | | | | | | | | | | |
|---|---|---|---|---|---|---|---|---|---|---|---|---|---|
| dataset1 | glmnet_elastic_1se | E_Routine_plus_Core5_vs_B_Routine | B_Routine_clinical | E_Routine_plus_Core5 | 100 | 0.049 | 0.048 | 0.008 | 0.094 | 100% | -0.064 | 89% | consistent_internal_incremental_value |
| dataset1 | glmnet_elastic_1se | F_Cancer_plus_Core5_vs_C_Cancer | C_Cancer_markers_positive_reference | F_Cancer_markers_plus_Core5 | 100 | 0.053 | 0.054 | -0.002 | 0.121 | 96% | -0.064 | 100% | probable_internal_incremental_value |
| dataset1 | glmnet_elastic_1se | H_Full_plus_Core5_vs_G_Routine_plus_Cancer | G_Routine_plus_Cancer_reference | H_Routine_plus_Cancer_plus_Core5 | 100 | 0.017 | 0.018 | -0.007 | 0.047 | 86% | -0.044 | 85% | probable_internal_incremental_value |
| dataset1 | glmnet_lasso_1se | D_Core5_vs_C_Cancer_positive_reference | C_Cancer_markers_positive_reference | D_Core5_shared_metabolites | 100 | -0.044 | -0.042 | -0.182 | 0.078 | 27% | 0.019 | 25% | no_internal_incremental_value |
| dataset1 | glmnet_lasso_1se | E_Routine_plus_Core5_vs_B_Routine | B_Routine_clinical | E_Routine_plus_Core5 | 100 | 0.048 | 0.048 | 0.001 | 0.099 | 97% | -0.046 | 94% | consistent_internal_incremental_value |
| dataset1 | glmnet_lasso_1se | F_Cancer_plus_Core5_vs_C_Cancer | C_Cancer_markers_positive_reference | F_Cancer_markers_plus_Core5 | 100 | 0.049 | 0.044 | 0.003 | 0.113 | 98% | -0.049 | 97% | consistent_internal_incremental_value |
| dataset1 | glmnet_lasso_1se | H_Full_plus_Core5_vs_G_Routine_plus_Cancer | G_Routine_plus_Cancer_reference | H_Routine_plus_Cancer_plus_Core5 | 100 | 0.015 | 0.015 | -0.009 | 0.046 | 80% | -0.028 | 82% | probable_internal_incremental_value |
| dataset2 | glm_fixed | D_Core5_vs_C_Cancer_positive_reference | C_Cancer_markers_positive_reference | D_Core5_shared_metabolites | 100 | 0.010 | 0.010 | -0.115 | 0.144 | 55% | 0.000 | 50% | small_or_uncertain_internal_i |

| | | | | | | | | | | | | | |
|---|---|---|---|---|---|---|---|---|---|---|---|---|---|
| | | | | | | | | | | | | | ncremental_value |
| dataset2 | glm_fixed | E_Routine_plus_Core5_vs_B_Routine | B_Routine_clinical | E_Routine_plus_Core5 | 100 | 0.074 | 0.071 | -0.014 | 0.159 | 96% | -0.049 | 89% | probable_internal_incremental_value |
| dataset2 | glm_fixed | F_Cancer_plus_Core5_vs_C_Cancer | C_Cancer_markers_positive_reference | F_Cancer_markers_plus_Core5 | 100 | 0.057 | 0.056 | -0.004 | 0.135 | 96% | -0.039 | 92% | probable_internal_incremental_value |
| dataset2 | glm_fixed | H_Full_plus_Core5_vs_G_Routine_plus_Cancer | G_Routine_plus_Cancer_reference | H_Routine_plus_Cancer_plus_Core5 | 100 | 0.009 | 0.015 | -0.057 | 0.049 | 65% | 0.006 | 51% | small_or_uncertain_internal_incremental_value |
| dataset2 | glmnet_elastic_1se | D_Core5_vs_C_Cancer_positive_reference | C_Cancer_markers_positive_reference | D_Core5_shared_metabolites | 100 | 0.007 | 0.006 | -0.134 | 0.132 | 52% | -0.056 | 97% | small_or_uncertain_internal_incremental_value |
| dataset2 | glmnet_elastic_1se | E_Routine_plus_Core5_vs_B_Routine | B_Routine_clinical | E_Routine_plus_Core5 | 100 | 0.081 | 0.085 | 0.006 | 0.171 | 98% | -0.048 | 99% | consistent_internal_incremental_value |
| dataset2 | glmnet_elastic_1se | F_Cancer_plus_Core5_vs_C_Cancer | C_Cancer_markers_positive_reference | F_Cancer_markers_plus_Core5 | 100 | 0.059 | 0.053 | 0.001 | 0.123 | 97% | -0.083 | 100% | consistent_internal_incremental_value |
| dataset2 | glmnet_elastic_1se | H_Full_plus_Core5_vs_G_Routine_plus_Cancer | G_Routine_plus_Cancer_reference | H_Routine_plus_Cancer_plus_Core5 | 100 | 0.027 | 0.026 | -0.010 | 0.086 | 88% | -0.028 | 79% | probable_internal_incremental_value |

| | | | | | | | | | | | | | |
|---|---|---|---|---|---|---|---|---|---|---|---|---|---|
| dataset2 | glmnet_lasso_1se | D_Core5_vs_C_Cancer_positive_reference | C_Cancer_markers_positive_reference | D_Core5_shared_metabolites | 100 | 0.008 | 0.005 | -0.119 | 0.144 | 52% | -0.039 | 85% | small_or_uncertain_internal_incremental_value |
| dataset2 | glmnet_lasso_1se | E_Routine_plus_Core5_vs_B_Routine | B_Routine_clinical | E_Routine_plus_Core5 | 100 | 0.089 | 0.086 | -0.008 | 0.184 | 95% | -0.047 | 96% | probable_internal_incremental_value |
| dataset2 | glmnet_lasso_1se | F_Cancer_plus_Core5_vs_C_Cancer | C_Cancer_markers_positive_reference | F_Cancer_markers_plus_Core5 | 100 | 0.066 | 0.061 | -0.007 | 0.147 | 96% | -0.072 | 99% | probable_internal_incremental_value |
| dataset2 | glmnet_lasso_1se | H_Full_plus_Core5_vs_G_Routine_plus_Cancer | G_Routine_plus_Cancer_reference | H_Routine_plus_Cancer_plus_Core5 | 100 | 0.026 | 0.024 | -0.023 | 0.065 | 84% | -0.019 | 73% | probable_internal_incremental_value |

*Note: Positive delta AUC favors the expanded model; negative delta Brier favors the expanded model.*

Supplementary Table S30. Feature sets used for shared-metabolite biomarker validation models using XGBoost.

| Dataset | Model Id | N Features | Feature Names |
|---|---|---|---|
| dataset1 | A_Basic_clinical | 3 | age; bmi; hypertension |
| dataset1 | B_Routine_clinical | 10 | age; bmi; hypertension; TG; HDL-C; GLU; GGT; ALP; RDW-SD; MCH |
| dataset1 | C_Cancer_markers_positive_reference | 4 | CA125; CEA; CA15-3; AFP |
| dataset1 | D_Core5_shared_metabolites | 5 | Elaidic acid; Hippuric acid; Betaine; Corticosterone; Taurocholic acid |
| dataset1 | E_Routine_plus_Core5 | 15 | age; bmi; hypertension; TG; HDL-C; GLU; GGT; ALP; RDW-SD; MCH; Elaidic acid; Hippuric acid; Betaine; Corticosterone; Taurocholic acid |
| dataset1 | F_Cancer_markers_plus_Core5 | 9 | CA125; CEA; CA15-3; AFP; Elaidic acid; Hippuric acid; Betaine; Corticosterone; Taurocholic acid |
| dataset1 | G_Routine_plus_Cancer_reference | 14 | age; bmi; hypertension; TG; HDL-C; GLU; GGT; ALP; RDW-SD; MCH; CA125; CEA; CA15-3; AFP |
| dataset1 | H_Routine_plus_Cancer_plus_Core5 | 19 | age; bmi; hypertension; TG; HDL-C; GLU; GGT; ALP; RDW-SD; MCH; CA125; CEA; CA15-3; AFP; Elaidic acid; Hippuric acid; Betaine; Corticosterone; Taurocholic acid |
| dataset2 | A_Basic_clinical | 3 | age; bmi; hypertension |
| dataset2 | B_Routine_clinical | 10 | age; bmi; hypertension; TG; HDL-C; GLU; GGT; ALP; RDW-SD; MCH |
| dataset2 | C_Cancer_markers_positive_reference | 4 | CA125; CEA; CA15-3; AFP |
| dataset2 | D_Core5_shared_metabolites | 5 | Elaidic acid; Hippuric acid; Betaine; Corticosterone; Taurocholic acid |
| dataset2 | E_Routine_plus_Core5 | 15 | age; bmi; hypertension; TG; HDL-C; GLU; GGT; ALP; RDW-SD; MCH; Elaidic acid; Hippuric acid; Betaine; Corticosterone; Taurocholic acid |
| dataset2 | F_Cancer_markers_plus_Core5 | 9 | CA125; CEA; CA15-3; AFP; Elaidic acid; Hippuric acid; Betaine; Corticosterone; Taurocholic acid |
| dataset2 | G_Routine_plus_Cancer_reference | 14 | age; bmi; hypertension; TG; HDL-C; GLU; GGT; ALP; RDW-SD; MCH; CA125; CEA; CA15-3; AFP |

| | | | |
|---|---|---|---|
| dataset2 | H_Routine_plus_Cancer_plus_Core5 | 19 | age; bmi; hypertension; TG; HDL-C; GLU; GGT; ALP; RDW-SD; MCH; CA125; CEA; CA15-3; AFP; Elaidic acid; Hippuric acid; Betaine; Corticosterone; Taurocholic acid |

*Note: dataset1 denotes the plasma subset; dataset2 denotes the urine subset when both datasets are shown.*

Supplementary Table S31. Incremental performance comparisons for Core-5 shared-metabolite feature sets using XGBoost.

| Dataset | Algorithm | Comparison Id | Base Model | Expanded Model | N Fold Pairs | Delta AUC mean | Delta AUC median | Delta AUC 2.5th pct | Delta AUC 97.5th pct | % folds AUC improved | Delta Brier mean | % folds Brier improved | Conclusion Flag |
|---|---|---|---|---|---|---|---|---|---|---|---|---|---|
| dataset1 | xgboost_fixed_earlystop | D_Core5_vs_C_Cancer_positive_reference | C_Cancer_markers_positive_reference | D_Core5_shared_metabolites | 50 | 0.030 | 0.028 | -0.064 | 0.143 | 66% | -0.024 | 58% | small_or_uncertain_xgboost_incremental_value |
| dataset1 | xgboost_fixed_earlystop | E_Routine_plus_Core5_vs_B_Routine | B_Routine_clinical | E_Routine_plus_Core5 | 50 | 0.061 | 0.070 | -0.065 | 0.135 | 90% | -0.023 | 56% | probable_xgboost_incremental_value |
| dataset1 | xgboost_fixed_earlystop | F_Cancer_plus_Core5_vs_C_Cancer | C_Cancer_markers_positive_reference | F_Cancer_markers_plus_Core5 | 50 | 0.053 | 0.055 | -0.075 | 0.140 | 88% | -0.023 | 62% | probable_xgboost_incremental_value |
| dataset1 | xgboost_fixed_earlystop | H_Full_plus_Core5_vs_G_Routine_plus_Cancer | G_Routine_plus_Cancer_reference | H_Routine_plus_Cancer_plus_Core5 | 50 | 0.037 | 0.040 | -0.030 | 0.116 | 88% | -0.021 | 66% | probable_xgboost_incremental_value |
| dataset2 | xgboost_fixed_earlystop | D_Core5_vs_C_Cancer_positive_reference | C_Cancer_markers_positive_reference | D_Core5_shared_metabolites | 50 | -0.045 | -0.034 | -0.206 | 0.062 | 32% | 0.018 | 38% | no_xgboost_incremental_value |

| | | | | | | | | | | | | |
|---|---|---|---|---|---|---|---|---|---|---|---|---|
| dataset2 | xgboost_fixed_earlystop | E_Routine_plus_Core5_vs_B_Routine | B_Routine_clinical | E_Routine_plus_Core5 | 50 | 0.046 | 0.056 | -0.110 | 0.171 | 78% | -0.024 | 72% | probable_xgboost_incremental_value |
| dataset2 | xgboost_fixed_earlystop | F_Cancer_plus_Core5_vs_C_Cancer | C_Cancer_markers_positive_reference | F_Cancer_markers_plus_Core5 | 50 | 0.018 | 0.026 | -0.084 | 0.094 | 78% | -0.008 | 58% | probable_xgboost_incremental_value |
| dataset2 | xgboost_fixed_earlystop | H_Full_plus_Core5_vs_G_Routine_plus_Cancer | G_Routine_plus_Cancer_reference | H_Routine_plus_Cancer_plus_Core5 | 50 | 0.025 | 0.018 | -0.029 | 0.123 | 78% | -0.010 | 64% | probable_xgboost_incremental_value |

*Note: Positive delta AUC favors the expanded model; negative delta Brier favors the expanded model.*

Supplementary Table S32. Optimized multiple reaction monitoring (MRM) parameters for targeted PFAS analysis.

| Acronym | Formula | Parent ions | Ion 1 [a] | CE(eV) | Ion 2 [b] | CE(eV) | DP | RT(min) |
|---|---|---|---|---|---|---|---|---|
| PFBA | C4HF7O2 | 213 | 168.9 | -15 | 89 | -15 | -60 | 3.88 |
| 13C4-PFBA | [13]C4HF7O2 | 216.9 | 172 | -13 | \ | \ | -50 | 3.85 |
| PFPeA | C5HF9O2 | 262.9 | 218.9 | -13 | 168.9 | -13 | -60 | 5.75 |
| 13C5-PFPeA | [13]C5HF9O2 | 267.7 | 222.8 | -14 | \ | \ | -100 | 5.74 |
| FBSA | C4H2F9NO2S | 298 | 77.9 | -27 | 119 | -31 | -60 | 6.62 |
| PFBS | C4HF9O3S | 298.9 | 79.9 | -25 | 98.9 | -55 | -80 | 5.95 |
| 13C3-PFBS | [13]C3CHF9O3S | 301.5 | 82.8 | -67.5 | \ | \ | -100 | 8.05 |
| PFHxA | C6HF11O2 | 312.9 | 268.9 | -36 | 118.9 | -61 | -100 | 6.49 |
| 13C5-PFHxA | [13]C5CHF11O2 | 317.7 | 272.8 | -17 | \ | \ | -100 | 5.94 |
| HFPO-DA | C6HF11O3 | 329 | 255.2 | -17 | 168.9 | -40 | -50 | 6.62 |
| PFPeS | C5HF11O3S | 348.6 | 79.9 | -14 | 98.8 | -14 | -60 | 6.55 |
| PFHpA | C7HF13O2 | 362.9 | 318.9 | -75 | 168.9 | -24 | -100 | 6.94 |
| 13C4-PFHpA | [13]C4C3HF13O2 | 366.5 | 321.8 | -13.6 | \ | \ | -50 | 6.47 |
| DONA | C7H2F12O4 | 376.7 | 84.9 | -14 | 250.8 | -40 | -50 | 6.96 |
| PFHxS | C6HF13O3S | 398.9 | 79.9 | -20 | 98.9 | -42 | -60 | 6.97 |
| 13C3- PFHxS | [13]C3C3HF13O3S | 401.6 | 79.8 | -90 | \ | \ | -80 | 8.25 |
| PFOA | C8HF15O2 | 412.9 | 368.9 | -85 | 168.9 | -24 | -100 | 7.29 |
| 13C8-PFOA | [13]C8HF15O2 | 420.6 | 375.7 | -15 | \ | \ | -80 | 6.91 |
| 6:2FTSA | C8H5F13O3S | 427.1 | 409 | -33.6 | 389 | -95 | -120 | 7.29 |
| PFHpS | C7HF15O3S | 448.9 | 79.9 | -15 | 98.8 | -70 | -50 | 7.29 |
| PFNA | C9HF17O2 | 462.9 | 418.9 | -100 | 168.9 | -27 | -100 | 7.58 |
| 13C9-PFNA | [13]C9HF17O2 | 471.7 | 426.7 | -15 | \ | \ | -80 | 6.94 |
| PFOS | C8HF17O3S | 498.9 | 79.9 | -16 | 98.9 | -92 | -50 | 7.58 |

| | | | | | | | | |
|---|---|---|---|---|---|---|---|---|
| 13C8-PFOS | [13]C8HF17O3S | 506.6 | 79.8 | -90 | \ | \ | -80 | 8.6 |
| PFDA | C10HF19O2 | 512.9 | 468.9 | -37 | 268.9 | -100 | -100 | 7.84 |
| 13C6-PFDA | [13]C6C4HF19O2 | 518.6 | 473.7 | -17.8 | \ | \ | -80 | 7.28 |
| 6:2Cl-PFESA | C8ClF16HO4S | 530.5 | 351 | -110 | 199 | -27 | -100 | 7.71 |
| PFNS | C9H3F19O3S | 548.5 | 79.8 | -17 | 98.9 | -67 | -60 | 7.81 |
| PFUdA | C11HF21O2 | 562.9 | 518.9 | -120 | 268.9 | -27 | -100 | 8.06 |
| 13C7-PFUdA | [13]C7C4HF21O2 | 569.6 | 524.7 | -16.8 | \ | \ | -80 | 7.55 |
| PFDS | C10HF21O3S | 598.9 | 79.9 | -18 | 98.9 | -110 | -60 | 8.06 |
| PFDoA | C12HF23O2 | 612.9 | 568.9 | -18 | 268.9 | -90 | -60 | 8.27 |
| 13C2-PFDoA | [13]C2C10HF23O2 | 614.9 | 569.9 | -19 | \ | \ | -80 | 7.57 |
| 8:2Cl-PFESA | C10ClF20HO4S | 630.9 | 82.9 | -125 | 451.1 | -27 | -50 | 8.12 |
| PFTrDA | C13HF25O2 | 662.9 | 618.9 | -40 | 268.9 | -33 | -100 | 8.44 |
| PFTeDA | C14HF27O2 | 712.9 | 668.9 | -20 | 268.9 | -35 | -60 | 8.61 |
| 13C2-PFTeDA | [13]C2C12HF27O2 | 714.5 | 669.6 | -20.7 | \ | \ | -80 | 7.83 |
| PFHxDA | C16HF31O2 | 812.9 | 768.9 | -20 | 268.9 | -42 | -60 | 8.93 |
| PFOdA | C18HF35O2 | 912.9 | 868.9 | -21 | 568.9 | -42 | -60 | 9.21 |

a: quantitative ion.

b: qualitative ion.

Supplementary Table S33. Calibration equations, coefficients of determination, limits of detection and quantification, recovery, and precision for targeted PFAS analysis.

| Compound | Regression equation | R2 | LOD (μg/L) | LOQ (μg/L) | Recovery % (1 μg/L) | Recovery % (10 μg/L) | RSD |
|---|---|---|---|---|---|---|---|
| 6:2CI-PFESA | y = 0.09701 x + 0.00241 | 0.99346 | 0.002 | 0.006 | 78.4 | 91.48 | 8.1 |
| 6:2FTSA | y = 0.19761 x + 1.4577 | 0.999 | 0.008 | 0.03 | 120.2 | 116.7 | 4.2 |
| 8:2CI-PFESA | y = 0.88191 x + 0.05250 | 0.99806 | 0.004 | 0.012 | 129.4 | 106.16 | 5.6 |
| DONA | y = 0.57052 x + 0.04188 | 0.99987 | 0.006 | 0.024 | 71.6 | 101.3 | 1.8 |
| FBSA | y = 0.85846 x + 0.01573 | 0.9912 | 0.002 | 0.008 | 77.1 | 109 | 7.7 |
| HFPO-DA(GenX) | y = 0.10346 x + 0.00126 | 0.99427 | 0.004 | 0.016 | 119.2 | 83 | 12.3 |
| PFBA | y = 0.16026 x + 0.00389 | 0.99868 | 0.008 | 0.03 | 99.7 | 104.7 | 4.1 |
| PFBS | y = 0.25057 x + 0.17016 | 0.99713 | 0.002 | 0.008 | 143.6 | 104.12 | 10.5 |
| PFDA | y = 0.43973 x + 0.01063 | 0.99941 | 0.008 | 0.032 | 109.8 | 86.34 | 7.9 |
| PFDoA | y = 0.19189 x + 0.2812 | 0.99547 | 0.006 | 0.02 | 129.4 | 87.96 | 5.7 |
| PFDS | y = 0.61913 x + 0.02203 | 0.99463 | 0.004 | 0.016 | 116.8 | 108.3 | 6.1 |
| PFHpA | y = 1.40801 x + 0.07199 | 0.99911 | 0.022 | 0.074 | 117.6 | 63.08 | 8.4 |
| PFHpS | y = 0.31540 x + 0.07223 | 0.99886 | 0.004 | 0.012 | 115.4 | 77.38 | 6.3 |
| PFHxA | y = 0.25057 x + 0.17016 | 0.99713 | 0.016 | 0.056 | 134.2 | 72.68 | 14.2 |
| PFHxDA | y = 0.35742 x + 0.00188 | 0.9991 | 0.016 | 0.054 | 97.6 | 115.54 | 9.13 |
| PFHxS | y = 1.96947 x + 0.14404 | 0.99235 | 0.006 | 0.02 | 91.6 | 98.46 | 6.2 |
| PFNA | y = 1.64896 x + 0.23217 | 0.99345 | 0.008 | 0.03 | 104.8 | 79 | 7.6 |
| PFNS | y = 0.02254 x + 0.03046 | 0.99246 | 0.002 | 0.008 | 64.2 | 72.34 | 8.4 |
| PFOA | y = 0.54963 x + 0.04730 | 0.99812 | 0.004 | 0.012 | 91.2 | 91.9 | 7.5 |
| PFOdA | y = 0.22154 x + 0.23046 | 0.99846 | 0.002 | 0.006 | 156.8 | 99.22 | 13.8 |
| PFOS | y = 0.34476 x + 0.31829 | 0.99765 | 0.006 | 0.022 | 83.8 | 92.96 | 9.3 |
| PFPeA | y = 0.75892 x + 0.09364 | 0.99783 | 0.034 | 0.112 | 84.6 | 121.6 | 2.2 |
| PFPeS | y = 0.91946 x + 0.10197 | 0.999 | 0.004 | 0.014 | 97.6 | 80.2 | 12.7 |
| PFTeDA | y = 0.39603 x + 0.00922 | 0.99198 | 0.01 | 0.036 | 112.6 | 97.94 | 13.8 |

| | | | | | | | |
|---|---|---|---|---|---|---|---|
| PFTrDA | y = 1.06730 x + 0.09720 | 0.99743 | 0.012 | 0.042 | 81 | 129.86 | 8.1 |
| PFUdA | y = 0.02254 x + 0.23046 | 0.99846 | 0.006 | 0.02 | 103.4 | 94.28 | 11.9 |

### Supplementary Methods

#### S1.Targeted PFAS quantification and quality control

Detailed procedures for targeted PFAS analysis were based on an isotope-dilution UPLC-MS/MS workflow. Each sample was processed using 200 μL of plasma. Isotope-labeled internal standards were added before sample preparation, followed by protein precipitation or solid-phase extraction, centrifugation, concentration, and reconstitution before instrumental analysis. The analytical sequence was designed to minimize PFAS background contamination by using PFAS-free materials during collection, processing, and storage. **Detailed information on reagent selection and sample-preparation procedures has been reported in our previous study and is therefore not repeated here (*1*).**

The UPLC-MS/MS system was operated in multiple reaction monitoring mode. Quantification was based on matrix-matched calibration curves or standard curves, with signal correction by isotope-labeled internal standards. Concentrations below the limit of detection were replaced by the limit of detection divided by the square root of 2. For regression analyses, PFAS concentrations were log2-transformed and then standardized to z scores; therefore, odds ratios (ORs) for single-PFAS models correspond to a one-standard-deviation increase in log2-transformed PFAS concentration.

#### S2. Plasma untargeted small-molecule metabolomics

Plasma metabolite extraction was performed by organic-solvent protein precipitation and small-molecule extraction. Briefly, 100 μL of human plasma was mixed with a fourfold volume of precooled methanol/acetonitrile (1:1, v/v), vortexed for 30 s, sonicated in an ice-water bath for 10 min, and incubated at -20 °C for 1 h. Samples were centrifuged at 12,000 rpm for 15 min at 4 °C, and the supernatant was collected and vacuum-dried. Dried extracts were reconstituted in 100 μL acetonitrile/water (1:1, v/v), vortexed for 30 s, sonicated in an ice-water bath for 5 min, and centrifuged again at 12,000 rpm for 15 min at 4 °C. The final supernatant was transferred to LC-MS vials. Pooled QC samples were prepared by mixing equal-volume aliquots from study samples and were extracted and analyzed in parallel with the study samples. Cases and controls were randomized across sample-preparation and injection batches to reduce batch and run-order effects.

Chromatographic separation was performed using an ACQUITY UPLC BEH C18 column with a matched guard column at 40 °C. The run time was 25 min, the flow rate was 0.30 mL/min, and the injection volume was 5 μL. Positive-mode mobile phase A was water containing 0.1% formic acid, and mobile phase B was acetonitrile. Negative-mode mobile phase A was 0.1% $NH_3$-20 mM ammonium acetate in water, and mobile phase B was acetonitrile. Raw data were acquired using Xcalibur v4.1.

Raw files were processed in Compound Discoverer v3.3 for feature detection, peak alignment, gap filling, QC-based normalization, and metabolite annotation. The mass tolerance for precursor and fragment ions was set at 10 ppm, and the signal-to-noise threshold was set at 1.5. Features with RSD >=30% in pooled QC samples were excluded before downstream analysis, and only high-confidence annotated metabolites were retained. Detailed liquid chromatography–mass spectrometry acquisition parameters and chromatographic gradient conditions have been described in our previous study and are not repeated here (*2*).

**S3. Plasma lipidomics**

Plasma lipid extraction followed a modified Matyash biphasic extraction protocol (*3*). Briefly, 200 μL of plasma was thawed at 4 °C and mixed with 600 μL precooled methanol and 2 mL methyl tert-butyl ether (MTBE), corresponding to a methanol:MTBE ratio of 1.5:5 (v/v). The mixture was vortexed, sonicated in an ice-water bath for 10 min, incubated at -20 °C for 30 min, and centrifuged at 13,000 rpm for 15 min at 4 °C. The supernatant was collected and dried under gentle nitrogen at low temperature. The residue was reconstituted in 200 μL acetonitrile/isopropanol (7:3, v/v), sonicated for 3 min, centrifuged under the same conditions, filtered through a 0.22 μm organic-phase membrane, transferred to LC-MS vials with glass inserts, and stored at -80 °C until analysis.

The lipidomics platform used a Vanquish UHPLC system coupled to a Q Exactive Orbitrap high-resolution mass spectrometer in Full-MS/ddMS2 mode. The workflow was designed to cover major plasma lipid subclasses, including phosphatidylcholines, sphingomyelins, phosphatidylethanolamines, lysophosphatidylcholines, ceramides, diacylglycerols, and triacylglycerols. Detailed chromatographic and mass spectrometric acquisition parameters have been reported in our previous study and are therefore not repeated here (*4*).

Lipid identification was based on high-resolution MS and MS/MS spectral information. Raw data were processed in LipidSearch v5.1 for deconvolution, chromatographic peak extraction, lipid identification, and retention-time alignment. The search type was Product Search. Positive-mode adducts included H+, Na+, and NH4+; negative-mode adducts included H-, CH3-, and C2H3O2-. Precursor and product-ion mass tolerances were set at <5 ppm and <8 ppm, respectively. Peak-alignment parameters included retention-time tolerance <0.05 min, retention-time correction tolerance <0.3 min, signal-to-noise ratio >3.0, and intensity ratio >1.5. Manual curation was restricted to lipid features with Rank = 1 and Grade >= C, with other parameters kept at default settings.

Lipidomics quality assurance and quality control covered sample collection, storage, extraction, mass-spectrometric acquisition, data processing, and reporting. Pooled plasma QC samples were prepared from equal-volume aliquots of homogenized study samples, aliquoted, and stored at -80 °C to avoid repeated freeze-thaw cycles. QC aliquots were extracted in each analytical batch and inserted into the injection sequence at fixed intervals to monitor temporal drift and batch effects. Instrument-level QC included routine Orbitrap and UHPLC maintenance, mass calibration, ion-source cleaning, injection-system maintenance, and column-pressure monitoring. QC metrics included electrospray ionization stability, chromatographic peak shape and baseline, retention-time reproducibility, peak-area reproducibility, lipid-subclass distribution, and feature-level coefficient of variation. Lipid features with poor QC reproducibility, high missingness, low signal-to-noise ratio, or suspected blank/background origin were removed before statistical analysis.

**S4. Clinical and PFAS association model specifications**

For clinical association analyses, each clinical index was modeled separately with ovarian cancer case status as the binary outcome. Four nested logistic models were fitted: Model 1, unadjusted; Model 2, adjusted for age; Model 3, adjusted for age and BMI and used as the primary clinical association model; and Model 4, adjusted for age, BMI, and hypertension and used as the comorbidity-adjusted sensitivity model. Depending on the distribution and prespecified analytical scale of the clinical variable, ORs corresponded to the original scale or the log2-transformed scale.

For single-PFAS analyses, each PFAS was entered separately into logistic regression models after log2 transformation and z standardization. PFAS group burdens were calculated by summing raw concentrations within predefined groups before log2 transformation and z standardization. The predefined PFAS burden groups were total PFAS, perfluoroalkyl carboxylic acids, perfluoroalkyl sulfonic acids, alternative PFAS, legacy PFAS, and emerging alternative PFAS.

**S5. Mixture exposure analysis details**

Extended mixture analyses used quantile g-computation, generalized weighted quantile sum regression, and BKMR as complementary observational approaches. Quantile g-computation was implemented using qgcomp.glm.noboot. PFAS exposures were coded as quartiles, and the model estimated the overall mixture log-odds ratio, OR, and component weights. Generalized weighted quantile sum regression also used quartile-coded exposures, 100 bootstrap samples, and a 60% validation-set split; positive- and negative-direction models were fitted when model estimation was feasible. BKMR was run for 20,000 iterations to estimate flexible exposure-response functions and posterior inclusion probabilities.

**S6. Observational mediation prioritization and tandem association-chain analysis**

Indirect associations were estimated as products of coefficients. Sobel P values and 300 bootstrap resamples were used to summarize evidence for indirect associations, bootstrap confidence intervals, and approximate mediation proportions. For tandem association chains, PFAS -> omics feature -> clinical phenotype -> case status pathways were decomposed to estimate total indirect effects, tandem indirect effects, and individual mediation components. Pathway evidence was summarized according to bootstrap results and FDR levels.

**S7. Internal prediction modeling and XGBoost sensitivity analysis**

Internal prediction modeling evaluated whether the Core-5 shared metabolites improved discrimination beyond clinical and tumor-marker features. Fixed logistic regression, glmnet LASSO, and glmnet elastic-net logistic regression were assessed using grouped stratified 5-fold cross-validation repeated 20 times. For penalized models, cv.glmnet was used within each outer training fold to select lambda.1se. LASSO used alpha = 1, and elastic-net used alpha = 0.5. Imputation, scaling, feature selection, and model fitting were restricted to the training fold and then applied to the corresponding validation fold to reduce information leakage.

XGBoost was used only as a nonlinear internal sensitivity analysis, not as external validation. XGBoost used grouped stratified 5-fold cross-validation repeated 10 times. Within each outer training fold, 20% of the training data were held out as an internal validation set for early stopping and selection of the optimal number of boosting rounds. The model was then refitted on the complete outer training fold using the selected number of rounds and evaluated in the corresponding validation fold. This analysis tested whether the incremental signal of the Core-5 panel was dependent on penalized regression models.
